# Safety and efficacy of antiseptic cleansing to reduce vertical transmission of multi-drug resistant pathogens to neonates (NeoVT-AMR)

**DOI:** 10.1101/2025.11.04.25333207

**Authors:** Emily Beales, Michelle N Clements, Nicholas A Feasey, David Lissauer, Maranatha Banda, Bertha Maseko, Julia A Bielicki, Samantha Lissauer, Aisleen Bennett, Kondwani Kawaza, Luis A Gadama, A Sarah Walker, Mike Sharland, Louise F Hill

## Abstract

**Importance:** Neonatal sepsis causes significant mortality. Antiseptic application to the perineum and vagina of labouring women or to neonatal skin could reduce neonatal pathogenic bacterial colonisation, lowering sepsis risk.

**Objective:** This study aimed to identify a safe, efficacious antiseptic regimen for a future effectiveness trial.

**Design:** Randomised controlled trial conducted between March 2022–March 2023. Laboratory staff (who assessed the primary outcome), were blinded to allocation.

**Setting:** The trial was conducted at Zomba Central Hospital, Malawi on the antenatal and labour wards (maternal stratum) and the neonatal and postnatal wards (neonatal stratum).

**Participants:** 149 women in labour and 147 neonates participated.

**Interventions:** Participants were individually randomised in an unblinded factorial design to antiseptic (chlorhexidine 1% [CHG1%], chlorhexidine 2% [CHG2%], octenidine 0.1%/phenoxyethanol

2% [OCT]) once or multiple times (maternal: ≤6 four-hourly applications; neonatal: ≤3 24- hourly applications postnatally) or to standard of care (SOC).

**Main Outcomes and Measures:** The primary outcome was change in total vaginal or neonatal skin bacterial load.

Secondary outcomes in both strata assessed safety by skin condition score, serious adverse events and temperature (neonatal strata only). Additionally, bacterial load of the neonates born to women in the maternal stratum was assessed.

**Results:** Mean baseline maternal vaginal bacterial load was 6.3 log_10_CFU (log_10_ colony forming units (log_10_CFU), SD=2.0), decreasing to 1.5 (SD=2.0) for CHG1% at 4hrs and 5.7 (SD=1.8) for SOC. CHG1% was more efficacious than OCT (absolute effect size=1.7 log_10_CFU, 95%CI[0.9,2.5], p<0.001) and SOC (effect size 3.5, 95%CI [2.4,4.5], p<0.001). There was no difference in efficacy between CHG1% and CHG2% or between application frequencies.

Mean baseline neonatal bacterial load was 4.3 log_10_CFU (SD=2.9), rising to 5.5 (SD=2.7) for CHG1% and 7.4 (SD=2.4) for SOC at 24 hours. CHG1% showed greater efficacy than SOC (effect size=1.2, 95%CI [0.2,2.3], p=0.023), with no difference between antiseptics. Multiple applications showed increasing benefits over time.

No safety concerns were observed.

**Conclusion and Relevance:** CHG1% was efficacious in both strata, with effect sizes higher in the maternal stratum. This suggests chlorhexidine cleanses could be incorporated into future multimodal infection prevention intervention trials.

**Trial registration:** NeoVT-AMR is registered on the ISRCTN registry (ISRCTN78026255), registration date 26th May 2022. Protocol and statistical analysis plan available at: https://doi.org/10.1186/ISRCTN78026255.

**Key points:** Question: is antiseptic application in labouring women and neonates safe and efficacious at reducing bacterial load?

Findings: this randomised clinical trial included 149 women in labour and 147 neonates. 1% chlorhexidine was efficacious in reducing bacterial load in both strata, with effect sizes higher in the maternal stratum. No safety concerns were observed.

Meaning: chlorhexidine cleanses could be incorporated into multimodal infection prevention intervention trials in neonates and labouring women.

## Introduction

Infection is a major cause of neonatal mortality, with serious bacterial infections (SBIs) responsible for around 680,000 neonatal deaths per year in low-income countries (LICs).^1^ Gram-negative pathogens, such as *Klebsiella* spp. and *Escherichia coli*, commonly cause SBIs in low- and middle-income countries (LMICs).^2^ The majority of SBIs identified in neonates in LICs, including Malawi, are due to early onset sepsis (EOS), occurring within 72 hours of birth.^3,4^ A major mechanism of acquisition of EOS pathogens is transfer from the maternal perineum and vagina during labour, i.e., vertical transmission.^5^

New strategies to reduce neonatal SBIs are urgently required. Antiseptics, when applied vaginally to labouring mothers or topically to neonates, are a potential method to reduce vertical transmission and colonisation with resistant pathogens. Antiseptics are inexpensive, widely used, generally well-tolerated, and easily administered, including in resource-limited settings. The most widely used antiseptics in maternal and neonatal populations are chlorhexidine (CHG), and octenidine 0.1% combined with phenoxyethanol 2% (OHP) (trade name Octenisept®). CHG is commonly applied to the skin before procedures on neonatal units and significantly reduces skin bacterial load.^6^ OHP is used to treat bacterial vaginosis and is the routine antiseptic on many neonatal units in Europe.^7^ Importantly, pre-clinical reports have shown higher activity of OCT against Gram-negative organisms compared to CHG.^8,9^ A large South African randomised controlled trial (RCT) of vaginal and neonatal CHG wipes to prevent early-onset Group B Streptococcus (GBS) disease found no evidence of an effect on culture positive or clinically diagnosed EOS.^10^ However, this trial was conducted 20 years ago when EOS epidemiology differed substantially, there was low incidence of culture positive EOS and it adopted a low concentration of 0.5% CHG.^10,11^ Large, non-randomised studies of CHG vaginal administration have reported reductions in infection-related mortality in infants.^12,13^ Although CHG may reduce neonatal sepsis; there is a lack of evidence for the impact of concentration^14^ and prevention of non-GBS EOS.^15^ The impact of antiseptic use on maternal infections should also be considered. It is estimated that obstetric infections are responsible for ∼11% of maternal deaths with infection prevention key to reducing infection-related maternal morbidity and mortality.^16^ Application of CHG vaginally has been shown in RCTs to reduce maternal infectious morbidity and shorten hospital stay when delivering via Caesarean section (C-section).^17,18^ Large pragmatic trials are required to investigate whether antiseptic use in neonates and labouring women reduces neonatal sepsis and maternal infections in moderate-high incidence settings, particularly with highly resistant organisms. However, uncertainty exists about optimal antiseptic choice, dosing, and administration frequency and whether maternal or neonatal administration is efficacious.

This trial aimed to investigate different topical antiseptic strategies in labouring women and newborns to reduce bacterial load in the female genital tract and on the skin of newborns.

Results from this trial will inform the design of a planned pragmatic clinical trial of antiseptic use to prevent EOS in neonates in LMICs.

## Methods

### Study design

NeoVT-AMR was an unblinded, factorial, individually randomised controlled trial with two independent strata: labouring women (maternal stratum) and neonates (neonatal stratum). Mothers and neonates were each allocated 1:1:1:1:1:1:1 in a 3x2+1 factorial with six intervention arms and one standard of care (SOC) arm (eFigure 1 in Supplement). The SOC arm was included as a benchmark to distinguish between potential scenarios of all treatments being equally efficacious or equally inefficacious. The trial was conducted at Zomba Central Hospital (ZCH), Zomba, Malawi on the antenatal and labour wards (maternal stratum) and the neonatal and postnatal wards (neonatal stratum). The trial received ethical approval from St George’s, University of London, UK, Research Ethics Committee (2020.0344), the Kamuzu University of Health Sciences, Blantyre, Malawi, Research and Ethics Committee (P.01/21/3248) and regulatory approval from The Pharmacy and Medicines Regulatory Authority of Malawi (PMRA/CTRC/IV/26072021132). Written informed consent was obtained from all women and parent(s)/guardian(s) of neonates prior to trial entry. The study was performed in accordance with the International Conference on Harmonisation of Technical Requirements for Registration of Pharmaceuticals for Human Use Good Clinical Practice Guidelines. Protocol amendments are outlined in the Supplement (eAppendix 1).

**Figure 1.**
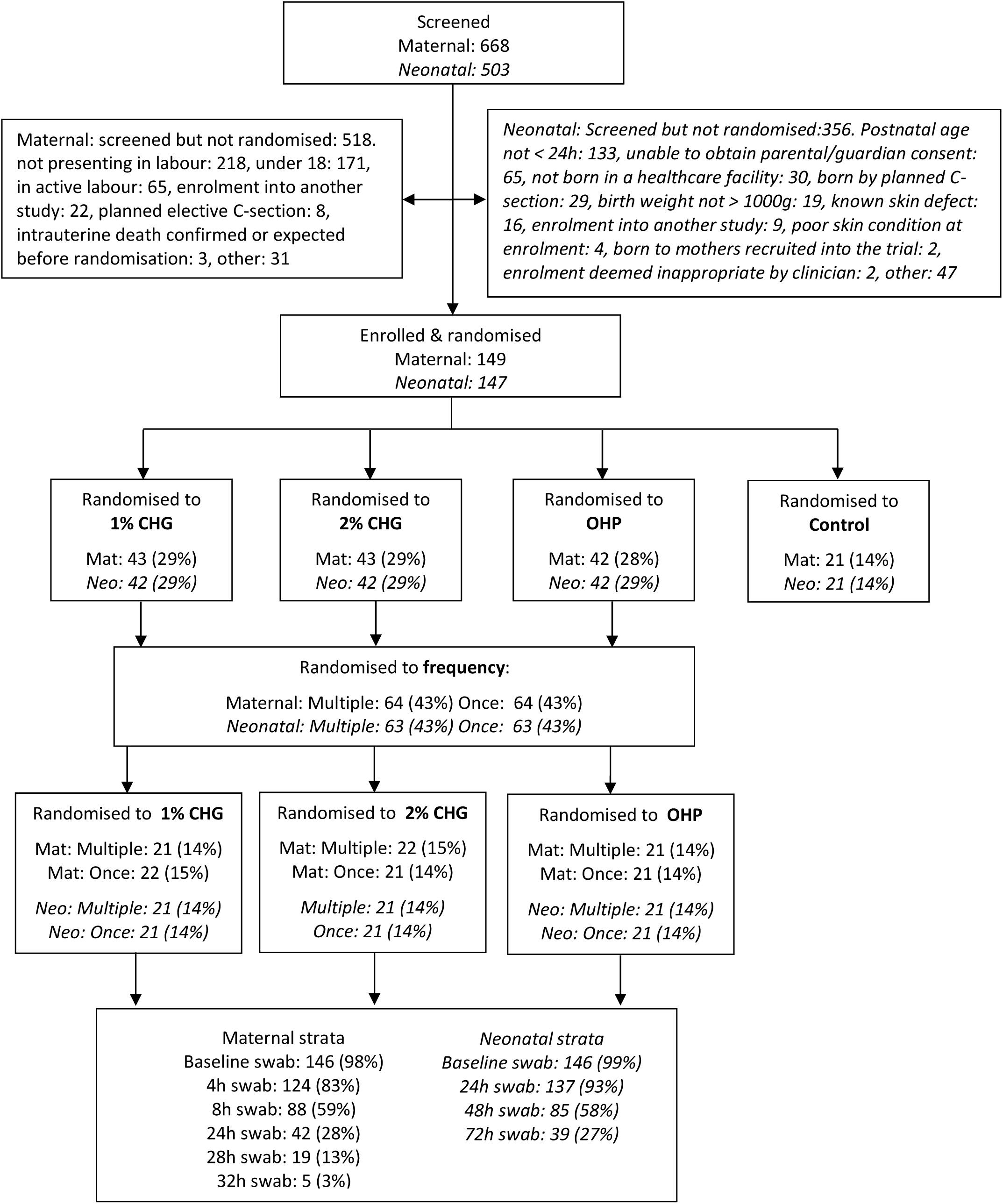
**CONSORT diagram for maternal and neonatal strata. Maternal: non-italic ; neonatal : italic**

### Participants

Eligible mothers were those presenting in labour, with or without rupture of membranes at time of randomisation. Exclusion criteria were mothers in active labour (regular contractions and/or cervical dilatation of >4cm), planned elective C-section delivery, minors (<18 years), contra-indication to digital vaginal examination, poor perineal and vaginal skin condition as assessed by enrolling research nurse, known or suspected allergy to CHG or OCT or phenoxyethanol, any recent or planned (within four hours) iodine application to the perineum or vagina (due to skin discolouration reaction with octenidine), expected or confirmed intrauterine death, unable to obtain consent, and antiseptic application or enrolment determined inappropriate by the enrolling clinician.

Eligible neonates were those born in a healthcare facility, <24 hours old (postnatally) with a birth weight >1000g. Exclusion criteria were neonates born to mothers recruited to the trial, born by elective C-section, poor skin condition (skin score ≥2 in any of the three domains (section 3.1 eMethods in Supplement), congenital/acquired skin disorder/defect at time of randomisation, recent or planned (within four hours) iodine application to body, planned/previous lumbar puncture, unable to obtain parental/guardian consent, antiseptic application or enrolment determined inappropriate by the enrolling clinician. The neonatal inclusion criteria were amended after trial commencement, from postnatal age at randomisation <12 hours to <24 hours and from “born in Zomba Central Hospital” to “born in a healthcare facility” to support neonatal enrolment and to better reflect the ZCH neonatal population (eAppendix 1 in Supplement).

### Randomisation and masking

Participants were randomised by a computer-generated list created by the Trial Statistician using random permuted blocks of size 13. Trial nurses enrolled participants and used the web-based database (RedCAP) to randomise participants. All trial activities including eligibility assessment, randomisation, intervention, and follow-up were conducted by specific trial staff. Due to the nature of the interventions, research personnel and participants/parents or guardians were not masked to regimen allocation. The laboratory team assessing the co-primary endpoint were blinded to randomisation.

### Procedures

In both the maternal and neonatal strata, the intervention arms assessed three different antiseptics (1% CHG (CHG1%), 2% CHG (CHG2%) or OHP), applied either once only (at 0 hours) or multiple times.

In the maternal stratum, the multiple application arms received antiseptic at four hourly intervals (0, 4, 8, 24, 28, 32 hours from randomisation) or until delivery if earlier than 32 hours. The SOC arm had sterile water applied as recommended by the World Health Organization (WHO)^19^ but not routinely practised in ZCH. The antiseptic or sterile water was applied to the vagina and perineum with soaked cotton wool using the 6-swab technique (section 3.2 eMethods in Supplement). In the neonatal stratum, the multiple application arms received antiseptic at 24 hourly intervals for up to 4 applications (0, 24, 48, 72 hours from randomisation) or until discharge home, whichever was earlier. Antiseptic was applied to the whole body with soaked cotton wool, excluding the face and scalp. The SOC arm received no cleanses.

Swabs were taken from the vagina and perineum (maternal stratum) and neck skin fold and peri-rectal area (neonatal stratum) at each treatment timepoint regardless of allocated arm, i.e., 0, 4, 8, 24, 28, 32 hours from randomisation for the maternal stratum and 0, 24, 48, 72 hours for the neonatal stratum. Swabs were taken prior to any antiseptic or water application where applicable. Swabs were placed in liquid amies transport media. Swabs in liquid amies transport media were vortexed and serial dilutions performed to 10^-4^. Neat, 10^-2^ and 10^-4^ dilutions were plated onto Uriselect and sheep blood agars and incubated overnight at 35°C in air. Colony counts were performed, and log colony-forming units (log_10_CFU) calculated. Organism identification and antibiotic susceptibility testing were undertaken. Full microbiological methods can be found in the Supplement (section 3.3 in eMethods).

### Outcomes

The primary outcome measure in both strata was change from baseline in total bacterial load in log_10_CFUs.

Secondary outcomes in both strata assessed safety by skin condition score (section 3.1 eMethods in Supplement) and serious adverse events (SAEs); using the Common Terminology Criteria for Adverse Events in the maternal stratum (eTable 2 in Supplement) and an adapted DAIDS score for neonates (eTable 3 in Supplement). Skin condition score was measured at baseline and at each treatment timepoint in all participants, regardless of whether treatment administration was required for their allocated arm and prior to any randomised treatment, where applicable. Neonates receiving SOC had a skin assessment once daily during study participation. SAEs were recorded whilst participants were admitted to hospital and following discharge via telephone on Day 28. An additional safety outcome in the neonatal stratum assessed temperature, both as change from pre- to post-treatment application and graded measuring hypothermia; temperature assessment was completed once daily for those receiving SOC. An additional secondary outcome assessed bacterial load (neck skin fold and peri-rectal) in the neonates born to women in the maternal stratum.

Additional efficacy outcomes specified in the Statistical Analysis Plan were log_10_CFUs of Gram-positive, Gram-negative and yeast species.

### Statistical analysis

A target sample size of 147 participants per stratum gave 90% power to detect a difference of 0.82 standard deviations (SDs) between antiseptics (two-sided α=0.012) and 0.58 SDs between frequencies (α=0.05) (80% power for 0.72 and 0.50 SDs, respectively). This also provided 90% power to detect a difference of 1.01 SD between each antiseptic and SOC (α=0.012), and 0.90 SD between frequency and SOC (α=0.025) (0.88 and 0.79 SDs at 80% power). These expected effects were judged by clinical and statistical members of the team to be sufficient for selecting a treatment regimen for a future trial given previous observed data showing decreases in log_10_CFU from baseline to 24h ranging from 0.2 SD to 2 SD.^6,20^ A Data Monitoring Committee assessed safety endpoints halfway through recruitment.

All analyses were ‘intention-to-treat’ (‘treatment policy’) in the Estimands framework. Each stratum was analysed independently. All participants with a baseline measurement and at least one measurement post-baseline were included in the analyses of the primary outcome for each stratum. Total log_10_CFU change was analysed using mixed effect models with normally distributed errors, including fixed effects for antiseptic (CHG1%, CHG2%, OHP); application frequency (once, multiple); hour of assessment (linear; deviation from linearity assessed using multivariable fractional polynomials); and baseline value (estimate not interpretable by design due to regression to the mean; deviation from linearity assessed using multivariable fractional polynomials), with comparisons using Wald tests. Individual was fitted as a random effect with unstructured covariance if this was a better fit to the data. As the goal was to identify a cleansing regimen to take forward to a larger trial, the SOC arm was included only as a benchmark, with smaller sample size, and was not used as the reference group. The reference group was a single application of CHG1% as this is the simplest intervention to implement in a large-scale clinical trial.

Interaction analyses between the co-primary outcomes were assessed one at a time in separate models and sensitivity of the models was assessed by analysing only the first measure after the baseline and the final measure after baseline. Pre-specified subgroup analyses fitted the interaction between each arm and the relevant subgroup factor one at a time in separate models. Bayesian ACCEPT analysis^21^ estimated the probability that total log_10_CFU truly differed from the reference group by different values (priors in section 3.5 eMethods in Supplement).

Skin score change, temperature change from pre- to post-application (treatment arms in neonatal stratum only) were analysed as per the primary outcome. SAEs and AEs were summarised overall and by MedDRA System Order Class and Preferred Term and compared using exact logistic regression.

## Results

Between 7^th^ March 2022 and 29^th^ March 2023, 149 women and 147 neonates were randomised to the maternal and neonatal stratum, respectively (Figure 1). The trial ended when the pre-defined sample size of 147 was met.

The average age at enrolment was 25.7 years (SD=5.9; Table 1) in the maternal stratum and 10.3 hours (SD=6.3) in the neonatal stratum, with estimated gestational age at labour 38 weeks (SD=1.5) in the maternal stratum and 37 (SD=3) in the neonatal stratum. In the maternal stratum, antibiotics were given to 26/149 (17%) of women during pregnancy and 5/149 (3%) women during labour. In the neonatal stratum, the mean birthweight was 2.7kg (SD=0.7kg) and 76/147 (52%) births were via spontaneous vaginal delivery with 61/147 (41%) via emergency C-section. In the neonatal stratum before enrolment, 31/147 (21%) neonates had received antibiotics, and 112/147 (76%) neonates had chlorhexidine cord care. Additional baseline characteristics are outlined in section 4.1 eResults in Supplement.

**Table 1.** Participant baseline characteristics.

|  |  | Overall | 1% CHG<br>(N=43) | 2% CHG<br>(N=43) | OHP<br>(N=42) | Once<br>(N=64) | Multiple<br>(N=64) | SOC<br>(N=21) |
| --- | --- | --- | --- | --- | --- | --- | --- | --- |
| **Maternal stratum** |  |  |  |  |  |  |  |  |
| Age at randomisation (years) | Mean (SD) | 25.7 (5.9) | 26.3 (6.6) | 25.4 (6.1) | 25.7 (5.6) | 27.5 (6.6) | 24.1 (5.0) | 24.8 (5.0) |
| Estimated gestational age at labour | Mean (SD) | 37.7 (1.5) | 37.8 (1.3) | 37.8 (1.6) | 37.4 (1.4) | 37.9 (1.2) | 37.5 (1.6) | 37.6 (1.9) |
| Antenatal care during pregnancy | N (%) | 144<br>(97%) | 43<br>(100%) | 43<br>(100%) | 37 (88%) | 62 (97%) | 61 (95%) | 21 (100%) |
| Tuberculosis | N (%) | 2 (1%) | 0 (0%) | 1 (2%) | 1 (2%) | 0 (0%) | 2 (3%) | 0 (0%) |
| HIV | N (%) | 14 (9%) | 5 (12%) | 4 (9%) | 4 (10%) | 10 (16%) | 3 (5%) | 1 (5%) |
| Antibiotics during pregnancy | N (%) | 26 (17%) | 5 (12%) | 10 (23%) | 9 (21%) | 13 (20%) | 11 (17%) | 2 (10%) |
| Antibiotics since admission | N (%) | 5 (3%) | 3 (7%) | 0 (0%) | 2 (5%) | 3 (5%) | 2 (3%) | 0 (0%) |
| Total log<sub>10</sub> CFU: baseline | Mean (SD) | 6.3 (2.0) | 6.2 (1.9) | 6.3 (1.9) | 6.6 (2.1) | 6.4 (1.7) | 6.3 (2.2) | 6.0 (2.1) |
| **Neonatal stratum** |  |  |  |  |  |  |  |  |
| Age at enrolment (hours) | Mean (SD) | 10.3 (6.3) | 10.8 (5.9) | 10.6 (6.6) | 9.4 (6.0) | 10.3 (6.1) | 10.3 (6.3) | 10.6 (7.2) |
| Estimated gestational age at labour (weeks) | Mean (SD) | 36.7 (3.1) | 36.9 (2.9) | 36.6 (3.1) | 37.0 (3.1) | 36.5 (3.0) | 37.2 (3.0) | 36.0 (3.1) |
| Sex | Female | 64 (44%) | 17 (40%) | 21 (50%) | 17 (40%) | 26 (41%) | 29 (46%) | 9 (43%) |
|  | Male | 82 (56%) | 24 (57%) | 21 (50%) | 25 (60%) | 37 (59%) | 33 (52%) | 12 (57%) |
|  | Unknown | 1 (1%) | 1 (2%) | 0 (0%) | 0 (0%) | 0 (0%) | 1 (2%) | 0 (0%) |
| Birth weight (g) | Mean (SD) | 2729<br>(712) | 2661<br>(671) | 2751<br>(731) | 2877<br>(724) | 2739<br>(690) | 2787<br>(732) | 2528 (713) |
| Where admitted | Nursery | 50 (34%) | 14 (33%) | 17 (40%) | 13 (31%) | 24 (38%) | 20 (32%) | 6 (29%) |
|  | Nursery HDU | 50 (34%) | 13 (31%) | 11 (26%) | 16 (38%) | 20 (32%) | 20 (32%) | 10 (48%) |
|  | Postnatal ward with Mother | 46 (31%) | 14 (33%) | 14 (33%) | 13 (31%) | 19 (30%) | 22 (35%) | 5 (24%) |
|  | Unknown | 1 (1%) | 1 (2%) | 0 (0%) | 0 (0%) | 0 (0%) | 1 (2%) | 0 (0%) |

|  |  | **Overall** | **1% CHG**<br>**(N=43)** | **2% CHG**<br>**(N=43)** | **OHP**<br>**(N=42)** | **Once**<br>**(N=64)** | **Multiple**<br>**(N=64)** | **SOC**<br>**(N=21)** |
| --- | --- | --- | --- | --- | --- | --- | --- | --- |
| Reason for admission | Birth asphyxia | 48 (33%) | 13 (31%) | 12 (29%) | 16 (38%) | 20 (32%) | 21 (33%) | 7 (33%) |
|  | Other | 54 (37%) | 15 (36%) | 17 (40%) | 13 (31%) | 25 (40%) | 20 (32%) | 9 (43%) |
|  | Routine admission | 44 (30%) | 13 (31%) | 13 (31%) | 13 (31%) | 18 (29%) | 21 (33%) | 5 (24%) |
|  | Unknown | 1 (1%) | 1 (2%) | 0 (0%) | 0 (0%) | 0 (0%) | 1 (2%) | 0 (0%) |
| Any antibiotics prescribed before enrolment since birth | Yes | 31 (21%) | 8 (19%) | 7 (17%) | 10 (24%) | 12 (19%) | 13 (21%) | 6 (29%) |
|  | No | 115 (78%) | 33 (79%) | 35 (83%) | 32 (76%) | 51 (81%) | 49 (78%) | 15 (71%) |
|  | Unknown | 1 (1%) | 1 (2%) | 0 (0%) | 0 (0%) | 0 (0%) | 1 (2%) | 0 (0%) |
| Chlorhexidine cord care given | Yes | 112 (76%) | 32 (76%) | 32 (76%) | 33 (79%) | 50 (79%) | 47 (75%) | 15 (71%) |
|  | No | 34 (23%) | 9 (21%) | 10 (24%) | 9 (21%) | 13 (21%) | 15 (24%) | 6 (29%) |
|  | Unknown | 1 (1%) | 1 (2%) | 0 (0%) | 0 (0%) | 0 (0%) | 1 (2%) | 0 (0%) |
| Mother received antibiotics in labour | Yes | 59 (40%) | 16 (38%) | 20 (48%) | 18 (43%) | 29 (46%) | 25 (40%) | 5 (24%) |
|  | No | 77 (52%) | 25 (60%) | 19 (45%) | 21 (50%) | 31 (49%) | 34 (54%) | 12 (57%) |
|  | Unknown | 11 (7%) | 1 (2%) | 3 (7%) | 3 (7%) | 3 (5%) | 4 (6%) | 4 (19%) |
| Total log<sub>10</sub> CFU: baseline | Mean (SD) | 4.3 (2.9) | 4.2 (2.8) | 4.3 (3.1) | 3.9 (2.8) | 3.9 (2.5) | 4.4 (3.2) | 5.3 (3.4) |

Individuals assigned to non-standard of care (SOC) arm appear twice in the table: in the randomised antiseptic and frequency. One neonate in the neonatal stratum was withdrawn before any baseline data were collected and is shown as unknown.

Compliance with the protocol was good with no individuals in the SOC arms receiving antiseptic application and no individuals in the single application arms receiving multiple applications (section 4.2 eResults in Supplement). The median number of antiseptic applications in the multiple application arm was 3 (IQR: 2, 3)) in both maternal and neonatal stratum. Rates of follow-up by telephone at Day 28 were 104/149 (70%) in the maternal stratum and 114/147 (78%) in the neonatal stratum.

In the maternal stratum, 98% (146/149) of women had a baseline swab, 83% (124/149) at 4h and 59% (88/149) at 8h (where differences between frequency arms may start to be detected); just 3% of women were swabbed at 32h (Figure 1). In the neonatal stratum, 99% of neonates (146/147) were swabbed at baseline, 93% (137/147) at 24h (137/147), 58% at 48h (85/147) (where differences between frequency arms may start to be detected) and 27% (39/147) had a 72h swab.

In the maternal stratum, compared to CHG1%, analysis of all data showed higher average total log_10_CFU with both OHP (effect size=1.7, 95% CI: (0.9, 2.5), p <0.0001; Table 2; Figure 2) and SOC (effect size=3.5, 95% CI: (2.4, 4.6), p <0.0001); there was no robust evidence that total log_10_CFU differed with CHG2% (effect size=-0.6, 95% CI: (-1.4, 0.2), p=0.12, Figure 2, Table 2). There was no evidence of differences with application frequency although the trend was towards lower total log_10_CFU in the multiple applications arm (effect size=-0.4, 95% CI: (-1.1, 0.2), p=0.20). Results were qualitatively similar for Gram-negative pathogens (Table 2) plus Gram-positive and yeast, including individual species (section 4.3 eResults in Supplement). There was no evidence of interactions between treatment arms (section 4.4 eResults in Supplement) nor for pre-specified subgroup effects (section 4.5 eResults in Supplement).

**Figure 2.**
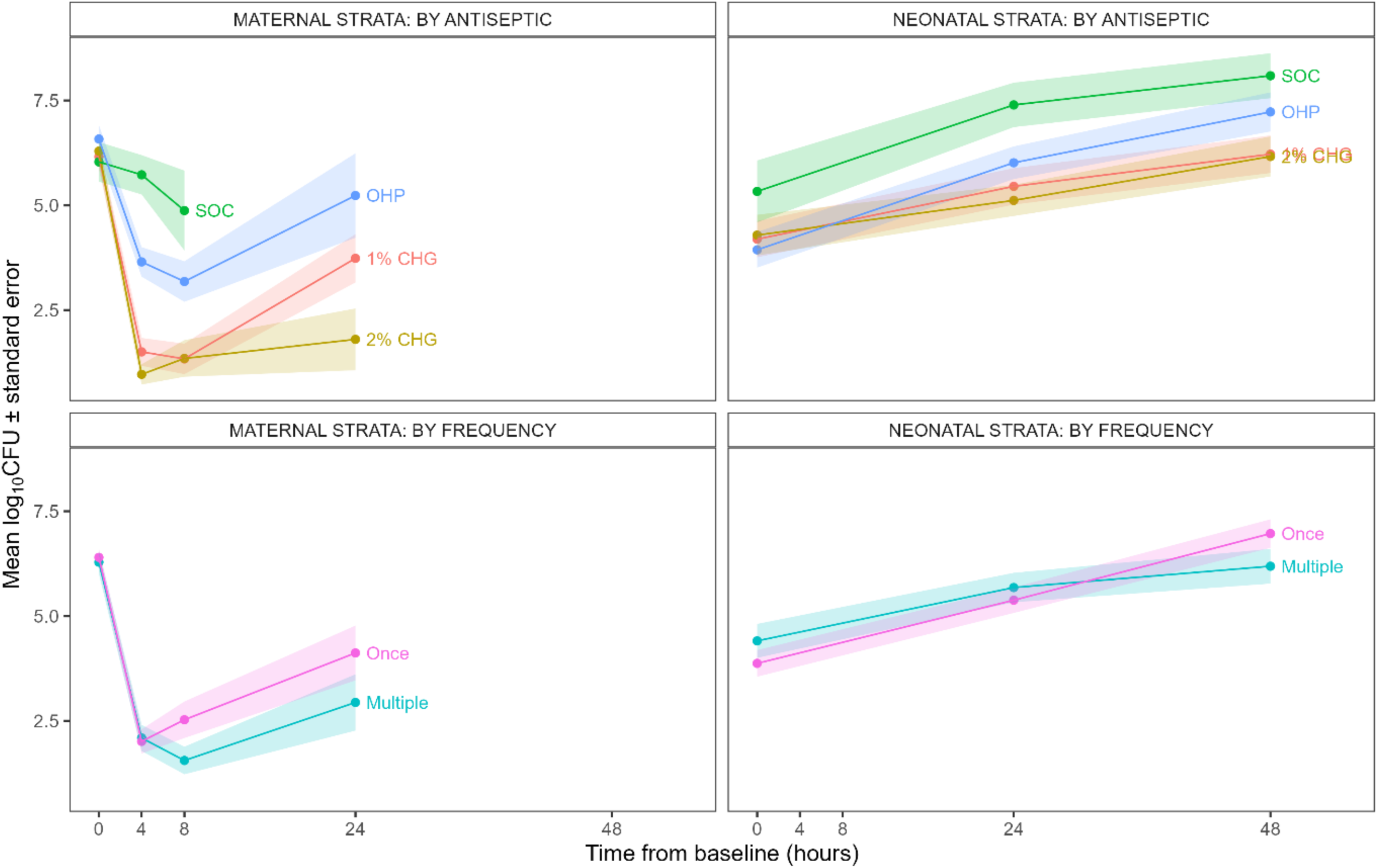
Total log10CFU over time by strata and arm. Note that data from non-standard of care (SOC) factorial arms (antiseptic and frequency) are averaged across the other factorial arm (frequency/antiseptic, respectively) and so participants in antiseptic arms appear twice (in antiseptic and frequency graphs). Datapointscomprising fewer than 10 participants removed for clarity. Time 0 corresponds to enrolment.

**Table 2.**
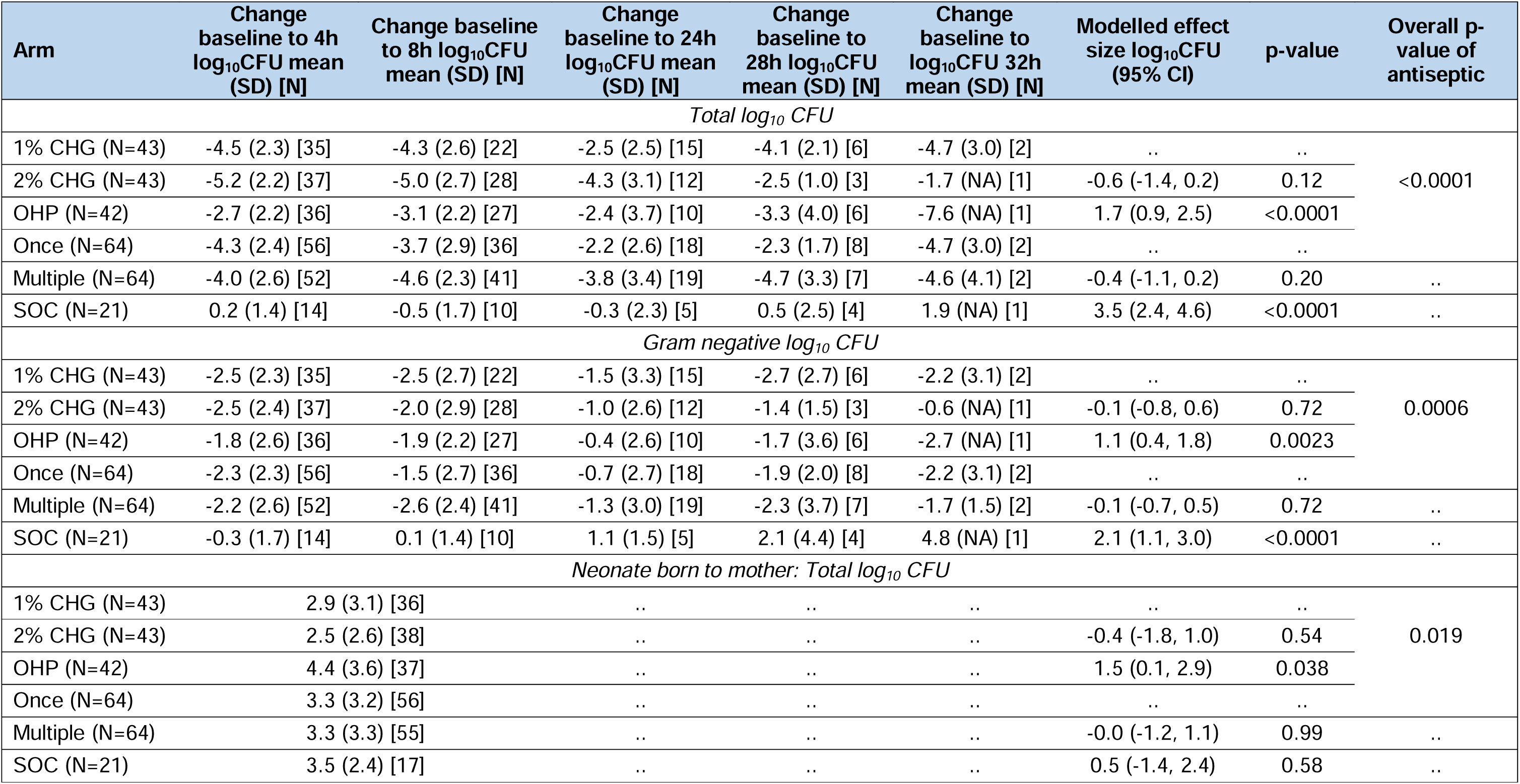
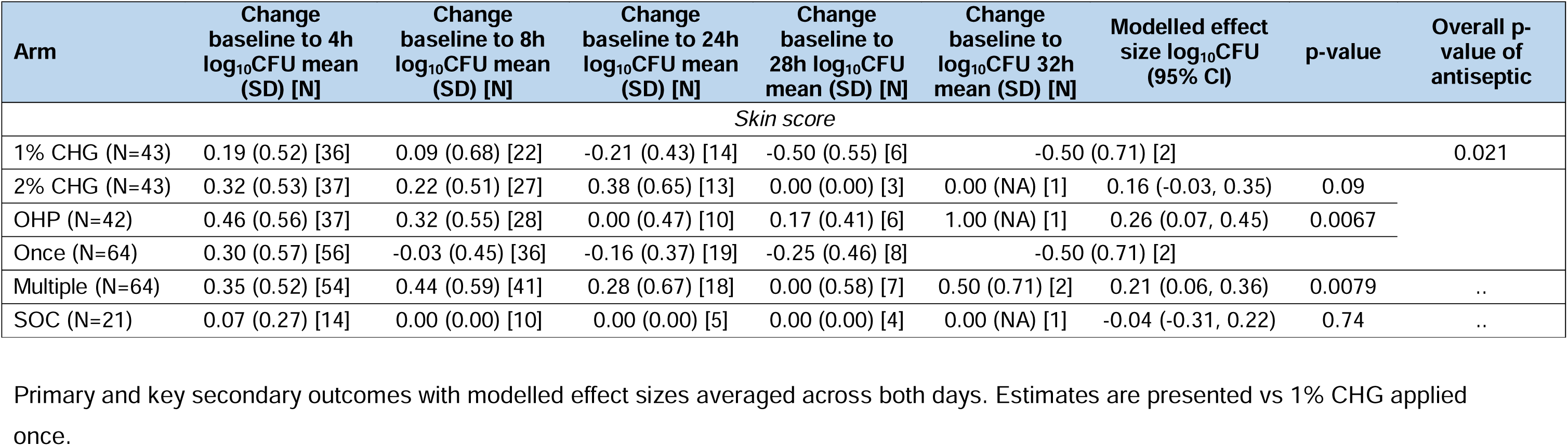
Maternal efficacy outcomes.

Note that data from non-standard of care (SOC) factorial arms (antiseptic and frequency) are averaged across the other factorial arm (frequency/antiseptic, respectively) and so participants in antiseptic arms appear twice (in antiseptic and frequency graphs). Datapoints comprising fewer than 10 participants removed for clarity. Time 0 corresponds to enrolment.

In the neonatal stratum, analysis of all swabs together compared to CHG1% showed no evidence of different average log_10_CFU with either CHG2% (effect size=-0.2, 95% CI: (-1.1, 0.7), p=0.63; Table 3) or OHP (effect size=0.7, 95% CI: (-0.2, 1.6), p=0.11) although the trends were in the same direction as the maternal stratum; total log_10_CFU was higher in the SOC arm compared to CHG1% (effect size=1.3, 95% CI: (0.2, 2.4), p=0.017). There was no evidence of a difference with application frequency (effect size=-0.5, 95% CI: (-1.2, 0.2), p=0.17) or evidence of differences between arms for Gram-negative pathogens (Table 3; individual species outcomes are shown in the Supplement (section 4.3 eResults). There was evidence of an interaction between multiple frequencies and time, with the benefits of multiple applications becoming greater over time (section 4.4 eResults in Supplement). Total log_10_CFU was higher in neonates born to mothers with HIV but there was no evidence of interactions between HIV status and treatment arms. There was, however, evidence of interactions between frequency of treatment and both admission ward and antibiotics given before enrolment (section 4.5 eResults in Supplement).

**Table 3.**
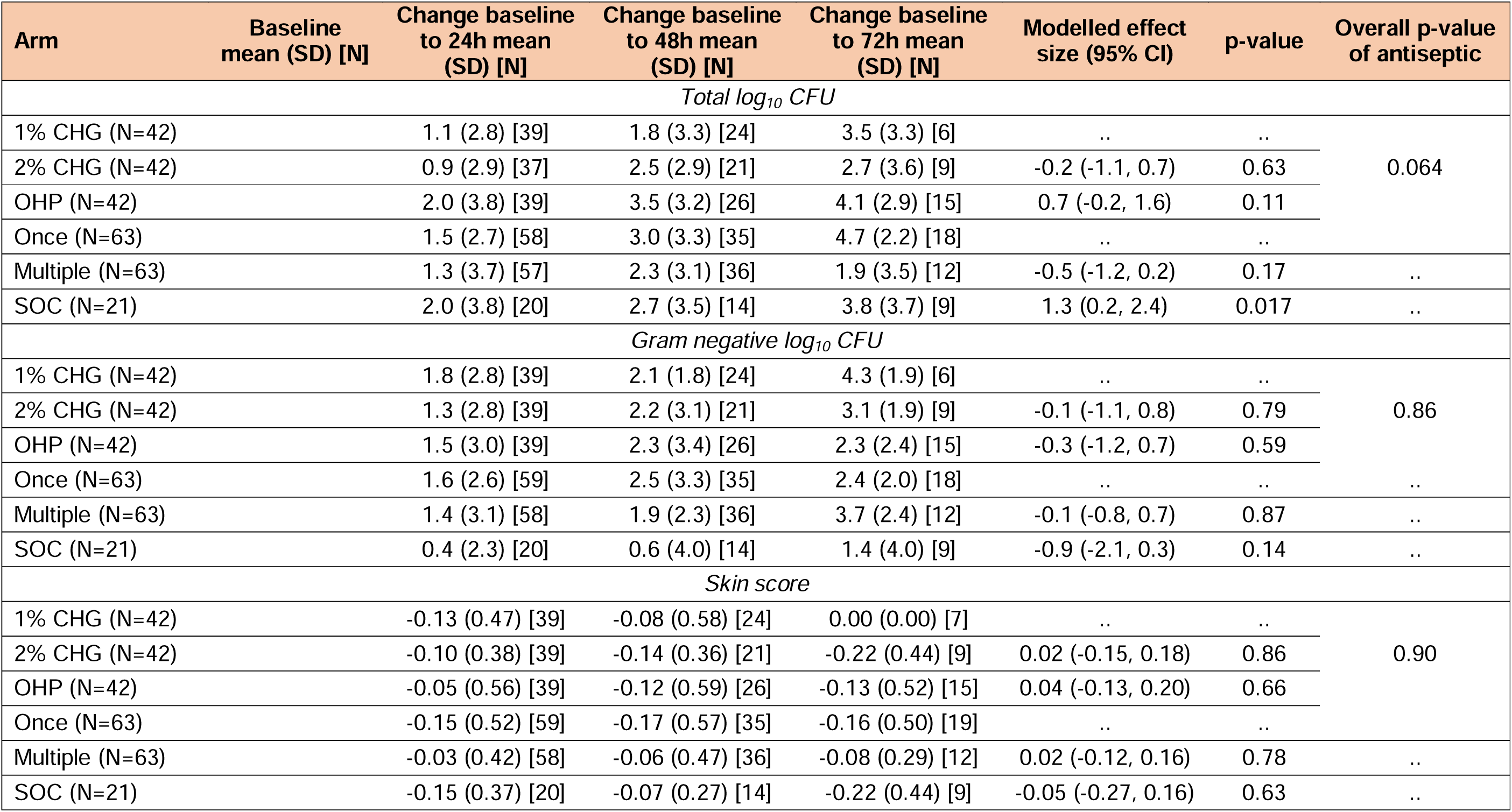
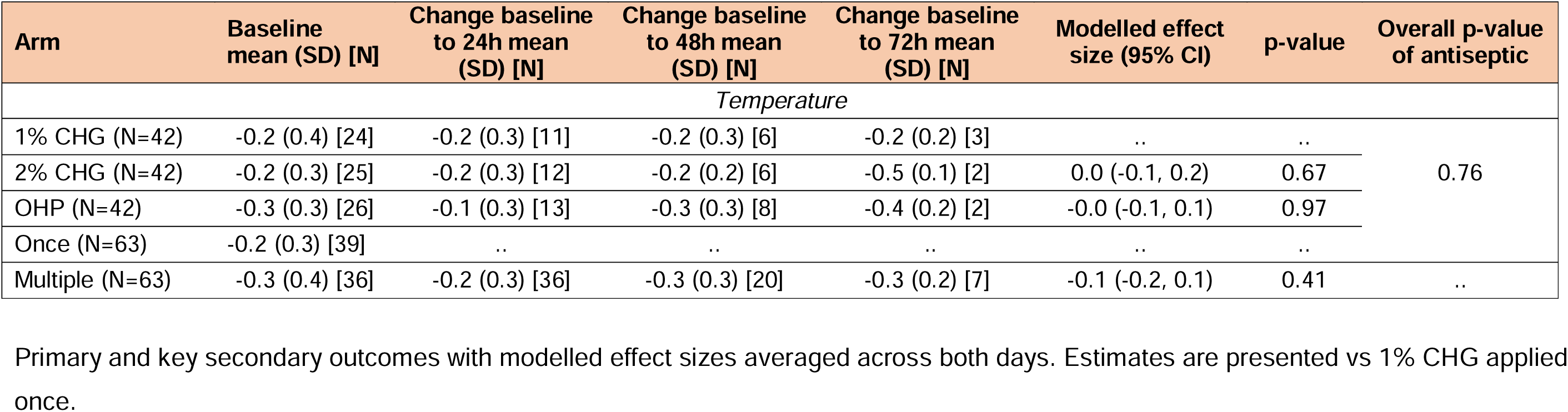
Neonatal efficacy outcomes.

Bayesian ACCEPT analysis (section 4.6 eResults in Supplement) showed a 94% and 63% probability that CHG2% was better than CHG1% in the maternal and neonatal strata, respectively, dropping to 62% and 22% when the probability of CHG2% being better than CHG1% by at least 0.5log CFU was considered. The probability of being better than CHG1% is 0% and 4% for OHP and 0% and 1% for those receiving SOC in the maternal and neonatal strata respectively. The probability of multiple applications being better than single was 81% in the maternal stratum and 82% in the neonatal stratum. Pre-specified sensitivity analyses showed similar patterns to the main analyses (section 4.7 eResults in Supplement).

In neonates born to mothers in the maternal stratum, similar trends to total log_10_CFU in the mothers persisted across antiseptic arms, with higher log_10_CFU in the OHP arm compared to the CHG1% arm (effect size=1.5, 95% CI: (0.1, 2.9), p=0.038; Table 2) but no evidence of a difference between CHG1% and CHG2% (effect size=-0.4, 95% CI: (-1.8, 1.0), p=0.054).

However, there was no evidence of any trends in difference between multiple and single applications (effect size=0.0, 95% CI: (-1.2, 11), p=0.99) nor between 1% and SOC (effect size=0.5, 95% CI: (-1.4, 2.4), p=0.58). Total bacterial load was higher in neonates born to mothers without HIV and spontaneous vaginal births though there was no evidence of any interactions with treatment arms (section 4.5 eResults in Supplement).

Skin scores in both strata were broadly 0 or 1 (out of 16 in the maternal stratum and 12 in the neonatal stratum), with just three instances of a skin score of 2 in the maternal stratum and five instances in the neonatal stratum and no instances of skin scores of 3 or more.

Statistical analysis in the maternal stratum (Table 2) showed higher scores in the OHP arm compared to the CHG1% arm (effect size=0.26, 95% CI: (0.07, 0.43), p=0.0067), no evidence of differences between the CHG1% and CHG2% arms (effect size=0.16, 95% CI: (-0.03, 0.35), p=0.09), higher scores in the multiple application arm compared to the single application arm (effect size=0.21, 95% CI: (0.06, 0.36), p=0.0079) but no evidence of a difference in skin scores between CHG1% and SOC (effect size=-0.04, 95% CI: (-0.31, 0.22), p=0.74). There was no evidence of differences between arms in neonatal temperature after antiseptic application (Table 3).

Seven SAEs were recorded in 7 women in the maternal stratum, with 6 relating to the neonate born to the mother, and 17 SAEs in 16 neonates in the neonatal stratum (sections 4.8, 4.9 eResults in Supplement). There was no evidence that rates of participants experiencing SAEs differed between the arms in either stratum although rates in the neonatal stratum were numerically higher in the CHG2% arm. There were no SAEs related to the antiseptic or grade 3 or 4 skin scores reported in either stratum. In the neonatal stratum, there were two instances of grade 3/4 temperature (below 35°C). One (34.4°C) was in a neonate in the SOC arm and so had not received antiseptic. The other (34.9°C) was in a neonate at the baseline assessment who received a single CHG1% application; the neonate was warmed to 36.5°C prior to antiseptic application and the temperature after application was 35.9°C.

## Discussion

NeoVT-AMR, a factorial RCT aimed to identify an efficacious antiseptic regimen, including optimal antiseptic concentration and application frequency, that could be safely evaluated in a future clinical trial aiming to reduce mortality associated with neonatal sepsis in low-income settings. The trial used a surrogate efficacy outcome of change in total bacterial load from baseline and was conducted in labouring women and neonates. CHG1% was safe and efficacious in both strata and could be taken forward to a future trial with clinical endpoints.

Lower total bacterial load was observed with CHG1% applied once compared to SOC in both strata. This trend was also observed for Gram-negative organisms, Gram-positive organisms, *Enterococcus* spp. and *Acinetobacter* spp. in the maternal stratum and Gram-positive organisms and *Klebsiella* spp. in the neonatal stratum. This is of particular importance as methods to prevent *Klebsiella* spp. infection in neonates are urgently needed, due to a drastic increase in resistance across Sub-Saharan Africa, including Malawi.^22^ The magnitude of effect of the primary endpoint was larger for the maternal group, perhaps reflecting the more frequent assessments (4 hourly in the maternal stratum and 24 hourly in the neonatal stratum), differences in the microbiological environment of the skin and vagina, or variations in the environmental exposures of mothers and neonates within the hospital.

These results show similar patterns to other studies investigating antiseptics in neonates^23–27^ and labouring women^28^ indicating positive effects on reducing colony counts with the use of CHG but with a strong rebound effect in neonates and possible reduced activity on Gram-negative organisms. There was, however, no robust evidence that the higher 2% concentration of CHG led to reduced bacterial load when compared to CHG1% in either stratum but reduced yeast colonisation was seen in the maternal group. These results are consistent with NeoCHG,^25^ a similar trial conducted in hospitalised neonates in Bangladesh and South Africa, which compared 0.5%, 1% and 2% CHG, applied on working days or alternate working days, with or without emollient to SOC. NeoCHG found a non-significant trend of greater efficacy with 1% CHG and no clear additional benefit with using 2% CHG.

Colony counts with OHP were higher than for CHG1% in the maternal stratum for total log_10_CFU, Gram-negative organisms, Gram-positive organisms, *Enterococcus* spp., *E. coli*, GBS, and *Staphylococcus aureus*. In the neonatal stratum, colony counts were higher for GBS only and there was no evidence of a difference between CHG1% and OHP for the remaining outcomes. Additionally, total log_10_CFU in neonates born to mothers in the maternal stratum was higher in the OHP arm compared to CHG1%. To our knowledge, this is the first *in vivo* comparison of the efficacy of OHP and CHG in reducing colonisation with potentially pathogenic organisms in these two patient populations. The findings suggest that OHP may be less efficacious for this purpose. The skin scores in the mothers treated with OHP were on average 0.26 points higher (worse) than in the SOC arm (95% CI: 0.07, 0.45; score range 0-16) although the clinical significance of this is unclear.

In the neonatal stratum, the benefits of multiple applications increased over time, which may reflect the rapid and consistent (re)colonisation of hospitalised neonates.^29^ Multiple applications may benefit neonates, a finding supported by NeoCHG, which found a non-significant trend of greater efficacy with higher application frequency.^25^ No robust evidence was observed in favour of different application frequencies in the maternal stratum, thus a single application may be sufficient. However, lower maternal sample sizes at later timepoints may mean that a true effect was not detectable. The timepoints in each stratum were selected to reflect the feasibility of application frequency for each patient group and to measure the antiseptic effects over time, and so the results may be more reflective of colonisation risk than the instantaneous effect of antiseptic application.

There was no evidence that SAE frequency differed between the trial arms in either stratum, or of a risk of post-antiseptic neonatal hypothermia, indicating that CHG1% is safe for future trials. These findings are consistent with NeoCHG which included low and very low birthweight neonates and found no safety signals with the use of CHG.^25^

Most research on antiseptic use during labour has focused on pre-C-section vaginal cleansing, with meta-analytical evidence indicating a benefit in infection reduction;^30^ this practice is recommended by the WHO. However, vaginal cleansing is not performed before vaginal births due to insufficient evidence of benefit; high-quality evidence is needed prior to making a recommendation given the implications for mothers of an additional vaginal procedure.^14^ This, therefore, remains a critical question, particularly in LICs, where infectious disease burden is greatest, and Gram-negative organisms predominate.^31,32^ This study demonstrated that vaginal cleansing during labour may reduce vaginal tract bacterial density so could impact maternal infection rates. An RCT powered to clinical endpoints could investigate this further.

Reducing bacterial load in the vaginal tract may benefit neonates by lowering the density of pathogenic bacteria, resulting in a decrease in vertical transmission. A South African trial conducted in 2009, that recruited over 8,000 women,^10^ found no difference in neonatal EOS rates following vaginal and neonatal cleansing with CHG versus control. However, the prevalence of Gram-negative neonatal bloodstream infections has increased substantially since^33^ and consequently vaginal cleansing may have additional benefits to neonates now. Although the use of CHG vaginal cleansing in labouring women has been deemed safe for their babies in this study and others,^28^ potential risks to neonates include altered skin- microbiome which should be balanced against infection risk.

Antiseptic cleansing of neonates could lead to a reduction in bloodstream infections and neonatal sepsis. However, causal pathways to infection are less clear than in mothers. Colonisation with pathogenic bacteria can be from the neonatal unit environment, such as cots and healthcare workers’ hands, ^34^ with *Klebsiella* spp. being a particular issue in African and Asian populations. Antiseptic cleansing may be most effective as part of a multimodal infection prevention strategy targeting healthcare-associated sepsis in neonates.^22,29^ In community-born neonates, CHG application reduced infection risk in LMICs, and a single CHG body wash lowered mortality in low-birthweight neonates.^35^ Evidence suggests CHG cleansing may benefit hospitalised neonates in LMICs. A Zambian study reported fewer culture-positive bloodstream infections with cleansing but no difference in culture-negative sepsis or mortality.^36^ An Indian trial showed a non-significant trend towards fewer Gram-positive bloodstream infections with CHG versus water.^37^ In Zambia, weekly CHG cleansing as part of a bundle reduced hospital mortality from 24% to 18%.^38^ CHG cleansing is widely used in NICUs globally, and a large trial evaluating its safety, efficacy, blood levels, and effect on the microbiome in high-risk neonatal populations is warranted.

Limitations of this trial include that it was conducted at a single centre, which could limit its generalisability. The trial was powered on bacterial load, including pathogenic and non-pathogenic organisms, which may not correlate with clinical outcomes. The reduced availability of data for later timepoints, due to mothers delivering or neonates being discharged, means that the information on antiseptic application frequency is less comprehensive; a larger sample size could have potentially combatted this. The skin scores used may be limited in their sensitivity to identify skin reactions.^39^

Strengths of the study include that neonates were recruited within the first 24 hours of life allowing assessment of the use of antiseptics in this early stage of colonisation. The efficient generation of data was enabled by the factorial trial design, which facilitated comparisons between two antiseptics (OHP and CHG), two concentrations of CHG (1% and 2%), and multiple antiseptic application frequencies against SOC within a single study. The trial incorporated maternal and neonatal strata, providing data on the impact of antiseptic use on bacterial colonisation of the maternal birth canal as well as directly on the neonatal skin.

In summary, this is the first trial to compare OHP and CHG in labouring women and neonates. A single application of CHG1% was beneficial in reducing the acquisition of potentially pathogenic bacteria in both the vaginal canal and on neonatal skin. No safety concerns were identified, supporting the use of CHG1% in a larger multifactorial infection prevention trial focused on clinical outcomes.

## Data Availability

Sharing of data will be considered based on a detailed proposal which should include aims, methods, and a statistical analysis plan. Requests will be checked for compatibility with regulatory and ethics committee requirements and with participant informed consent. Proposals should be addressed to Dr Louise F Hill and will be evaluated by the Sponsor.

## Acknowledgments

We wish to thank all the mothers, parents/guardians and children who participated in NeoVT-AMR as well as the wider research and clinical teams who contributed to the delivery of the trial. We are grateful to the members of the Data Monitoring Committee, Prof Elizabeth Molyneux, Dr Andrew Atkinson, and Prof Pisake Lumbiganon, for their time and valuable contribution. The chlorhexidine for NeoVT-AMR was kindly donated by Pharmanova, Blantyre, Malawi.

## Contributors

NeoVT-AMR was conceptualised by MS, JAB, AB, MNC, and ASW. The protocol and trial were designed by EB, MS, MNC, DL, NAF, JAB, and AB. NeoVT-AMR was conducted in Malawi by EB, MB, DL, NAF, and SL. The clinical trial was overseen by the Clinical Research Support Unit at the Malawi Liverpool Wellcome Trust, EB, LFH, DL, and MS. Microbiological analyses were overseen by NAF. MNC conducted the statistical analysis. MNC, EB, and LFH wrote the first draft of the paper. All authors contributed to the subsequent drafts, read, and approved the final version of the manuscript. MNC and ASW had full access to, and verified, all the data in the trial. All authors had final responsibility for the decision to submit for publication.

## References

1. Seale AC, Blencowe H, Manu AA, et al. Estimates of possible severe bacterial infection in neonates in sub-Saharan Africa, south Asia, and Latin America for 2012: a systematic review and meta-analysis. Lancet Infect Dis. 2014;14(8):731–741. doi:10.1016/S1473-3099(14)70804-7

2. Russell NJ, Stöhr W, Plakkal N, et al. Patterns of antibiotic use, pathogens, and prediction of mortality in hospitalized neonates and young infants with sepsis: A global neonatal sepsis observational cohort study (NeoOBS). PLoS Med. 2023;20(6):e1004179. doi:10.1371/journal.pmed.1004179

3. Investigators of the Delhi Neonatal Infection Study (DeNIS) collaboration. Characterisation and antimicrobial resistance of sepsis pathogens in neonates born in tertiary care centres in Delhi, India: a cohort study. Lancet Glob Health. 2016;4(10):e752-60. doi:10.1016/S2214-109X(16)30148-6

4. Mgusha Y, Nkhoma DB, Chiume M, et al. Admissions to a Low-Resource Neonatal Unit in Malawi Using a Mobile App and Dashboard: A 1-Year Digital Perinatal Outcome Audit. Front Digit Health. 2021;3:761128. doi:10.3389/fdgth.2021.761128

5. Wolf MF, Shqara RA, Naskovica K, et al. Vertical transmission of extended-spectrum, beta-lactamase-producing enterobacteriaceae during preterm delivery: A prospective study. Microorganisms. 2021;9(3):1–14. doi:10.3390/microorganisms9030506

6. Johnson J, Suwantarat N, Colantuoni E, et al. The impact of chlorhexidine gluconate bathing on skin bacterial burden of neonates admitted to the Neonatal Intensive Care Unit. Journal of Perinatology. 2019;39(1):63–71. doi:10.1038/s41372-018-0231-7

7. Simon A, Exner M. Prävention nosokomialer Infektionen bei intensivmedizinisch behandelten Frühgeborenen. Monatsschrift Kinderheilkunde. 2014;162(5):403–410. doi:10.1007/s00112-013-2974-8

8. Biermann C, Kribs A, Roth B, Tantcheva-Poor I. Use and cutaneous side effects of skin antiseptics in extremely low birth weight infants – A retrospective survey of the German NICUs. Klin Padiatr. 2016;228(04):208–212. doi:10.1055/s-0042-104122

9. Novakov Mikić A, Stojic S. Study results on the use of different therapies for the treatment of vaginitis in hospitalised pregnant women. Arch Gynecol Obstet. 2015;292(2):371–376. doi:10.1007/s00404-015-3638-9

10. Cutland CL, Madhi SA, Zell ER, et al. Chlorhexidine maternal-vaginal and neonate body wipes in sepsis and vertical transmission of pathogenic bacteria in South Africa: a randomised, controlled trial. Lancet. 2009;374(9705):1909-1916. doi:10.1016/S0140-6736(09)61339-8

11. Mullany LC, Biggar RJ. Vaginal and neonatal skin cleansing with chlorhexidine. Lancet. 2009;374(9705):1873-1875. doi:10.1016/S0140-6736(09)61593-2

12. Taha T, Biggar R, Broadhead R, al. et. Effect of cleansing the birth canal with antiseptic solution on maternal and newborn morbidity and mortality in Malawi: clinical trial. BMJ. 1997;315:216–219. doi:10.1136/bmj.315.7102.216

13. Bakr A, Karkour T. Effect of predelivery vaginal antisepsis on maternal and neonatal morbidity and mortality in Egypt. Journal of Womens Health (Larchmt). 2005;14:496–501. doi:10.1089/jwh.2005.14.496

14. Bell C, Hughes L, Akister T, Ramkhelawon V, Wilson A, Lissauer D. What is the result of vaginal cleansing with chlorhexidine during labour on maternal and neonatal infections? A systematic review of randomised trials with meta-analysis. BMC Pregnancy Childbirth. 2018;18(1):139. doi:10.1186/s12884-018-1754-9

15. Ohlsson A, Shah VS, Stade BC. Vaginal chlorhexidine during labour to prevent early- onset neonatal Group B streptococcal infection. Cochrane Database of Systematic Reviews. 2014;(12):CD003520. doi:10.1002/14651858.CD003520.pub3

16. Bonet M, Brizuela V, Abalos E, et al. Frequency and management of maternal infection in health facilities in 52 countries (GLOSS): a 1-week inception cohort study. Lancet Glob Health. 2020;8:e661–e671. doi:10.1016/S2214-109X(20)30109-1

17. Ogah CO, Anikwe CC, Ajah LO, et al. Preoperative vaginal cleansing with chlorhexidine solution in preventing post-cesarean section infections in a low resource setting: A randomized controlled trial. Acta Obstet Gynecol Scand. 2021;100(4):694–703. doi:10.1111/aogs.14060

18. Nnagbo EJ, Obi NS, Umeh AU. Effectiveness of Chlorhexidine vaginal cleansing in reducing post-caesarean endometritis at two tertiary hospitals in Enugu, Nigeria: A randomized controlled trial. Trop Doct. 2023;53(1):50–56. doi:10.1177/00494755221130561

19. World Health Organization. WHO recommendations for prevention and treatment of maternal peripartum infections. 2015. Accessed October 7, 2024. extension://efaidnbmnnnibpcajpcglclefindmkaj/https://iris.who.int/bitstream/handle/10665/186171/9789241549363_eng.pdf?sequence=1

20. Sankar MJ, Paul VK, Kapil A, et al. Does skin cleansing with chlorhexidine affect skin condition, temperature and colonization in hospitalized preterm low birth weight infants?: a randomized clinical trial. Journal of Perinatology. 2009;29(12):795–801. doi:10.1038/jp.2009.110

21. Clements MN, White IR, Copas AJ, et al. Improving Clinical Trial Interpretation with ACCEPT Analyses. NEJM Evidence. 2022;1(8):evidctw2200018. doi:10.1056/evidctw2200018

22. Heinz E, Pearse O, Zuza A, et al. Longitudinal analysis within one hospital in sub- Saharan Africa over 20 years reveals repeated replacements of dominant clones of Klebsiella pneumoniae and stresses the importance to include temporal patterns for vaccine design considerations. Genome Med. 2024;16(1). doi:10.1186/s13073-024-01342-3

23. Mullany LC, Khatry SK, Sherchand JB, et al. A randomized controlled trial of the impact of chlorhexidine skin cleansing on bacterial colonization of hospital-born infants in Nepal. Pediatric Infectious Disease Journal. 2008;27(6):505–511. doi:10.1097/INF.0b013e31816791a2

24. Darmstadt GL, Hossain MM, Choi Y, et al. Safety and effect of chlorhexidine skin cleansing on skin flora of neonates in Bangladesh. Pediatric Infectious Disease Journal. 2007;26(6):492–495. doi:10.1097/01.inf.0000261927.90189.88

25. Russell N, Clements MN, Azmery KS, et al. Safety and efficacy of whole-body chlorhexidine gluconate cleansing with or without emollient in hospitalised neonates (NeoCHG): a multicentre, randomised, open-label, factorial pilot trial. EClinicalMedicine. 2024;69:102463. doi:10.1016/j.eclinm.2024.102463

26. Anitha M, Sundaram MB, Kumutha J. Efficacy and safety of multiple whole-body cleansing with aqueous 0.5% chlorhexidine in stable NICU preterm infants - Randomized controlled trial. Journal of Neonatology. 2022;36(3):216–226. doi:10.1177/09732179221113684

27. Dramowski A, Pillay S, Bekker A, et al. Impact of 1% chlorhexidine gluconate bathing and emollient application on bacterial pathogen colonization dynamics in hospitalized preterm neonates – A pilot clinical trial. EClinicalMedicine. 2021;37. doi:10.1016/j.eclinm.2021.100946

28. Pereira L, Chipato T, Mashu A, et al. Randomized study of vaginal and neonatal cleansing with 1% chlorhexidine. International Journal of Gynecology and Obstetrics. 2011;112(3):234–238. doi:10.1016/j.ijgo.2010.09.009

29. Carvalho MJ, Sands K, Thomson K, et al. Antibiotic resistance genes in the gut microbiota of mothers and linked neonates with or without sepsis from low- and middle-income countries. Nat Microbiol. 2022;7(9):1337–1347. doi:10.1038/s41564-022-01184-y

30. Fadlalmola HA, Al-Sayaghi KM, Al-Hebshi AA, et al. Vaginal preparation with different antiseptic solutions before cesarean section for preventing postoperative infections: A systematic review and network meta-analysis. J Obstet Gynaecol Res. 2022;48(11):2659–2676. doi:10.1111/JOG.15377

31. Admas A, Gelaw B, Belaytessema, Worku A, Melese A. Proportion of bacterial isolates, their antimicrobial susceptibility profile and factors associated with puerperal sepsis among post-partum/aborted women at a referral Hospital in Bahir Dar, Northwest Ethiopia. Antimicrob Resist Infect Control. 2020;9(1):1–10. doi:10.1186/S13756-019-0676-2/TABLES/5

32. Van Dillen J, Zwart J, Schutte J, Van Roosmalen J. Maternal sepsis: Epidemiology, etiology and outcome. Curr Opin Infect Dis. 2010;23(3):249–254. doi:10.1097/QCO.0B013E328339257C

33. Thomas R, Ondongo-Ezhet C, Motsoaledi N, Sharland M, Clements M, Velaphi S. Incidence, pathogens and antimicrobial resistance of blood and cerebrospinal fluid isolates from a tertiary neonatal unit in South Africa: A 10 year retrospective review. PLoS One. 2024;19(1):e0297371. doi:10.1371/journal.pone.0297371

34. Pearse O. *Transmission of Extended-Spectrum Beta-Lactamase Klebsiella Pneumoniae on the Chatinkha Neonatal Unit, Blantyre, Malawi* . Liverpool School of Tropical Medicine; 2023. Accessed November 20, 2024. extension://efaidnbmnnnibpcajpcglclefindmkaj/https://archive.lstmed.ac.uk/23987/1/O%20Pearse_thesis_corrections_aug23_clean_v2.pdf

35. Tielsch JM, Darmstadt GL, Mullany LC, et al. Impact of Newborn Skin-Cleansing With Chlorhexidine on Neonatal Mortality in Southern Nepal: A Community-Based, Cluster- Randomized Trial.; 2008. doi:10.1542/peds.2006-1192

36. Westling T, Cowden C, Mwananyanda L, et al. Impact of chlorhexidine baths on suspected sepsis and bloodstream infections in hospitalized neonates in Zambia. Int J Infect Dis. 2020;96:54–60. doi:10.1016/J.IJID.2020.03.043

37. Gupta B, Vaswani N Das, Sharma D, Chaudhary U, Lekhwani S. Evaluation of efficacy of skin cleansing with chlorhexidine in prevention of neonatal nosocomial sepsis - a randomized controlled trial. J Matern Fetal Neonatal Med. 2016;29(2):242–247. doi:10.3109/14767058.2014.996126

38. Mwananyanda L, Pierre C, Mwansa J, et al. Preventing bloodstream infections and death in Zambian neonates: Impact of a low-cost infection control bundle. Clinical Infectious Diseases. 2019;69(8):1360–1367. doi:10.1093/cid/ciy1114

39. Darmstadt GL. Skin science to advance emollient therapy in the care and health of preterm infants. EClinicalMedicine. 2024;72:102618. doi:10.1016/j.eclinm.2024.102618

