## Supplement for "Safety and efficacy of antiseptic cleansing to reduce vertical transmission of multi-drug resistant pathogens to neonates (NeoVT-AMR)"

#### Supplementary material

1. **eFigure 1: Trial schema**

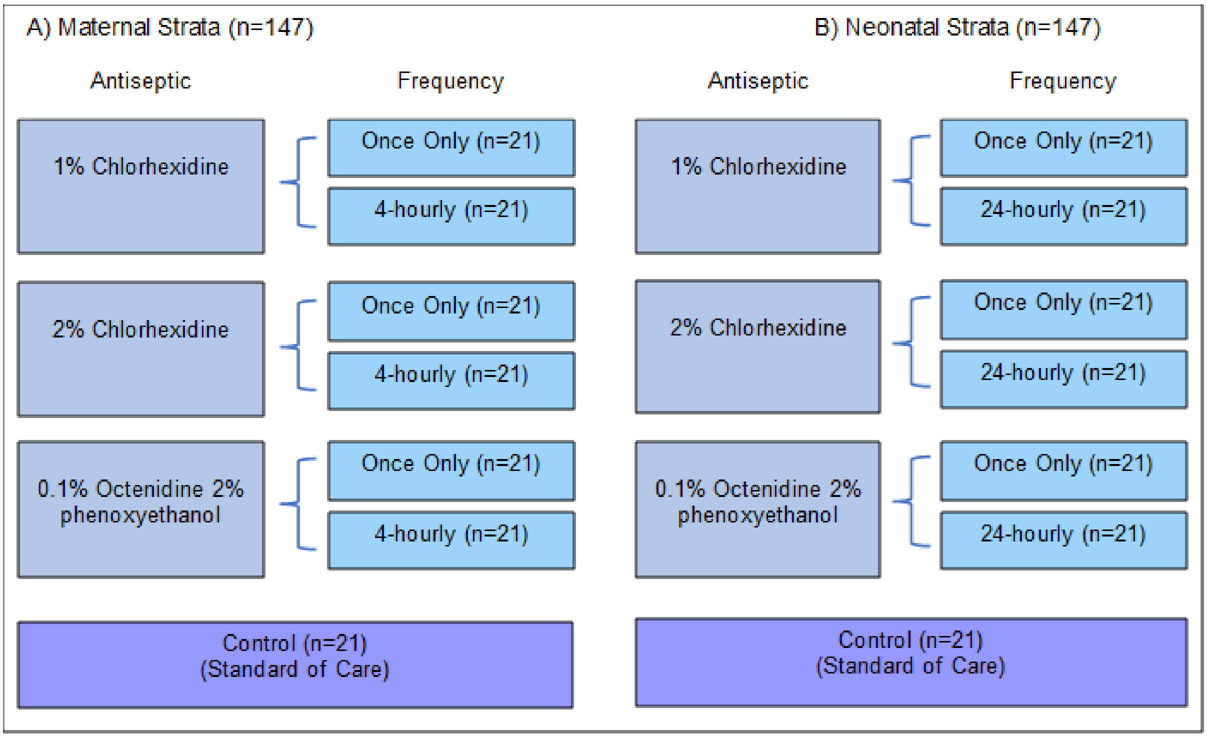

#### 2. eAppendix 1: Protocol amendments

Final protocol version was version 6.0. Recruitment commenced on version 3.0.

| Protocol versions | Summary of amendments | Comments |
|---|---|---|
| 3.0 to 5.0 | i) Inclusion criteria updated to allow recruitment of neonates born at any healthcare facility within the last 24 hours. Previous inclusion criterion only allowed recruitment of neonates born at Zomba Central Hospital within the last 12 hours.<br>ii) 28-day follow-up window increased from ±2 days to ±7 days. | There was no Protocol version 4.0. The College of Medicine Research & Ethics Committee (COMREC) requires version numbers of Protocols and corresponding Patient Information Sheets to be the same. Amendments were made to the Patient Information Sheet with no corresponding amendment to the Protocol and therefore, the protocol was updated to version 5.0 to correspond with the updates to the Patient Information Sheet. |
| 5.0 to 6.0 | The recruitment period was extended from January 2023 to March 2023. | |

#### 3. eMethods

##### 3.1 Skin score assessment criteria

##### Neonatal stratum skin score assessment criteria

Adapted from Lund C & Osborne JW.[1] The skin should be scored on each of the three domains (dryness, erythema and skin breakdown), and then the total score is the sum of the three components (minimum score 0, maximum score 12).

Highest grade from:

Grade 1 = Score of ≥1 in ≥2 categories

Grade 2 = Score of ≥2 in ≥2 categories

Grade 3 = Score of ≥3 in ≥2 categories

Grade 4 = Score of ≥4 in ≥2 categories

| Score | Description |
|---|---|
| Domain 1 | Dryness |
| 0 | Normal |
| 1 | Dry skin +/- scaling <50% |
| 2 | Dry skin + scaling >50% +/- cracking/fissures <50% |
| 3 | Very dry + cracking / fissures >50% |
| 4 | Very dry + cracking / fissures >75% |
| Domain 2 | Erythema |
| 0 | Normal |
| 1 | Erythema <25% |
| 2 | Erythema 25-50% |
| 3 | Severe Erythema 50-75% |
| 4 | Severe Erythema >75% |
| Domain 3 | Skin breakdown |
| 0 | Normal |
| 1 | Skin breakdown/ulceration/vesicles <5% |
| 2 | Skin breakdown/ulceration/vesicles 5-25% |
| 3 | Skin breakdown/ulceration/vesicles 25-50% |
| 4 | Skin breakdown/ulceration/vesicles >50% |

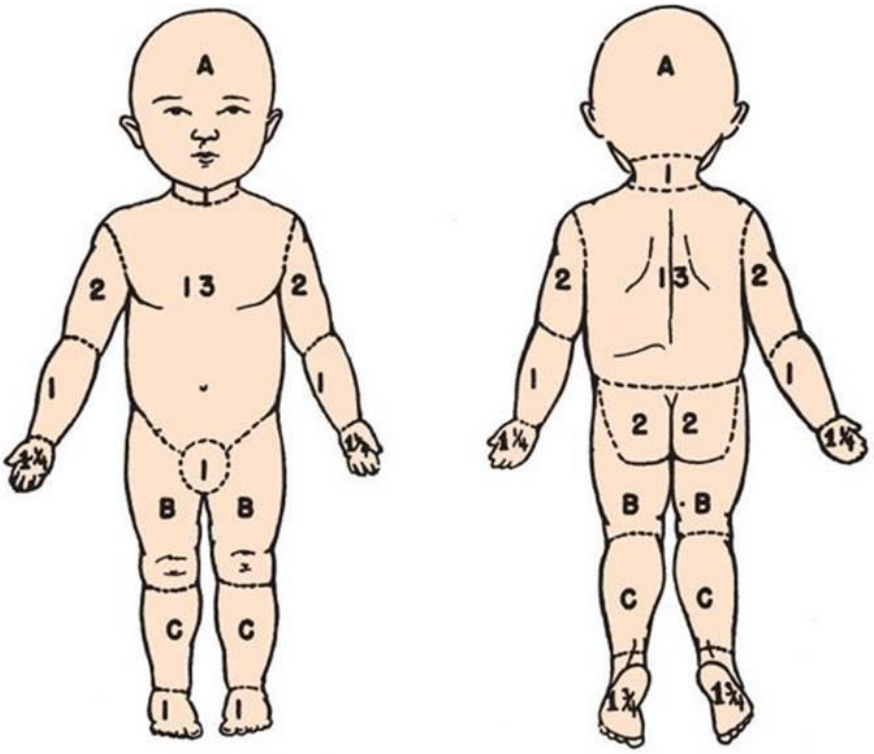

**RELATIVE PERCENTAGES OF AREAS AFFECTED BY GROWTH**

| Area | Age 0 | 1 | 5 |
|---|---|---|---|
| A = ½ of Head | 9½ | 8½ | 6½ |
| B = ½ of One Thigh | 2¾ | 3¼ | 4 |
| C = ½ of One Leg | 2½ | 2½ | 2¾ |

**% BURN BY AREAS**

Probable 3rd° Burn { Head____ Neck____ Body____ Up. Arm____ Forearm____ Hands____ Genitals____ Buttocks____ Thighs____ Legs____ Feet____

Total Burn { Head____ Neck____ Body____ Up. Arm____ Forearm____ Hands____ Genitals____ Buttocks____ Thighs____ Legs____ Feet____

Sum of All Areas________ Probably 3rd° ________ Total Burn________

**Figure demonstrating skin affected area assessment.**

Modified Lund and Browder chart for estimation of body surface area burn involvement in infants and children.[2]

##### Maternal stratum skin score assessment criteria

Adapted from NCI. National Cancer Institute Common Terminology Criteria for Adverse Events.[3]

The vaginal and vulval area should be scored on each of the four domains: 1 symptom (irritation) and 3 signs (erythema, skin breakdown, swelling). The total score is the sum of the three components (minimum score 0, maximum score 16).

Highest grade from:

Grade 1 = Score of ≥1 in ≥2 categories

Grade 2 = Score of ≥2 in ≥2 categories

Grade 3 = Score of ≥3 in ≥2 categories

Grade 4 = Score of ≥4 in ≥2 categories

**PART A – Question to labouring women:**

| 1. Vaginal/vulval irritation | | | | |
|---|---|---|---|---|
| What is the severity of your vaginal irritation now at its worst? | | | | |
| None = 0 | Mild = 1 | Moderate = 2 | Severe = 3 | Very severe = 4 |

**PART B – On inspection of external genitalia:**

| 2. Vaginal/vulval erythema | | | | |
|---|---|---|---|---|
| What is the severity of erythema? (select the most severe that is present) | | | | |
| None = 0 | Mild = 1 | Moderate = 2 | Severe = 3 | Very severe = 4 |

| 3. Vaginal/vulval skin breakdown | | | | |
|---|---|---|---|---|
| What is the severity of skin breakdown? (select the most severe that is present) | | | | |
| None = 0 | Mild = 1 | Moderate = 2 | Severe = 3 | Very severe = 4 |

| 4. Vaginal/vulval swelling | | | | |
|---|---|---|---|---|
| What is the severity of swelling? (select the most severe that is present) | | | | |
| None = 0 | Mild = 1 | Moderate = 2 | Severe = 3 | Very severe = 4 |

##### 3.2 6–swab technique for maternal antiseptic application

Using six cotton wool balls soaked in antiseptic (chlorhexidine (CHG)1%, CHG2% or (octenidine 0.1% combined with phenoxyethanol 2% (OHP)) or sterile water (SOC).

1. Swab first labia majora with a downward motion using 1 swab per stroke (extending down to perineum)
2. Swab second labia majora with a downward motion using 1 swab per stroke (extending down to perineum)
3. Swab first labia minora with a downward motion using 1 swab per stroke
4. Swab second labia minora with a downward motion using 1 swab per stroke
5. Swab rest of vulva and vaginal introitus using 1 swab
6. Insert cotton wool into vagina and cervical os, using circular motions cleanse from cervical os outwards to cleanse the inside of the vagina

##### 3.3 Collection of microbiological samples and laboratory procedures

###### Swabs and processing

Swab collection was only performed by the study nurses to ensure standardisation of sample collection. Dry flocked cotton swabs were used. In the maternal stratum, low vaginal and perineal swabs were taken at 0, 4, 8, 24, 28, 32 hours from randomisation; if heavily soiled, the area was cleansed with sterile water prior to swabbing (see table below). In the neonatal stratum, neck (cervical skin fold) and peri-rectal swabs were taken; no skin preparation was used (see 'colonisation sampling procedure' below). Swabs were placed in liquid amies transport medium (LTM). Samples were refrigerated for 24-48 hours prior to being processed in the laboratory. Swabs in LTM were vortexed, and ten-fold dilutions of the LTM with saline were performed, down to a dilution of $10^{-4}$. 20µL aliquots of the neat LTM were inoculated onto UriSelect agar (Bio Rad, Hercules, USA) and blood sheep agar (made in-house); and the $10^{-2}$ and $10^{-4}$ dilutions were inoculated onto UriSelect agar only. Agar plates were incubated overnight (18-20 hours) at 35°C in air.

The UriSelect and blood sheep agar plates with colonies most suitable for colony counts were selected for manual colony counting. Total number of colonies as well as colony counts for Gram-positive organisms, Gram-negative organisms and any identified species were also recorded. The colony counts were converted to $\log_{10}$CFU.

###### Colonisation sampling procedure

| Sampling site | Sampling method |
|---|---|
| **Maternal stratum** | |
| Low vaginal | Skin outside the vagina was gently spread with one hand. The swab tip was inserted into the vaginal introitus, and gently slid 3 cm into the vagina. The swab was gently rotated 360 degrees 10 times, taking 1 second for each turn. The swab tip was withdrawn without touching the skin. |
| Perineal | The perineum was wiped with cotton wool soaked in tap water when visibly soiled. Taking care to avoid other contact with the swab, the swab was rotated 360 degrees against the perineal skin (the area between the anus and external genitalia) for 5 seconds from one side to the next. This was then repeated one further time. |
| **Neonatal stratum** | |
| Neck (cervical skin fold) | The swab was applied between the neck folds whilst continuously rotating the tip backwards and forwards. The anterior 180-degree portion of the neck was swabbed. The process was repeated one further time. |
| Peri-rectal area | The swab was rotated 360 degrees around the peri-rectal area whilst continuously rotating the tip. This was then repeated one further time. |

###### Organism identification

Organisms were presumptively identified based on the colony colour on the UriSelect agar (Bio-Rad, Redmond, USA) with more precise identification undertaken, where relevant, using Gram staining as well as specific identification tests. Colour and indole tests were used to differentiate *Escherichia coli* from other Enterobacterales. Potential *Klebsiella* spp., *Serratia spp.* and *Enterobacter* spp. (purple-blue colonies), were identified using the Analytical Profile Index (API) (20E) (Biomerieux, Marcy-l'Etoile, France). Oxidase was adopted to differentiate Gram-negative bacilli (Pseudomonas spp. or Acinetobacter spp.). Catalase was used to differentiate Gram-positive cocci (*Staphylococcus* spp. or *Streptococcus* spp.). Coagulase was used on catalase positive organisms to differentiate *Staphylococcus aureus* from other *Staphylococcus* spp.. Catalase negative organisms were further differentiated based on the type of haemolysis on blood agar (α- and β-haemolytic streptococci) and Lancefield typing (β-haemolytic streptococci groups A or B) was conducted using the Streptococcal Latex Grouping Kit (Pro-Lab, Ontario, Canada).

###### Susceptibility testing

Susceptibility testing was performed on *Staphylococcus aureus, Streptococcus* spp., *Enterococcus* spp. and Enterobacterales and non-fermenting Gram-negative colonising organisms using the standard EUCAST disc diffusion method and interpreted using the EUCAST clinical breakpoints v14.0 or CLSI criteria (27th edition, 2017) where breakpoints were not available. Antibiotics tested against each species are outlined in the below table.

#### eTable 1: antibiotic susceptibility testing by identified colonising organisms

| Organism | AK 30 | AMP 2 | AMP 10 | C3 0 | CAZ 10 | CIP 5 | CN 10 | CPD 10 | CRO 30 | E1 5 | FOX 30 | MERO 10 | OXA 1 | P 1 | PE F5 | PTZ3 6 | QD 15 | TE30 | VA 5 |
|---|---|---|---|---|---|---|---|---|---|---|---|---|---|---|---|---|---|---|---|
| Enterobacterales | ■ | | | ■ | | ■ | ■ | ■ | | | | ■ | | | | | | | |
| *Acinetobacter* spp. | ■ | | | | | ■ | ■ | | | | | ■ | | | | | | | |
| B- haem streptococcus (A, B, C, G) | | | | ■ | | | | | | ■ | | | | ■ | | | | | ■ |
| *Enterococcus* spp. | | ■ | | ■ | | ■ | | | | | | | | | | | ■ | | ■ |
| *Staphylococcus aureus* | | | | ■ | | ■ | ■ | | | ■ | ■ | | | | | | | ■ | |

■ = antibiotic was tested against organism

*AK30 = amikacin 30µg; AMP2 = Ampicillin 2µg; C30 = Chloramphenicol 30µg; CAZ10 = Ceftazidime 10µg; CIP5 = Ciprofloxacin 5µg; CN10 = Gentamicin 10µg; CPD10 = Cefpodoxime 10µg; CRO30 = Ceftriaxone 30µg; E15 = Erythromycin 15µg; FOX30 = Cefoxitin 30µg; MERO10 = Meropenem 10µg; OXA1 = Oxacillin 1µg; P1 = Penicillin 1µg; PEF5 = Pefloxacin 5µg; PTZ36 = Piperacillin-tazobactam 36µg (30-6); QD15 = Quinupristin-dalfopristin 1 µg; TE30 = tetracycline 30µg VA5 = Vancomycin 5µg*

#### 3.4 Adverse event grading scales eTable 2 and eTable 3

### eTable 2: Maternal stratum adverse event grading scale

Adapted from Common Terminology Criteria for Adverse Events[3]

| Parameter | Grade 0 Normal | Grade 1 Mild | Grade 2 Moderate | Grade 3 Severe | Grade 4 Potentially Life-Threatening | Grade 5 |
|---|---|---|---|---|---|---|
| Blood and lymphatic system disorders | | | | | | |
| Anaemia | Haemoglobin (Hb) >10g/dL | Hb <10g/dL | Hb 8-10 g/dL | Hb ≤ 8 g/dL; transfusion indicated | Life threatening consequences; urgent intervention indicated | Death |
| Disseminated intravascular coagulation | – | – | Laboratory findings with no bleeding | Laboratory findings and bleeding | Life threatening consequences; urgent intervention indicated | Death |
| Hemolysis | – | Laboratory evidence of haemolysis only (e.g. direct antiglobulin test (DAT); Coombs; schistocytes; decreased haptoglobin) | Evidence of haemolysis and ≥2g decrease in haemoglobin | Transfusion or medical intervention indicated (e.g. steroids) | Life threatening consequences; urgent intervention indicated | Death |
| Blood and lymphatic system disorders – other | – | Asymptomatic or mild symptoms; clinical or diagnostic observations only; intervention not indicated | Moderate; minimal, local or non-invasive intervention indicated; limiting age-appropriate instrumental activities of daily living (ADL) | Severe or medically significant but not immediately life-threatening; hospitalization or prolongation of existing hospitalization indicated; limiting self-care ADL | Life-threatening consequences; urgent intervention indicated | Death |
| Cardiac disorders | | | | | | |
| Sinus tachycardia | – | Asymptomatic, intervention not indicated | Symptomatic, non-urgent medical intervention indicated | Urgent medical intervention indicated | – | - – |
| Supraventricular tachycardia | – | Asymptomatic, intervention not indicated | Non-urgent medical intervention indicated | Symptomatic, urgent intervention indicated | Life threatening consequences | Death |
| Cardiac disorders – other, specify | – | Asymptomatic or mild symptoms; clinical or | Moderate; minimal, local or non-invasive intervention | Severe or medically significant but not immediately life- | Life-threatening consequences; urgent intervention indicated | Death |

| Parameter | Grade 0 Normal | Grade 1 Mild | Grade 2 Moderate | Grade 3 Severe | Grade 4 Potentially Life-Threatening | Grade 5 |
|---|---|---|---|---|---|---|
| | | diagnostic observations only; intervention not indicated | indicated; limiting age-appropriate instrumental ADL | threatening; hospitalisation or prolongation of existing hospitalisation indicated; limiting self-care ADL | | |
| Gastrointestinal | | | | | | |
| Anal fistula | Asymptomatic | Symptomatic | Invasive intervention not indicated | Invasive intervention indicated | Life threatening consequences; urgent intervention indicated | – |
| Haemorrhoids | – | Asymptomatic; clinical or diagnostic observations only; intervention not indicated | Symptomatic, banding or medical intervention indicated | Severe symptoms, invasive intervention indicated | – | – |
| Haemorrhoidal haemorrhage | – | Mild symptoms; intervention not indicated | Moderate symptoms; intervention indicated | Transfusion indicated; invasive intervention indicated; hospitalisation | Life-threatening consequences; urgent intervention indicated | Death |
| Nausea | – | Loss of appetite without alteration in eating habits | Oral intake decreased without significant weight loss, dehydration or malnutrition | Inadequate oral caloric or fluid intake; tube feeding, TPN, or hospitalization indicated | – | – |
| Vomiting | – | Intervention not indicated | Outpatient intravenous (IV) hydration; medical intervention indicated | Tube feeding, total parenteral nutrition (TPN) or hospitalisation indicated | Life-threatening consequences | Death |
| Ileus | – | Asymptomatic and radiologic observations only | Symptomatic, altered GI function, bowel rest indicated | Severely altered GI function; TPN indicated, tube placement indicated | Life-threatening consequences; urgent intervention indicated | Death |
| General disorders | | | | | | |
| Neonatal death | – | – | – | – | Neonatal loss of life | – |
| Oedema of limbs | – | 5-10% inter-limb discrepancy in volume or circumference at point of greatest visible difference; | >10-30% inter-limb discrepancy in volume or circumference at point of greatest visible difference; readily apparent obscuration of anatomic | >30% inter-limb discrepancy in volume; gross deviation from normal anatomic contour; limiting self-care ADL | – | – |

| Parameter | Grade 0 Normal | Grade 1 Mild | Grade 2 Moderate | Grade 3 Severe | Grade 4 Potentially Life-Threatening | Grade 5 |
|---|---|---|---|---|---|---|
| | | swelling or obscuration of anatomic architecture on close inspection | architecture; obliteration of skin folds; readily apparent deviation from normal anatomic contour; limiting instrumental ADL | | | |
| Fever | – | 38.0–39.0°C | 39.0–40.0°C | >40.0°C for <24 hours | >40.0°C for >24 hours | Death |
| Immune system disorders | | | | | | |
| Allergic reaction | – | Systemic intervention not indicated | Oral intervention indicated | Bronchospasm; hospitalisation indicated for clinical sequelae; intravenous intervention indicated | Life-threatening consequences; urgent intervention indicated | Death |
| Anaphylaxis | – | – | – | Symptomatic bronchospasm, with or without urticaria; parenteral intervention indicated; allergy-related oedema/angioedema; hypotension | Life-threatening consequences; urgent intervention indicated | Death |
| Infections | | | | | | |
| Abdominal infection | – | – | Oral intervention indicated | IV antibiotic, antifungal or antiviral intervention indicated; invasive intervention indicated | Life-threatening consequences; urgent intervention indicated | Death |
| Bacteraemia | – | – | Blood culture positive with no signs or symptoms | | | |
| Cervicitis infection | – | – | Localised, local intervention indicated (e.g. topical antibiotic, antifungal or antiviral) | IV antibiotic, antifungal or antiviral intervention indicated; invasive intervention indicated | Life-threatening consequences; urgent intervention indicated | Death |
| Pelvic infection | – | – | Moderate symptoms, oral | IV antibiotic, antifungal or antiviral | Life-threatening consequences; | Death |

| Parameter | Grade 0 Normal | Grade 1 Mild | Grade 2 Moderate | Grade 3 Severe | Grade 4 Potentially Life-Threatening | Grade 5 |
|---|---|---|---|---|---|---|
| | | | intervention indicated | intervention indicated; invasive intervention indicated | urgent intervention indicated | |
| Sepsis | – | – | – | Blood culture positive with signs or symptoms; treatment indicated | Life threatening consequences; urgent intervention indicated | Death |
| Thrush | – | Asymptomatic; local symptomatic management | Oral intervention indicated | IV antifungal intervention indicated | – | – |
| Urinary tract infection | – | – | Localised oral intervention indicated | IV antibiotic, antifungal or antiviral intervention indicated; invasive intervention indicated | Life threatening consequences; urgent intervention indicated | Death |
| Uterine infection | – | – | Moderate symptoms; oral intervention indicated | IV antibiotic, antifungal or antiviral intervention indicated; invasive intervention indicated | Life threatening consequences; urgent intervention indicated | Death |
| Vaginal infection | – | Localised, local intervention indicated | Oral intervention indicated | IV antibiotic, antifungal or antiviral intervention indicated; invasive intervention indicated | Life threatening consequences; urgent intervention indicated | Death |
| Vulval infection | – | Localised, local intervention indicated | Oral intervention indicated | IV antibiotic, antifungal or antiviral intervention indicated; invasive intervention indicated | Life threatening consequences; urgent intervention indicated | Death |
| Wound infection | – | Localised, local intervention indicated | Oral intervention indicated | IV antibiotic, antifungal or antiviral intervention indicated; invasive intervention indicated | Life threatening consequences; urgent intervention indicated | Death |
| Injury and procedural complications | | | | | | |

| Parameter | Grade 0 Normal | Grade 1 Mild | Grade 2 Moderate | Grade 3 Severe | Grade 4 Potentially Life-Threatening | Grade 5 |
|---|---|---|---|---|---|---|
| Intraoperative arterial injury; intraoperative gastrointestinal injury; intraoperative reproductive tract injury; urinary injury | – | Primary repair of injured organ/structure indicated | Partial resection of injured organ/structure indicated | Complete resection or reconstruction of injured organ/structure indicated; limiting self-care ADL | Life-threatening consequences; urgent intervention indicated | Death |
| Intraoperative haemorrhage | – | – | – | Postoperative invasive intervention indicated, hospitalisation | Life-threatening consequences; urgent intervention indicated | Death |
| Postoperative haemorrhage | – | Mild symptoms; intervention not indicated | Moderate bleeding requiring transfusion <2 units | Transfusion indicated ≥2 units; invasive intervention indicated; hospitalisation | Life-threatening consequences, urgent intervention indicated | Death |
| Uterine perforation | – | | Invasive intervention not indicated | Invasive intervention indicated | Life-threatening consequences, urgent intervention indicated | Death |
| Wound complication | – | Observation only, topical intervention indicated | Bedside local care indicated | Operative intervention indicated | Life-threatening consequences | Death |
| Wound dehiscence | – | Incisional separation, intervention not indicated | Incisional separation, local care (e.g. suturing) or medical intervention indicated (e.g. analgesic) | Fascial disruption or dehiscence without evisceration; revision by operative intervention indicated | Life-threatening consequences; symptomatic hernia with evidence of strangulation; fascial disruption with evisceration; major reconstruction flap, grafting, resection | Death |
| Pregnancy, puerperium and perinatal conditions | | | | | | |
| Fetal growth retardation | – | – | <10% percentile of weight for gestational age | <5% percentile of weight for gestational age | <1% percentile of weight for gestational age | – |
| Pregnancy loss | – | – | – | – | Fetal loss at any gestational age | – |
| Premature delivery | – | Delivery of a liveborn infant at >34 to 37 weeks gestation | Delivery of a liveborn infant at >28 to 34 weeks gestation | Delivery of a liveborn infant at 24 to 28 weeks gestation | Delivery of a liveborn infant at 24 weeks of gestation or less | – |

| Parameter | Grade 0 Normal | Grade 1 Mild | Grade 2 Moderate | Grade 3 Severe | Grade 4 Potentially Life-Threatening | Grade 5 |
|---|---|---|---|---|---|---|
| Renal and urinary disorders | | | | | | |
| Acute kidney injury | – | – | – | Hospitalisation indicated | Life-threatening consequences; dialysis indicated | Death |
| Reproductive system | | | | | | |
| Vaginal discharge | Normal for patient | Mild vaginal discharge (greater than baseline for patient) | Moderate to heavy vaginal discharge; use of perineal pad or tampon indicated | – | – | – |
| Uterine haemorrhage | No haemorrhage | Mild symptoms; intervention not indicated | Moderate symptoms; intervention indicated | Transfusion indicated; invasive intervention indicated; hospitalisation | Life threatening consequences; urgent intervention indicated | Death |
| Vaginal haemorrhage | – | Mild symptoms; intervention not indicated | Moderate symptoms; intervention indicated | Transfusion indicated; invasive intervention indicated; hospitalisation | Life threatening consequences; urgent intervention indicated | Death |
| Vaginal perforation | – | – | Invasive intervention not indicated | Invasive intervention indicated | Life threatening consequences; urgent intervention indicated | Death |
| Respiratory | | | | | | |
| Hypoxia | – | – | Decreased oxygen saturation with exercise (e.g. pulse oximeter <88%); intermittent supplemental oxygen | Decreased oxygen saturation at rest (e.g. pulse oximeter <88%) | Life threatening compromise; urgent intervention needed | Death |
| Vascular | | | | | | |
| Thromboembolic event | – | Medical intervention not indicated (e.g. superficial thrombosis) | Medical intervention indicated | Urgent medical intervention indicated (e.g. pulmonary embolism) | Life-threatening consequences with haemodynamic or neurologic instability | Death |

#### eTable 3: Neonatal stratum adverse event grading scale

Adapted DAIDS score for neonates reproduced from the IMPAACT study[4]

| Parameter | Grade 0<br>Normal | Grade 1<br>Mild | Grade 2<br>Moderate | Grade 3<br>Severe | Grade 4<br>Potentially Life-Threatening |
|---|---|---|---|---|---|
| Apnoea*<br><br>*Apnoea spell defined as apnoea event >20s or associated with bradycardia, hypoxia or cyanosis | No apnoeas | <6 spells* per day | 6<12 spells* per day or nasal cannula for apnoea | 12 or more spells* per day or nasal continuous positive airway pressure (NCPAP) for apnoea | Requires intubation for apnoea |
| Anaemia | Haemoglobin (Hb) >10 g/dL | Hb 8 – 10 g/dL | Hb ≤8 g/dL | Requires packed red cell transfusion, no clinical signs | Requires packed red cell transfusion, clinical signs of shock |
| Congenital anomalies | None | Minor (no impairment of function) | Minor (no impairment of function), future treatment may be needed | Major (impairment of function), no immediate treatment needed | Major (impairment of function), immediate treatment needed |
| Congenital heart disease | No congenital heart disease | Minor (no impairment of function), no treatment needed | Minor (no impairment of function), future treatment may be needed | Major (impairment of function), no immediate treatment needed | Major (impairment of function), immediate treatment needed |
| Electrolyte/ Metabolic disorders | None | ------------------ | Electrolyte/Metabolic disorder, no systemic signs | ---------------------- | Electrolyte/Metabolic disorder with systemic signs |
| Gastrointestinal dysfunction [including necrotising enterocolitis (NEC)] | None | Not ill, abnormal abdominal exam, no treatment needed (e.g. abdominal distension but not NPO) | Mild illness, nil per os (NPO) <3 days (e.g. rule out NEC evaluation, Stage I NEC) | Moderate illness, NPO 3 – 7 days, or medical treatment (e.g. Stage II NEC) | Severe illness, NPO >7 days, or surgical treatment needed (e.g. Stage III NEC) |
| Hypertension | No blood pressure (BP) performed | Systolic BP >80–100, no treatment | Systemic BP >100, no treatment | Treated with one agent | Treatment with multiple agents |
| Hypotension | No BP performed | Mild clinical signs, no treatment needed | Symptomatic, treated with | Symptomatic, treated with | Clinical signs of shock or requiring use of |

| Parameter | Grade 0 Normal | Grade 1 Mild | Grade 2 Moderate | Grade 3 Severe | Grade 4 Potentially Life-Threatening |
|---|---|---|---|---|---|
| | | | intravenous fluids | single medication | multiple medications |
| Intraventricular haemorrhage | No abnormality noted or not assessed | Germinal matrix haemorrhage | Blood in ventricle, no enlargement | Blood in ventricle, with ventricular enlargement | Parenchymal haemorrhage and/or need for ventricular drainage |
| Jaundice | No jaundice | Mild jaundice, no treatment | Phototherapy and/or intravenous immunoglobulin | Exchange transfusion | Acute bilirubin encephalopathy |
| Neonatal abstinence syndrome (NAS) | No history of abstinence syndrome | NAS signs, no medical treatment | NAS controlled with single drug | NAS controlled with two drugs | NAS with seizures |
| Neurologic compromise | Normal neurological examinations | Mildly abnormal neurologic exam, no clinical or electroencephalogram (EEG) seizure activity | Stage I encephalopathy or possible clinical or EEG seizure activity but no treatment | Stage II encephalopathy or single drug seizure therapy | Stage III encephalopathy or multiple drug seizure therapy |
| Neutropenia | Absolute neutrophil count (ANC) >1000/mm$^3$ | ANC <1000/mm$^3$ | ANC <500/mm$^3$ | Treated with granulocyte colony-stimulating factor | White blood cell transfusion |
| Patent ductus arteriosus | No murmur | Clinical signs, no treatment | Treatment with fluid restriction or diuretics | Treatment with indomethacin or ibuprofen | Surgical ligation |
| Persistent pulmonary hypertension | No difference in pre- and post-ductal difference | Supplemental $O_2$ but no mechanical ventilation | Conventional ventilator <5 days | Conventional ventilation 5 – 10 days, alternative ventilation (e.g. high frequency oscillatory ventilation (HFOV), sildenafil and/or nitric oxide | Mechanical ventilation >10 days |
| Renal dysfunction | Wet diapers documented | Urine output 1 < 1.5 mL/kg/hr | Urine output 0.5 < 1.0 mL/kg/hr | Urine output 0 < 0.5 mL/kg/hr | Prolonged anuria |
| Respiratory Insufficiency | Baby is in room air – no | Nasal cannula oxygen with $FiO_2$ <0.5 | Continuous positive airway pressure | Conventional ventilation | Alternative ventilation (e.g. HFOV) |

| Parameter | Grade 0 Normal | Grade 1 Mild | Grade 2 Moderate | Grade 3 Severe | Grade 4 Potentially Life-Threatening |
|---|---|---|---|---|---|
| | supplemental oxygen | | (CPAP) or nasal cannula with $FiO_2$ >0.5 | | |
| Retinopathy of prematurity (ROP) | Normal vascularisation or not assessed | Incomplete vascularisation | Pre-threshold ROP | Threshold ROP or ROP treatment | Retinal detachment |
| Sepsis | No sepsis symptoms and signs | Septic evaluation, no treatment | Rule out sepsis, antibiotics for ≤72 hours | A confirmed blood stream infection with a course of antibiotic treatment, no septic shock or meningitis | A confirmed blood stream infection with a course of antibiotic treatment with septic shock or meningitis |
| Thrombocytopaenia | >100,000 cells/$mm^3$ | 75,000-100,000 cells/$mm^3$ | 50,000 < 75,000 cells/$mm^3$ | 25,000<50,000 cells/$mm^3$ | <25,000 cells/$mm^3$ or platelet transfusion |

##### 3.5 Priors used in Bayesian ACCEPT analyses

###### Non-informative priors

Non informative priors were selected to capture a wide range of possible values. The same priors will be used for the maternal and the neonatal stratum.

- Intercept mean =4, SD = 50
- Difference mean =0, SD = 20

###### Informative priors (optimistic and sceptical)

Informative priors were determined based on NeoCHG data (unpublished) and expert clinician opinion (Nicholas Feasey, David Lissauer, Emily Beales, Louise Hill, Neal Russell) during an online meeting on 21/03/2023. There is a lack of robust data to support the use of strong priors, especially in the maternal stratum.

###### Intercept specification

The intercept (SOC) was selected as data from NeoCHG could be used. The maternal stratum will use data from d8 in NeoCHG, reflecting bacterial load with time to accumulate, whereas the neonatal stratum will use data from baseline reflecting the recent birth of the neonates. Using mean values from the study and approximations of SD with a multiplier on the SD to capture extra uncertainty, the intercepts will be specified as following a normal distribution with parameters:

- $\log_{10}$CFU in maternal stratum: mean 8.4; standard deviation 2.6; multiplier 3
- $\log_{10}$CFU in neonatal stratum: mean 4.5; standard deviation 3; multiplier 2

A higher multiplier was chosen for the maternal stratum as there is greater uncertainty in the expected values. Priors may be reformulated to a reference of 1%CHG/once before analysis to match reference specification in focal analysis.

###### Difference specification

Clinicians specified the mean difference that optimistic and sceptical persons knowledgeable on the topic might assume, using a reference of no antiseptic application. The approach taken was that both optimistic and sceptical persons were assumed to have some belief that the opposing view could be correct, given the assumption of equipoise for the trial to be able to go ahead. The mean values selected were:

###### Maternal stratum

| Parameter | Optimistic mean | Sceptical mean |
|---|---|---|
| 0.5% CHG | Lower by 0.5 | No effect |
| 2% CHG | Lower by 1 | No effect |
| OHP | Lower by 1 | No effect |
| Single application | Lower by 0.5 | No effect |
| Multiple application | Lower by 1 | No effect |

*CHG = chlorhexidine; OHP = octenidine 0.1% combined with phenoxyethanol 2%*

###### Neonatal stratum

| Parameter | Optimistic mean | Sceptical mean |
|---|---|---|
| 0.5% CHG | Lower by 0.5 | No effect |
| 2% CHG | Lower by 1 | No effect |
| Emollient | Lower by 1 | No effect |
| Alternate day application | Lower by 0.5 | No effect |
| SOC | Lower by 1 | No effect |

*CHG = chlorhexidine; OHP = octenidine 0.1% combined with phenoxyethanol 2%*

Larger differences were selected than those found in NeoCHG due to the differing site conditions in NeoVT-AMR.

Differences were specified to follow a Normal distribution with mean as above and a certain percentage of the distribution more extreme than the opposing value (sceptical for optimistic priors and optimistic for sceptical priors) reflecting probability of alternate scenario being correct. This equates to a Normal distribution with:

- Mean = optimistic or sceptical mean

- SD = abs(sceptical mean – optimistic mean)/critical_value

Where the critical value is from the t-distribution with infinite degrees of freedom and probability equal to the alternate scenario being correct.

The probability of the alternate scenario being correct was:

- Maternal stratum: 0.2
- Neonatal stratum: 0.1

A larger probability was chosen for the maternal stratum to reflect the greater uncertainty in this stratum.

Plots of the differences are:

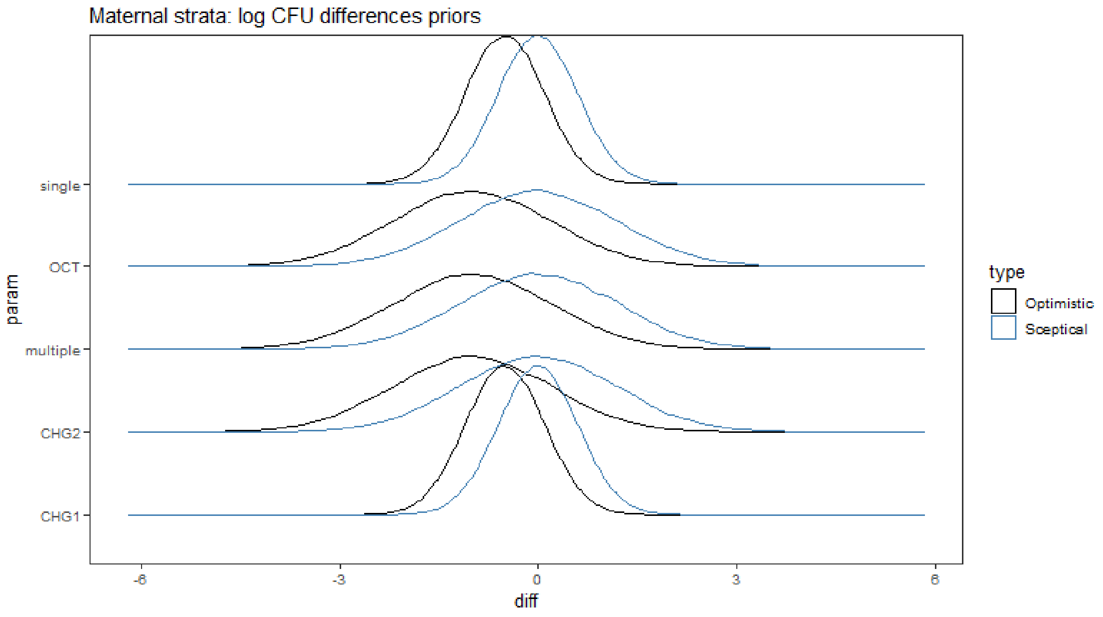

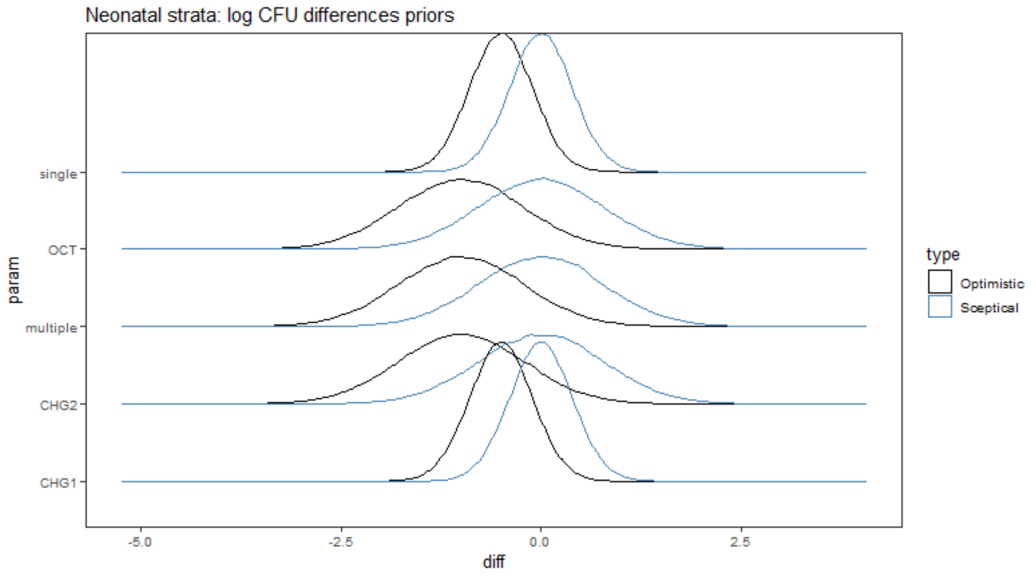

*Note: different scales in each graph*

#### 4. eResults

##### 4.1 Additional baseline variables: eTable 4 and eTable 5

**eTable 4: Maternal additional baseline variables**

| Variable | Overall | 1% CHG (N=43) | 2% CHG (N=43) | OHP (N=42) | Multiple (N=64) | Once (N=64) | SOC (N=21) |
|---|---|---|---|---|---|---|---|
| Gravidity (mean (SD)) | 2.35 (1.36) | 2.44 (1.39) | 2.33 (1.57) | 2.36 (1.25) | 2 (1.08) | 2.75 (1.57) | 2.19 (1.12) |
| Parity (mean (SD)) | 1.26 (1.36) | 1.33 (1.38) | 1.28 (1.59) | 1.29 (1.22) | 0.95 (1.09) | 1.64 (1.58) | 1.05 (1.12) |
| Gestational diabetes | 1 (1%) | 0 (0%) | 0 (0%) | 1 (2%) | 0 (0%) | 1 (2%) | 0 (0%) |
| Cervical dilation at enrolment | 2.06 (1.04) | 2.3 (1.24) | 1.81 (0.82) | 2.1 (1.08) | 2.17 (0.94) | 1.97 (1.19) | 2 (0.84) |
| Rupture of membranes at enrolment | 13 (9%) | 4 (9%) | 4 (9%) | 4 (10%) | 6 (9%) | 6 (9%) | 1 (5%) |
| If ruptured, spontaneous | 13 (100%) | 4 (100%) | 4 (100%) | 4 (100%) | 6 (100%) | 6 (100%) | 1 (100%) |
| Prolonged rupture of membranes | 5 (38%) | 2 (50%) | 1 (25%) | 2 (50%) | 2 (33%) | 3 (50%) | 0 (0%) |
| Offensive liquor | 0 (0%) | 0 (0%) | 0 (0%) | 0 (0%) | 0 (0%) | 0 (0%) | 0 (0%) |

*CHG = chlorhexidine; OHP = octenidine 0.1% combined with phenoxyethanol 2%*

**eTable 5: Neonatal additional baseline variables**

| Variable | Level | Overall | 1% CHG (N=43) | 2% CHG (N=43) | OHP (N=42) | Multiple (N=64) | Once (N=64) | SOC (N=21) |
|---|---|---|---|---|---|---|---|---|
| Prolonged rupture of membranes (>18h) | Yes | 10 (7%) | 3 (7%) | 2 (5%) | 5 (12%) | 3 (5%) | 7 (11%) | 0 (0%) |
| | No | 123 (84%) | 36 (86%) | 37 (88%) | 32 (76%) | 54 (86%) | 51 (81%) | 18 (86%) |
| | Unknown | 14 (10%) | 3 (7%) | 3 (7%) | 5 (12%) | 6 (10%) | 5 (8%) | 3 (14%) |
| Offensive liquor | Yes | 11 (7%) | 2 (5%) | 2 (5%) | 4 (10%) | 5 (8%) | 3 (5%) | 3 (14%) |
| | No | 122 (83%) | 36 (86%) | 36 (86%) | 34 (81%) | 48 (76%) | 58 (92%) | 16 (76%) |
| | Unknown | 14 (10%) | 4 (10%) | 4 (10%) | 4 (10%) | 10 (16%) | 2 (3%) | 2 (10%) |
| Mode of delivery | Vaginal spontaneous | 76 (52%) | 24 (57%) | 18 (43%) | 22 (52%) | 35 (56%) | 29 (46%) | 12 (57%) |
| | Vaginal assisted | 9 (6%) | 2 (5%) | 3 (7%) | 3 (7%) | 2 (3%) | 6 (10%) | 1 (5%) |
| | Emergency caesarean section | 61 (41%) | 15 (36%) | 21 (50%) | 17 (40%) | 25 (40%) | 28 (44%) | 8 (38%) |
| | Unknown | 1 (1%) | 1 (2%) | 0 (0%) | 0 (0%) | 1 (2%) | 0 (0%) | 0 (0%) |
| Maternal HIV status | Positive | 11 (7%) | 4 (10%) | 3 (7%) | 3 (7%) | 5 (8%) | 5 (8%) | 1 (5%) |

| Variable | Level | Overall | 1% CHG (N=43) | 2% CHG (N=43) | OHP (N=42) | Multiple (N=64) | Once (N=64) | SOC (N=21) |
|---|---|---|---|---|---|---|---|---|
| | Negative | 131 (89%) | 36 (86%) | 36 (86%) | 39 (93%) | 56 (89%) | 55 (87%) | 20 (95%) |
| | Unknown | 5 (3%) | 2 (5%) | 3 (7%) | 0 (0%) | 2 (3%) | 3 (5%) | 0 (0%) |
| APGAR Score at 1 minute | 2 | 3 (2%) | 2 (5%) | 1 (2%) | 0 (0%) | 3 (5%) | 0 (0%) | 0 (0%) |
| | 3 | 12 (8%) | 3 (7%) | 3 (7%) | 5 (12%) | 7 (11%) | 4 (6%) | 1 (5%) |
| | 4 | 11 (7%) | 4 (10%) | 2 (5%) | 3 (7%) | 4 (6%) | 5 (8%) | 2 (10%) |
| | 5 | 11 (7%) | 2 (5%) | 4 (10%) | 3 (7%) | 4 (6%) | 5 (8%) | 2 (10%) |
| | 6 | 11 (7%) | 1 (2%) | 2 (5%) | 4 (10%) | 3 (5%) | 4 (6%) | 4 (19%) |
| | 7 | 29 (20%) | 6 (14%) | 12 (29%) | 9 (21%) | 9 (14%) | 18 (29%) | 2 (10%) |
| | 8 | 43 (29%) | 15 (36%) | 12 (29%) | 11 (26%) | 20 (32%) | 18 (29%) | 5 (24%) |
| | 9 | 24 (16%) | 7 (17%) | 6 (14%) | 6 (14%) | 10 (16%) | 9 (14%) | 5 (24%) |
| | Unknown | 3 (2%) | 2 (5%) | 0 (0%) | 1 (2%) | 3 (5%) | 0 (0%) | 0 (0%) |
| APGAR Score at 5 minutes | 4 | 1 (1%) | 1 (2%) | 0 (0%) | 0 (0%) | 1 (2%) | 0 (0%) | 0 (0%) |
| | 5 | 10 (7%) | 1 (2%) | 3 (7%) | 5 (12%) | 8 (13%) | 1 (2%) | 1 (5%) |
| | 6 | 12 (8%) | 5 (12%) | 3 (7%) | 3 (7%) | 4 (6%) | 7 (11%) | 1 (5%) |
| | 7 | 12 (8%) | 4 (10%) | 3 (7%) | 2 (5%) | 5 (8%) | 4 (6%) | 3 (14%) |
| | 8 | 14 (10%) | 3 (7%) | 4 (10%) | 5 (12%) | 4 (6%) | 8 (13%) | 2 (10%) |
| | 9 | 16 (11%) | 2 (5%) | 8 (19%) | 4 (10%) | 5 (8%) | 9 (14%) | 2 (10%) |
| | 10 | 79 (54%) | 24 (57%) | 21 (50%) | 22 (52%) | 33 (52%) | 34 (54%) | 12 (57%) |
| | Unknown | 3 (2%) | 2 (5%) | 0 (0%) | 1 (2%) | 3 (5%) | 0 (0%) | 0 (0%) |
| APGAR Score at 10 minutes | 4 | 1 (1%) | 1 (2%) | 0 (0%) | 0 (0%) | 1 (2%) | 0 (0%) | 0 (0%) |
| | 5 | 1 (1%) | 0 (0%) | 1 (2%) | 0 (0%) | 1 (2%) | 0 (0%) | 0 (0%) |
| | 6 | 2 (1%) | 0 (0%) | 1 (2%) | 1 (2%) | 2 (3%) | 0 (0%) | 0 (0%) |
| | 7 | 13 (9%) | 4 (10%) | 4 (10%) | 2 (5%) | 6 (10%) | 4 (6%) | 3 (14%) |
| | 8 | 12 (8%) | 4 (10%) | 1 (2%) | 5 (12%) | 5 (8%) | 5 (8%) | 2 (10%) |
| | 9 | 9 (6%) | 2 (5%) | 4 (10%) | 2 (5%) | 2 (3%) | 6 (10%) | 1 (5%) |
| | 10 | 79 (54%) | 21 (50%) | 25 (60%) | 20 (48%) | 31 (49%) | 35 (56%) | 13 (62%) |
| | Unknown | 30 (20%) | 10 (24%) | 6 (14%) | 12 (29%) | 15 (24%) | 13 (21%) | 2 (10%) |

*CHG = chlorhexidine; OHP = octenidine 0.1% combined with phenoxyethanol 2%*

#### 4.2 Receipt of interventions: eTable 6, eTable 7, eTable 8 and eTable 9

##### eTable 6: Maternal stratum receipt of interventions by randomisation to 1% CHG, 2% CHG, OHP or standard of care (SOC)

| Variable | Level/Metric | 1% CHG (N=43) | 2% CHG (N=43) | OHP (N=42) | SOC (N=21) | p-value | p-value drug only |
|---|---|---|---|---|---|---|---|
| Total number of antiseptic applications | median | 1 | 1 | 1 | 0 | <0.001 | 0.826 |
| | (IQR) | (1, 2) | (1, 3) | (1, 3) | (0, 0) | | |
| | N | 43 | 43 | 42 | 21 | | |
| Number of antiseptic applications | 0 | 1 (2%) | .. | 1 (2%) | 21 (100%) | <0.001 | 0.77 |
| | 1 | 25 (58%) | 24 (56%) | 23 (55%) | .. | | |
| | 2 | 6 (14%) | 5 (12%) | 3 (7%) | .. | | |
| | 3 | 5 (12%) | 8 (19%) | 11 (26%) | .. | | |
| | 4 | 4 (9%) | 4 (9%) | 1 (2%) | .. | | |
| | 5 | 2 (5%) | 1 (2%) | 2 (5%) | .. | | |
| | 6 | .. | 1 (2%) | 1 (2%) | .. | | |
| Last antiseptic application | 0h | 25 (58%) | 24 (56%) | 23 (55%) | .. | <0.001 | 0.778 |
| | 4h | 6 (14%) | 5 (12%) | 3 (7%) | .. | | |
| | 8h | 5 (12%) | 8 (19%) | 11 (26%) | .. | | |
| | 24h | 4 (9%) | 4 (9%) | 1 (2%) | .. | | |
| | 28h | 2 (5%) | 1 (2%) | 2 (5%) | .. | | |
| | 32h | .. | 1 (2%) | 1 (2%) | .. | | |
| | No antiseptic | 1 (2%) | .. | 1 (2%) | 21 (100%) | | |
| Number of antiseptic applications before 4h swab | median | 1 | 1 | 1 | 0 | <0.001 | n/a |
| | (IQR) | (1, 1) | (1, 1) | (1, 1) | (0, 0) | | |
| | N | 36 | 37 | 37 | 14 | | |
| Number of antiseptic applications before 8h swab | median | 2 | 2 | 2 | 0 | <0.001 | 0.885 |
| | (IQR) | (1, 2) | (1, 2) | (1, 2) | (0, 0) | | |
| | N | 22 | 28 | 28 | 10 | | |
| Number of antiseptic applications before 24h swab | median | 1 | 3 | 1 | 0 | <0.001 | 0.342 |
| | (IQR) | (1, 3) | (1, 3) | (1, 3) | (0, 0) | | |
| | N | 15 | 12 | 10 | 5 | | |
| Number of antiseptic applications before 28h swab | median | 1 | 2 | 2 | 0 | 0.089 | 0.856 |
| | (IQR) | (1, 3) | (2, 3) | (1, 4) | (0, 0) | | |
| | N | 6 | 2 | 6 | 4 | | |
| Number of antiseptic applications before 32h swab | median | 1 | 5 | 5 | 0 | <0.001 | <0.001 |
| | (IQR) | (1, 1) | (5, 5) | (5, 5) | (0, 0) | | |
| | N | 2 | 1 | 1 | 1 | | |
| Skin score assessed at all measured timepoints | | 39 (93%) | 40 (93%) | 41 (100%) | 20 (100%) | 0.322 | 0.278 |

*CHG = chlorhexidine; OHP = octenidine 0.1% combined with phenoxyethanol 2%*

##### eTable 7: Maternal stratum receipt of interventions by randomisation to single or multiple application (excluding SOC)

| Variable | Level/Metric | Once (N=64) | Multiple (N=64) | p-value |
|---|---|---|---|---|
| Total number of antiseptic applications | median | 1 | 3 | <0.001 |
| | (IQR) | (1, 1) | (2, 3) | |
| | N | 64 | 64 | |
| Number of antiseptic applications | 0 | 2 (3%) | .. | <0.001 |
| | 1 | 62 (97%) | 10 (16%) | |
| | 2 | .. | 14 (22%) | |
| | 3 | .. | 24 (38%) | |
| | 4 | .. | 9 (14%) | |
| | 5 | .. | 5 (8%) | |
| | 6 | .. | 2 (3%) | |
| Last antiseptic application | 0h | 62 (97%) | 10 (16%) | <0.001 |
| | 4h | .. | 14 (22%) | |
| | 8h | .. | 24 (38%) | |
| | 24h | .. | 9 (14%) | |
| | 28h | .. | 5 (8%) | |
| | 32h | .. | 2 (3%) | |
| | No antiseptic | 2 (3%) | .. | |
| Number of antiseptic applications before 4h swab | median | 1 | 1 | n/a |
| | (IQR) | (1, 1) | (1, 1) | |
| | N | 56 | 54 | |
| Number of antiseptic applications before 8h swab | median | 1 | 2 | <0.001 |
| | (IQR) | (1, 1) | (2, 2) | |
| | N | 36 | 42 | |
| Number of antiseptic applications before 24h swab | median | 1 | 3 | <0.001 |
| | (IQR) | (1, 1) | (3, 3) | |
| | N | 18 | 19 | |
| Number of antiseptic applications before 28h swab | median | 1 | 4 | <0.001 |
| | (IQR) | (1, 1) | (4, 4) | |
| | N | 8 | 6 | |
| Number of antiseptic applications before 32h swab | median | 1 | 5 | <0.001 |
| | (IQR) | (1, 1) | (5, 5) | |
| | N | 2 | 2 | |
| Skin score assessed at all measured timepoints | | 60 (97%) | 60 (94%) | 0.418 |

*CHG = chlorhexidine; OHP = octenidine 0.1% combined with phenoxyethanol 2%*

#### eTable 8: Neonatal stratum receipt of interventions by randomisation to 1% CHG, 2% CHG, OHP or SOC

| Variable | Level/Metric | 1% CHG (N=42) | 2% CHG (N=42) | OHP (N=42) | SOC (N=21) | p-value | p-value drug only |
|---|---|---|---|---|---|---|---|
| Total number of antiseptic applications | median | 1 | 1 | 1 | 0 | <0.001 | 0.734 |
| | (IQR) | (1, 3) | (1, 2) | (1, 3) | (0, 0) | | |
| | N | 42 | 42 | 42 | 21 | | |
| Number of antiseptic applications | 0 | 1 (2%) | .. | .. | 21 (100%) | <0.001 | 0.619 |
| | 1 | 23 (55%) | 22 (52%) | 22 (52%) | .. | | |
| | 2 | 5 (12%) | 11 (26%) | 6 (14%) | .. | | |
| | 3 | 10 (24%) | 6 (14%) | 9 (21%) | .. | | |
| | 4 | 3 (7%) | 3 (7%) | 5 (12%) | .. | | |
| Last antiseptic application | 0h | 22 (52%) | 22 (52%) | 22 (52%) | .. | <0.001 | 0.809 |
| | 24h | 6 (14%) | 10 (24%) | 6 (14%) | .. | | |
| | 48h | 10 (24%) | 6 (14%) | 9 (21%) | .. | | |
| | 72h | 3 (7%) | 4 (10%) | 5 (12%) | .. | | |
| | No antiseptic | 1 (2%) | .. | .. | 21 (100%) | | |
| Number of antiseptic applications before 24h swab | median | 1 | 1 | 1 | 0 | <0.001 | 0.371 |
| | (IQR) | (1, 1) | (1, 1) | (1, 1) | (0, 0) | | |
| | N | 39 | 39 | 39 | 20 | | |
| Number of antiseptic applications before 48h swab | median | 2 | 1 | 2 | 0 | <0.001 | 0.702 |
| | (IQR) | (1, 2) | (1, 2) | (1, 2) | (0, 0) | | |
| | N | 24 | 21 | 26 | 14 | | |
| Number of antiseptic applications before 72h swab | median | 2 | 1 | 1 | 0 | <0.001 | 0.77 |
| | (IQR) | (1, 3) | (1, 3) | (1, 3) | (0, 0) | | |
| | N | 6 | 9 | 15 | 9 | | |
| Skin score assessed before all antiseptic applications / swabs | | 41 (100%) | 42 (100%) | 42 (100%) | 21 (100%) | n/a | n/a |
| Skin score assessed after all antiseptic applications | | 41 (100%) | 42 (100%) | 42 (100%) | .. | n/a | n/a |
| Temperature assessed before all antiseptic applications / swabs | | 40 (98%) | 42 (100%) | 41 (98%) | 21 (100%) | n/a | n/a |
| Temperature assessed after all antiseptic applications | | 41 (100%) | 41 (98%) | 42 (100%) | .. | n/a | n/a |

*CHG = chlorhexidine; OHP = octenidine 0.1% combined with phenoxyethanol 2%*

#### eTable 9: Neonatal stratum receipt of interventions by randomisation to single or multiple application (excluding SOC)

| Variable | Level/Metric | Once (N=63) | Multiple (N=63) | p-value |
|---|---|---|---|---|
| Total number of antiseptic applications | median | 1 | 3 | <0.001 |
| | (IQR) | (1, 1) | (2, 3) | |
| | N | 63 | 63 | |
| Number of antiseptic applications | 0 | .. | 1 (2%) | <0.001 |
| | 1 | 63 (100%) | 4 (6%) | |
| | 2 | .. | 22 (35%) | |
| | 3 | .. | 25 (40%) | |
| | 4 | .. | 11 (17%) | |
| Last antiseptic application | 0h | 63 (100%) | 3 (5%) | <0.001 |
| | 24h | .. | 22 (35%) | |
| | 48h | .. | 25 (40%) | |
| | 72h | .. | 12 (19%) | |
| | No antiseptic | | 1 (2%) | |
| Number of antiseptic applications before 24h swab | median | 1 | 1 | 0.315 |
| | (IQR) | (1, 1) | (1, 1) | |
| | N | 59 | 58 | |
| Number of antiseptic applications before 48h swab | median | 1 | 2 | <0.001 |
| | (IQR) | (1, 1) | (2, 2) | |
| | N | 35 | 36 | |
| Number of antiseptic applications before 72h swab | median | 1 | 3 | <0.001 |
| | (IQR) | (1, 1) | (3, 3) | |
| | N | 18 | 12 | |
| Skin score assessed before all antiseptic applications/swabs | | 63 (100%) | 62 (100%) | n/a |
| Skin score assessed after all antiseptic applications | | 63 (100%) | 62 (100%) | n/a |
| Temperature assessed before all antiseptic applications/swabs | | 61 (97%) | 62 (100%) | n/a |
| Temperature assessed after all antiseptic applications | | 63 (100%) | 61 (98%) | n/a |

*CHG = chlorhexidine; OHP = octenidine 0.1% combined with phenoxyethanol 2%*

##### 4.3 Additional secondary outcomes: eTable 10 and eTable 11

#### eTable 10: Additional secondary outcomes maternal stratum

| Variable | Arm | Change baseline to 4h mean (SD) [N] | Change baseline to 8h mean (SD) [N] | Change baseline to 24h mean (SD) [N] | Change baseline to 28h mean (SD) [N] | Change baseline to 32h mean (SD) [N] | Modelled effect size (95% CI) | p-value |
|---|---|---|---|---|---|---|---|---|
| Gram pos $log_{10}$CFU | 1% CHG (N=43) | -4.2 (2.2) [35] | -3.8 (2.4) [22] | -2.8 (2.7) [15] | -3.5 (1.5) [6] | -4.0 (2.1) [2] | .. | |
| | 2% CHG (N=43) | -5.0 (1.9) [37] | -4.8 (2.5) [28] | -4.5 (3.0) [12] | -2.4 (1.0) [3] | -2.4 (NA) [1] | -0.5 (-1.2, 0.2) | 0.17 |
| | OHP (N=42) | -2.7 (1.9) [36] | -3.5 (2.1) [27] | -2.9 (3.9) [10] | -4.5 (4.8) [6] | -8.7 (NA) [1] | 1.2 (0.5, 2.0) | 0.0013 |
| | Once (N=64) | -4.0 (2.1) [56] | -3.5 (2.6) [36] | -2.3 (2.5) [18] | -2.0 (1.5) [8] | -4.0 (2.1) [2] | .. | |
| | Multiple (N=64) | -3.9 (2.3) [52] | -4.5 (2.1) [41] | -4.5 (3.4) [19] | -5.6 (3.5) [7] | -5.5 (4.5) [2] | -0.6 (-1.2, 0.0) | 0.063 |
| | SOC (N=21) | -0.3 (1.0) [14] | -1.1 (2.1) [10] | -1.9 (2.8) [5] | 0.2 (2.3) [4] | -2.6 (NA) [1] | 2.6 (1.6, 3.6) | <0.0001 |
| Yeast $log_{10}$CFU | 1% CHG (N=43) | 0.2 (1.0) [35] | 0.5 (1.8) [22] | 0.5 (1.2) [15] | 0.0 (0.0) [6] | 1.3 (1.9) [2] | .. | |
| | 2% CHG (N=43) | -0.4 (1.6) [37] | -0.4 (1.4) [28] | -0.1 (2.3) [12] | -0.3 (0.5) [3] | 0.0 (NA) [1] | -0.6 (-1.1, -0.1) | 0.03 |
| | OHP (N=42) | -0.1 (0.6) [36] | 0.0 (1.2) [27] | -0.2 (0.5) [10] | -0.3 (0.6) [6] | 0.0 (NA) [1] | -0.4 (-1.0, 0.1) | 0.091 |
| | Once (N=64) | -0.0 (1.4) [56] | 0.2 (1.8) [36] | -0.1 (1.8) [18] | 0.0 (0.0) [8] | 1.3 (1.9) [2] | .. | |
| | Multiple (N=64) | -0.2 (0.8) [52] | -0.2 (1.1) [41] | 0.3 (1.2) [19] | -0.4 (0.6) [7] | 0.0 (0.0) [2] | -0.2 (-0.6, 0.2) | 0.31 |
| | SOC (N=21) | -0.1 (0.3) [14] | 0.3 (1.7) [10] | 0.3 (0.7) [5] | -1.6 (2.4) [4] | 2.7 (NA) [1] | -0.4 (-1.1, 0.3) | 0.29 |
| *Enterococcus* spp. $log_{10}$CFU | 1% CHG (N=43) | -2.3 (2.5) [35] | -2.3 (2.3) [22] | -2.0 (2.4) [15] | -2.7 (1.2) [6] | -2.1 (1.2) [2] | .. | |
| | 2% CHG (N=43) | -2.5 (2.6) [37] | -2.3 (2.8) [28] | -2.2 (3.1) [12] | -1.7 (4.3) [3] | 2.5 (NA) [1] | -0.1 (-0.8, 0.5) | 0.64 |
| | OHP (N=42) | -1.6 (2.1) [36] | -2.1 (2.2) [27] | -1.5 (2.9) [10] | -2.3 (3.1) [6] | -2.3 (NA) [1] | 0.6 (0.0, 1.3) | 0.045 |
| | Once (N=64) | -2.2 (2.1) [56] | -1.8 (2.0) [36] | -1.3 (2.0) [18] | -2.1 (2.2) [8] | -2.1 (1.2) [2] | .. | |
| | Multiple (N=64) | -2.1 (2.7) [52] | -2.5 (2.8) [41] | -2.5 (3.2) [19] | -2.6 (3.1) [7] | 0.1 (3.4) [2] | -0.2 (-0.7, 0.3) | 0.43 |
| | SOC (N=21) | -1.2 (1.4) [14] | -0.9 (1.8) [10] | -1.5 (1.3) [5] | 1.1 (0.9) [4] | -0.4 (NA) [1] | 1.1 (0.2, 2.0) | 0.012 |
| *S. aureus* $log_{10}$CFU | 1% CHG (N=43) | -0.1 (0.4) [35] | 0.0 (0.0) [22] | 0.0 (0.0) [15] | 0.0 (0.0) [6] | 0.0 (0.0) [2] | .. | |
| | 2% CHG (N=43) | -0.3 (0.8) [37] | -0.3 (0.9) [28] | -0.3 (1.0) [12] | -1.1 (1.9) [3] | 0.0 (NA) [1] | -0.0 (-0.1, 0.1) | 0.82 |
| | OHP (N=42) | -0.1 (0.5) [36] | 0.2 (0.6) [27] | 0.0 (0.0) [10] | 0.0 (0.0) [6] | 0.0 (NA) [1] | 0.1 (0.0, 0.1) | 0.039 |

| Variable | Arm | Change baseline to 4h mean (SD) [N] | Change baseline to 8h mean (SD) [N] | Change baseline to 24h mean (SD) [N] | Change baseline to 28h mean (SD) [N] | Change baseline to 32h mean (SD) [N] | Modelled effect size (95% CI) | p-value |
|---|---|---|---|---|---|---|---|---|
| | Once (N=64) | -0.1 (0.5) [56] | -0.0 (0.8) [36] | -0.2 (0.8) [18] | -0.4 (1.2) [8] | 0.0 (0.0) [2] | .. | |
| | Multiple (N=64) | -0.2 (0.7) [52] | -0.1 (0.5) [41] | 0.0 (0.0) [19] | 0.0 (0.0) [7] | 0.0 (0.0) [2] | -0.0 (-0.1, 0.0) | 0.62 |
| | SOC (N=21) | 0.1 (0.4) [14] | 0.0 (0.0) [10] | 0.0 (0.0) [5] | 0.0 (0.0) [4] | 0.0 (NA) [1] | 0.0 (-0.1, 0.1) | 0.38 |
| GBS $log_{10}$CFU | 1% CHG (N=43) | -0.3 (1.1) [35] | -0.3 (1.2) [22] | -0.2 (0.9) [15] | 0.0 (0.0) [6] | 0.0 (0.0) [2] | .. | |
| | 2% CHG (N=43) | -0.4 (1.0) [37] | -0.3 (0.8) [28] | -0.6 (1.0) [12] | -1.0 (1.7) [3] | 0.0 (NA) [1] | 0.0 (-0.2, 0.2) | 0.9 |
| | OHP (N=42) | -0.0 (1.2) [36] | -0.3 (2.0) [27] | -0.7 (3.9) [10] | -1.6 (4.0) [6] | -8.9 (NA) [1] | 0.2 (0.0, 0.5) | 0.038 |
| | Once (N=64) | -0.3 (1.4) [56] | -0.4 (1.2) [36] | -0.2 (1.6) [18] | -0.4 (1.0) [8] | 0.0 (0.0) [2] | .. | |
| | Multiple (N=64) | -0.2 (0.7) [52] | -0.2 (1.6) [41] | -0.7 (2.5) [19] | -1.4 (3.7) [7] | -4.4 (6.3) [2] | -0.1 (-0.2, 0.1) | 0.54 |
| | SOC (N=21) | 0.2 (0.7) [14] | 0.2 (0.5) [10] | 0.0 (0.0) [5] | 0.0 (0.0) [4] | 0.0 (NA) [1] | 0.2 (-0.1, 0.5) | 0.26 |
| *Klebsiella spp.* $log_{10}$CFU | 1% CHG (N=43) | -0.1 (0.6) [35] | -0.2 (0.7) [22] | 0.7 (1.8) [15] | 0.2 (2.3) [6] | 0.2 (0.2) [2] | .. | |
| | 2% CHG (N=43) | -0.0 (0.2) [37] | 0.1 (0.5) [28] | 0.5 (1.3) [12] | 0.0 (0.0) [3] | 0.0 (NA) [1] | -0.0 (-0.4, 0.3) | 0.97 |
| | OHP (N=42) | -0.1 (1.1) [36] | -0.4 (0.9) [27] | 0.2 (1.6) [10] | -0.5 (0.9) [6] | -1.0 (NA) [1] | 0.1 (-0.2, 0.5) | 0.51 |
| | Once (N=64) | -0.1 (0.4) [56] | 0.0 (0.6) [36] | 0.8 (1.5) [18] | 0.5 (1.5) [8] | 0.2 (0.2) [2] | .. | |
| | Multiple (N=64) | -0.1 (1.0) [52] | -0.3 (0.8) [41] | 0.2 (1.5) [19] | -0.8 (1.3) [7] | -0.5 (0.7) [2] | -0.1 (-0.4, 0.2) | 0.62 |
| | SOC (N=21) | 0.0 (0.8) [14] | 1.1 (1.9) [10] | 0.7 (1.6) [5] | 1.4 (2.9) [4] | 2.9 (NA) [1] | 0.4 (-0.1, 0.9) | 0.11 |
| *Serratia* spp. $log_{10}$CFU | 1% CHG (N=43) | -0.0 (0.3) [35] | 0.0 (0.1) [22] | 0.0 (0.0) [15] | 0.0 (0.0) [6] | 0.0 (0.0) [2] | .. | |
| | 2% CHG (N=43) | -0.1 (0.5) [37] | -0.1 (0.6) [28] | -0.2 (0.8) [12] | -1.0 (1.7) [3] | -2.9 (NA) [1] | -0.0 (-0.0, 0.0) | 0.071 |
| | OHP (N=42) | -0.1 (0.3) [36] | -0.1 (0.3) [27] | 0.0 (0.0) [10] | 0.0 (0.0) [6] | 0.0 (NA) [1] | -0.0 (-0.0, 0.0) | 0.07 |
| | Once (N=64) | -0.0 (0.2) [56] | -0.0 (0.1) [36] | 0.0 (0.0) [18] | 0.0 (0.0) [8] | 0.0 (0.0) [2] | .. | |
| | Multiple (N=64) | -0.1 (0.5) [52] | -0.1 (0.5) [41] | -0.2 (0.7) [19] | -0.4 (1.1) [7] | -1.5 (2.1) [2] | -0.0 (-0.0, 0.0) | 0.84 |
| | SOC (N=21) | 0.0 (0.0) [14] | 0.0 (0.0) [10] | 0.0 (0.0) [5] | 0.0 (0.0) [4] | 0.0 (NA) [1] | -0.0 (-0.0, 0.0) | 0.17 |
| *E. coli* $log_{10}$CFU | 1% CHG (N=43) | -1.4 (1.8) [35] | -1.5 (1.9) [22] | -0.5 (2.4) [15] | -2.1 (2.4) [6] | -1.3 (1.8) [2] | .. | |
| | 2% CHG (N=43) | -0.9 (1.3) [37] | -0.6 (1.5) [28] | -0.5 (1.2) [12] | -0.2 (0.3) [3] | 2.3 (NA) [1] | 0.1 (-0.4, 0.6) | 0.69 |
| | OHP (N=42) | -0.6 (2.3) [36] | -1.1 (2.3) [27] | -0.8 (3.6) [10] | -1.1 (4.2) [6] | 0.0 (NA) [1] | 0.8 (0.3, 1.3) | 0.0034 |
| | Once (N=64) | -0.9 (1.6) [56] | -0.5 (1.6) [36] | 0.1 (1.9) [18] | -1.0 (1.1) [8] | -1.3 (1.8) [2] | .. | |
| | Multiple (N=64) | -1.0 (2.0) [52] | -1.5 (2.1) [41] | -1.2 (2.7) [19] | -1.7 (4.4) [7] | 1.2 (1.6) [2] | -0.2 (-0.6, 0.2) | 0.36 |

| Variable | Arm | Change baseline to 4h mean (SD) [N] | Change baseline to 8h mean (SD) [N] | Change baseline to 24h mean (SD) [N] | Change baseline to 28h mean (SD) [N] | Change baseline to 32h mean (SD) [N] | Modelled effect size (95% CI) | p-value |
|---|---|---|---|---|---|---|---|---|
| | SOC (N=21) | -0.4 (1.1) [14] | -0.2 (1.7) [10] | 0.7 (1.0) [5] | 3.2 (3.6) [4] | 5.8 (NA) [1] | 0.7 (-0.0, 1.4) | 0.053 |
| *Enterobacter* spp. $log_{10}CFU$ | 1% CHG (N=43) | -0.1 (0.4) [35] | 0.0 (0.0) [22] | 0.2 (0.8) [15] | 0.0 (0.0) [6] | 0.0 (0.0) [2] | .. | |
| | 2% CHG (N=43) | -0.0 (0.2) [37] | -0.0 (0.3) [28] | -0.1 (0.4) [12] | -0.4 (0.8) [3] | 0.3 (NA) [1] | -0.1 (-0.3, 0.2) | 0.54 |
| | OHP (N=42) | -0.3 (1.1) [36] | -0.0 (0.8) [27] | 0.8 (1.7) [10] | -0.3 (0.7) [6] | 0.0 (NA) [1] | 0.0 (-0.2, 0.3) | 0.71 |
| | Once (N=64) | -0.1 (0.4) [56] | -0.0 (0.2) [36] | -0.1 (0.3) [18] | -0.2 (0.5) [8] | 0.0 (0.0) [2] | .. | |
| | Multiple (N=64) | -0.2 (0.9) [52] | -0.0 (0.6) [41] | 0.6 (1.4) [19] | -0.3 (0.7) [7] | 0.2 (0.2) [2] | 0.1 (-0.1, 0.3) | 0.18 |
| | SOC (N=21) | -0.3 (1.0) [14] | -0.4 (1.1) [10] | -0.1 (0.2) [5] | 0.0 (0.0) [4] | 0.0 (NA) [1] | -0.2 (-0.4, 0.1) | 0.29 |
| *Proteus* spp. $log_{10}CFU$ | 1% CHG (N=43) | -0.0 (0.2) [35] | 0.0 (0.0) [22] | 0.0 (0.0) [15] | 0.0 (0.0) [6] | 0.0 (0.0) [2] | .. | |
| | 2% CHG (N=43) | 0.0 (0.0) [37] | 0.0 (0.3) [28] | 0.0 (0.0) [12] | 0.0 (0.0) [3] | 0.0 (NA) [1] | 0.0 (-0.0, 0.0) | 0.24 |
| | OHP (N=42) | 0.0 (0.0) [36] | 0.0 (0.0) [27] | 0.0 (0.0) [10] | 0.0 (0.0) [6] | 0.0 (NA) [1] | -0.0 (-0.0, 0.0) | 0.97 |
| | Once (N=64) | -0.0 (0.2) [56] | 0.0 (0.0) [36] | 0.0 (0.0) [18] | 0.0 (0.0) [8] | 0.0 (0.0) [2] | .. | |
| | Multiple (N=64) | 0.0 (0.0) [52] | 0.0 (0.2) [41] | 0.0 (0.0) [19] | 0.0 (0.0) [7] | 0.0 (0.0) [2] | 0.0 (-0.0, 0.0) | 0.36 |
| | SOC (N=21) | 0.0 (0.0) [14] | 0.0 (0.0) [10] | 0.0 (0.0) [5] | 0.0 (0.0) [4] | 0.0 (NA) [1] | 0.0 (-0.0, 0.0) | 0.8 |
| *Pseudomonas* spp. $log_{10}CFU$ | 1% CHG (N=43) | -0.1 (0.6) [35] | -0.2 (0.7) [22] | -0.2 (0.6) [15] | 0.0 (0.0) [6] | 0.0 (0.0) [2] | .. | |
| | 2% CHG (N=43) | -0.0 (0.2) [37] | 0.0 (0.0) [28] | 0.0 (0.0) [12] | 0.0 (0.0) [3] | 0.0 (NA) [1] | -0.0 (-0.0, 0.0) | 0.42 |
| | OHP (N=42) | -0.1 (0.8) [36] | -0.2 (0.9) [27] | 0.0 (0.0) [10] | 0.0 (0.0) [6] | 0.0 (NA) [1] | -0.0 (-0.0, 0.0) | 0.2 |
| | Once (N=64) | -0.0 (0.2) [56] | 0.0 (0.0) [36] | 0.0 (0.0) [18] | 0.0 (0.0) [8] | 0.0 (0.0) [2] | .. | |
| | Multiple (N=64) | -0.2 (0.8) [52] | -0.2 (0.9) [41] | -0.1 (0.6) [19] | 0.0 (0.0) [7] | 0.0 (0.0) [2] | 0.0 (-0.0, 0.0) | 0.73 |
| | SOC (N=21) | 0.0 (0.0) [14] | 0.0 (0.0) [10] | 0.0 (0.0) [5] | 0.0 (0.0) [4] | 0.0 (NA) [1] | -0.0 (-0.0, 0.0) | 0.59 |
| *Acinetobacter* spp. $log_{10}CFU$ | 1% CHG (N=43) | -1.3 (1.9) [35] | -1.1 (2.2) [22] | -1.8 (2.5) [15] | -1.9 (2.2) [6] | -2.4 (3.4) [2] | .. | |
| | 2% CHG (N=43) | -1.9 (2.5) [37] | -1.9 (2.6) [28] | -1.2 (2.4) [12] | -1.4 (2.5) [3] | 0.0 (NA) [1] | -0.2 (-0.6, 0.2) | 0.32 |
| | OHP (N=42) | -1.4 (2.1) [36] | -1.2 (1.9) [27] | -1.2 (2.0) [10] | -1.5 (1.9) [6] | -3.8 (NA) [1] | 0.1 (-0.3, 0.5) | 0.73 |
| | Once (N=64) | -1.7 (2.2) [56] | -1.4 (2.2) [36] | -1.9 (2.3) [18] | -2.6 (2.0) [8] | -2.4 (3.4) [2] | .. | |
| | Multiple (N=64) | -1.4 (2.2) [52] | -1.5 (2.4) [41] | -1.1 (2.4) [19] | -0.5 (1.4) [7] | -1.9 (2.7) [2] | -0.0 (-0.3, 0.3) | 0.89 |
| | SOC (N=21) | 0.1 (1.6) [14] | 0.1 (0.6) [10] | 0.1 (0.3) [5] | -1.1 (2.2) [4] | 0.0 (NA) [1] | 1.5 (1.0, 2.1) | <0.0001 |

*CFU = colony forming units; CHG = chlorhexidine' GBS = Group B Streptococcus; OHP = octenidine 0.1% combined with phenoxyethanol 2%; SOC = standard of care*

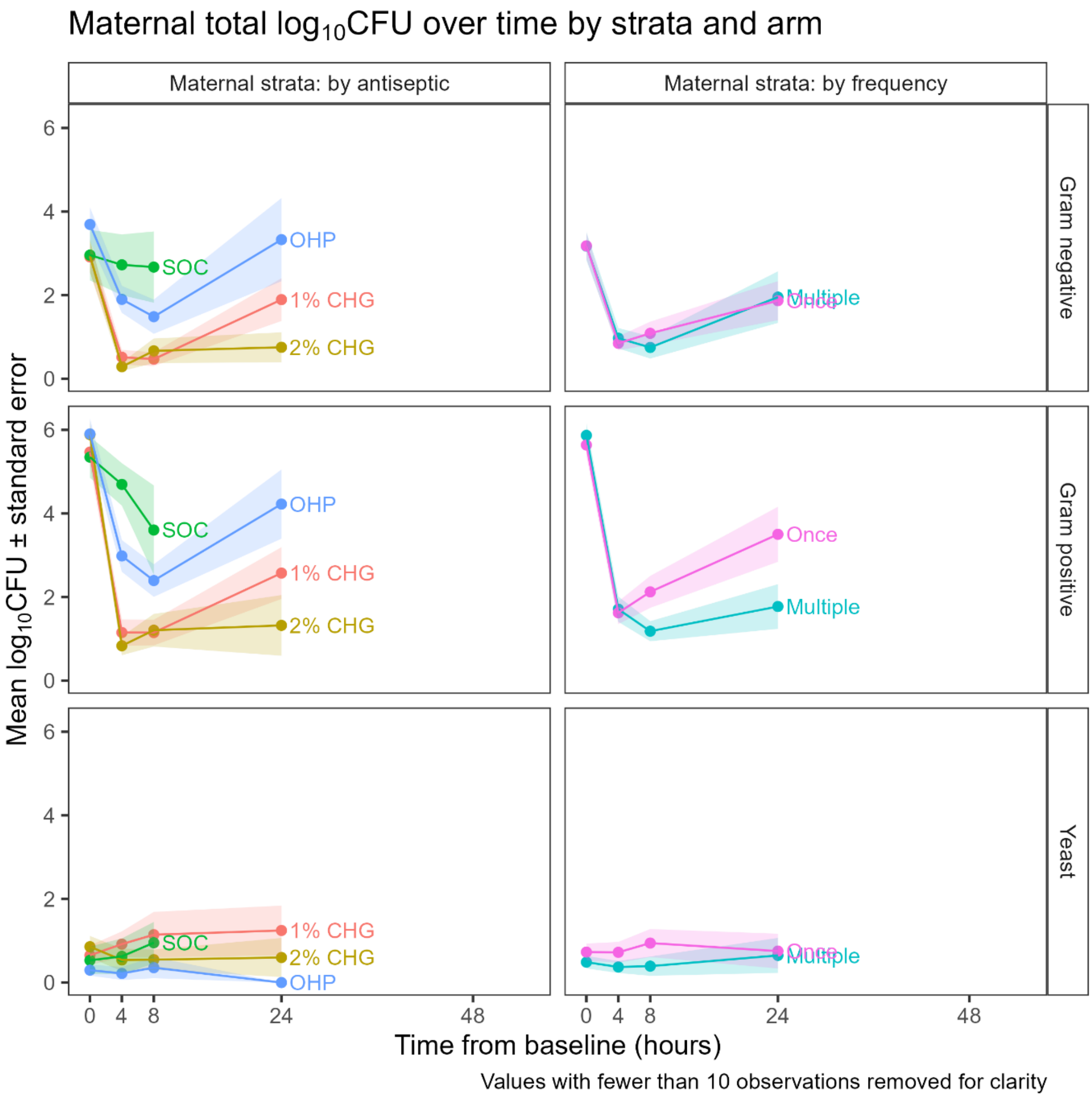
Maternal total log10CFU over time by strata and arm
Maternal strata: by antiseptic
Maternal strata: by frequency
Gram negative
Gram positive
Yeast
OHP
SOC
1% CHG
2% CHG
Multiple
Once
Mean log10CFU ± standard error
Time from baseline (hours)
0 4 8 24 48
Values with fewer than 10 observations removed for clarity

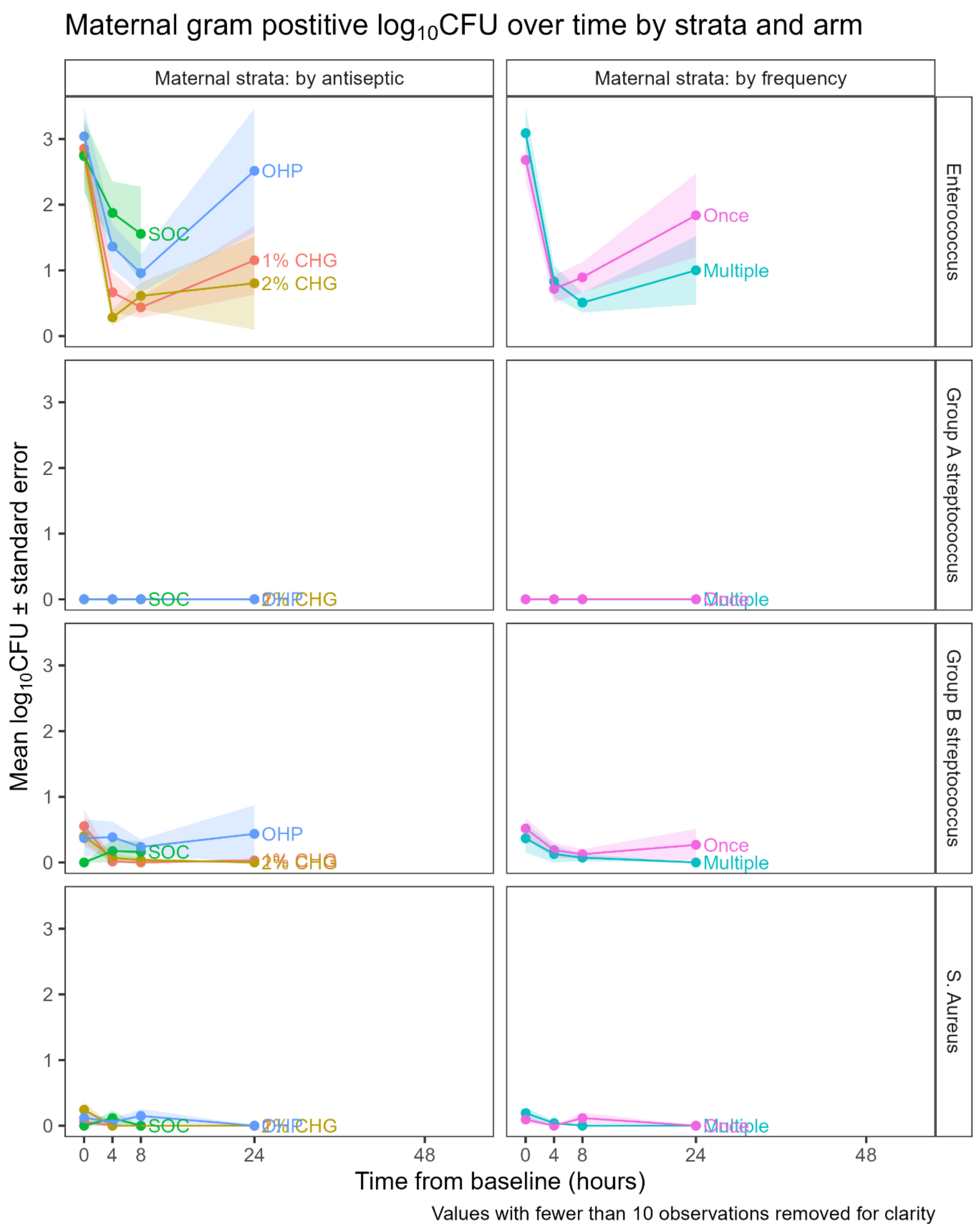
Maternal gram postitive log10CFU over time by strata and arm
Maternal strata: by antiseptic
Maternal strata: by frequency
Enterococcus
Group A streptococcus
Group B streptococcus
S. Aureus
OHP
SOC
1% CHG
2% CHG
Once
Multiple
Mean log10CFU ± standard error
Time from baseline (hours)
Values with fewer than 10 observations removed for clarity

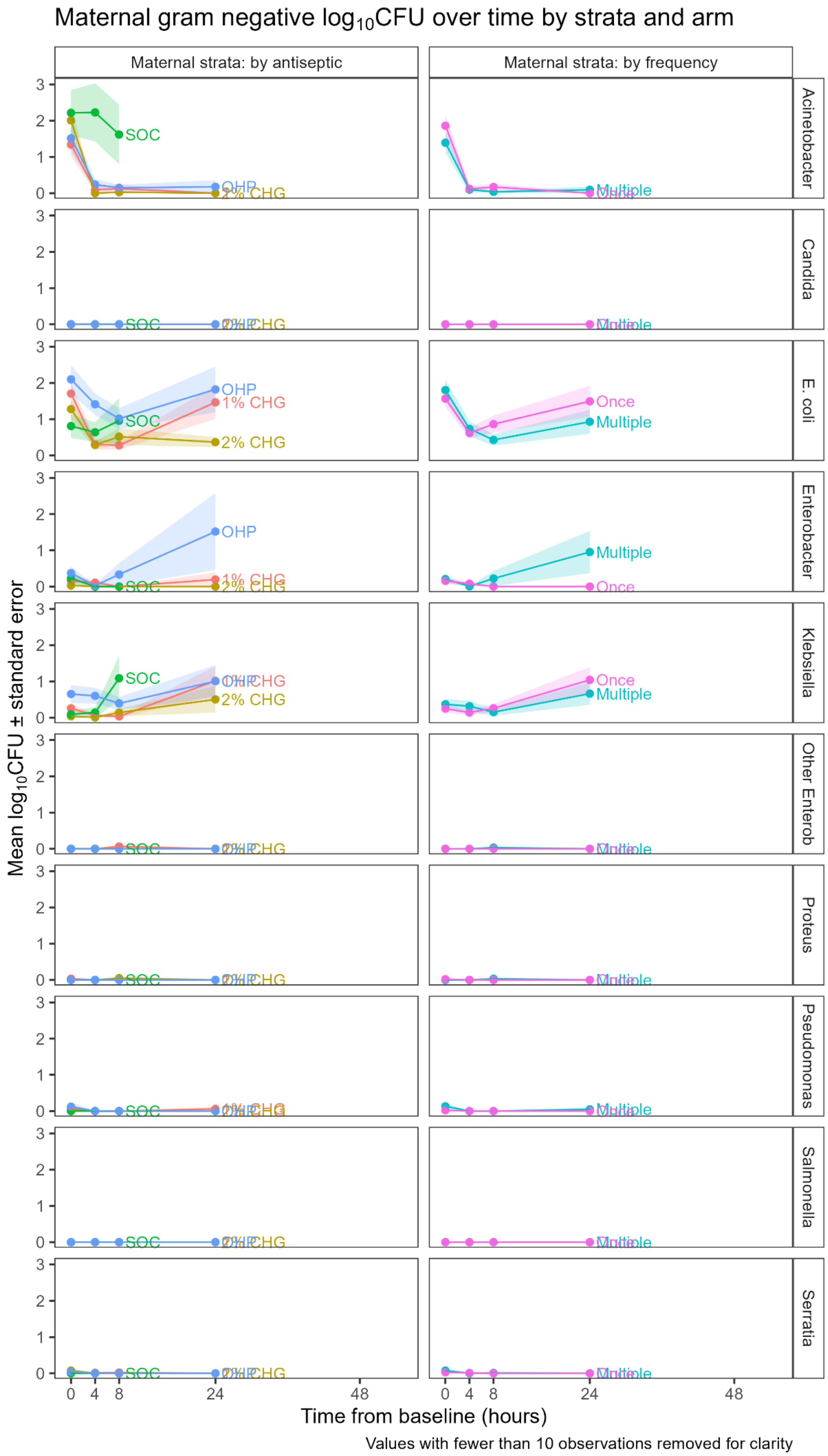

Maternal gram negative log10CFU over time by strata and arm
Maternal strata: by antiseptic
Maternal strata: by frequency
Acinetobacter
Candida
E. coli
Enterobacter
Klebsiella
Other Enterob
Proteus
Pseudomonas
Salmonella
Serratia
SOC
OHP
1% CHG
2% CHG
Once
Multiple
Mean log10CFU ± standard error
Time from baseline (hours)
Values with fewer than 10 observations removed for clarity

#### eTable 11: additional secondary outcomes neonatal stratum

| Variable | Arm | Change baseline to 24h mean (SD) [N] | Change baseline to 48h mean (SD) [N] | Change baseline to 72h mean (SD) [N] | Modelled effect size (95% CI) | p-value |
|---|---|---|---|---|---|---|
| Gram pos $log_{10}$CFU | 1% CHG (N=42) | 0.3 (3.3) [39] | 1.7 (3.1) [24] | 3.3 (2.9) [6] | .. | |
| | 2% CHG (N=42) | 0.3 (2.7) [39] | 2.2 (2.6) [21] | 2.1 (5.1) [9] | -0.5 (-1.4, 0.4) | 0.29 |
| | OHP (N=42) | 1.2 (3.6) [39] | 3.5 (3.2) [26] | 4.1 (3.2) [15] | 0.8 (-0.1, 1.7) | 0.096 |
| | Once (N=63) | 0.9 (2.8) [59] | 3.0 (3.0) [35] | 4.6 (2.4) [18] | .. | |
| | Multiple (N=63) | 0.3 (3.6) [58] | 2.0 (3.1) [36] | 1.4 (4.7) [12] | -0.6 (-1.3, 0.2) | 0.12 |
| | SOC (N=21) | 1.4 (3.6) [20] | 2.7 (3.5) [14] | 3.7 (3.6) [9] | 1.3 (0.1, 2.4) | 0.035 |
| Yeast $log_{10}$CFU | 1% CHG (N=42) | 0.1 (0.6) [39] | 0.0 (0.0) [24] | 0.0 (0.0) [6] | .. | |
| | 2% CHG (N=42) | 0.0 (0.2) [39] | 0.0 (0.0) [21] | 0.0 (0.0) [9] | -0.1 (-0.2, 0.1) | 0.45 |
| | OHP (N=42) | 0.0 (0.0) [39] | 0.0 (0.0) [26] | 0.0 (0.0) [15] | -0.1 (-0.2, 0.1) | 0.32 |
| | Once (N=63) | 0.1 (0.5) [59] | 0.0 (0.0) [35] | 0.0 (0.0) [18] | .. | |
| | Multiple (N=63) | 0.1 (0.3) [58] | 0.0 (0.0) [36] | 0.0 (0.0) [12] | -0.0 (-0.1, 0.1) | 0.87 |
| | SOC (N=21) | 0.3 (1.1) [20] | 0.0 (0.0) [14] | 0.0 (0.0) [9] | 0.0 (-0.1, 0.2) | 0.57 |
| *Enterococcus* spp. $log_{10}$CFU | 1% CHG (N=42) | 0.6 (3.0) [39] | 2.6 (3.1) [24] | 4.7 (3.1) [6] | .. | |
| | 2% CHG (N=42) | 0.6 (2.7) [39] | 2.3 (2.9) [21] | 2.4 (5.6) [9] | -0.5 (-1.6, 0.5) | 0.32 |
| | OHP (N=42) | 0.8 (2.7) [39] | 2.8 (3.1) [26] | 3.3 (2.9) [15] | -0.2 (-1.3, 0.8) | 0.65 |
| | Once (N=63) | 0.9 (2.7) [59] | 3.2 (2.9) [35] | 4.3 (2.7) [18] | .. | |
| | Multiple (N=63) | 0.4 (2.9) [58] | 2.0 (3.1) [36] | 1.9 (5.0) [12] | -0.5 (-1.4, 0.3) | 0.21 |
| | SOC (N=21) | 1.3 (4.5) [20] | 2.3 (4.3) [14] | 2.5 (4.3) [9] | -0.3 (-1.6, 1.1) | 0.68 |
| *S. aureus* $log_{10}$CFU | 1% CHG (N=42) | 0.1 (0.6) [39] | 0.3 (1.3) [24] | 0.0 (0.0) [6] | .. | |
| | 2% CHG (N=42) | -0.4 (1.1) [39] | -0.3 (1.4) [21] | -0.7 (1.5) [9] | -0.2 (-0.5, 0.2) | 0.29 |
| | OHP (N=42) | 0.0 (0.9) [39] | 0.3 (1.5) [26] | 0.3 (1.6) [15] | 0.0 (-0.3, 0.4) | 0.91 |
| | Once (N=63) | 0.0 (1.0) [59] | 0.2 (1.5) [35] | 0.0 (1.8) [18] | .. | |
| | Multiple (N=63) | -0.2 (0.8) [58] | 0.0 (1.3) [36] | -0.2 (0.6) [12] | -0.1 (-0.4, 0.2) | 0.38 |

| Variable | Arm | Change baseline to 24h mean (SD) [N] | Change baseline to 48h mean (SD) [N] | Change baseline to 72h mean (SD) [N] | Modelled effect size (95% CI) | p-value |
|---|---|---|---|---|---|---|
| | SOC (N=21) | -0.5 (1.2) [20] | -0.5 (1.1) [14] | -0.4 (1.1) [9] | -0.3 (-0.7, 0.1) | 0.15 |
| GBS $log_{10}$CFU | 1% CHG (N=42) | -0.1 (0.3) [39] | -0.1 (0.3) [24] | 0.0 (0.0) [6] | .. | |
| | 2% CHG (N=42) | 0.0 (0.0) [39] | 0.0 (0.0) [21] | 0.0 (0.0) [9] | 0.0 (-0.2, 0.2) | 1 |
| | OHP (N=42) | 0.0 (0.7) [39] | 0.1 (0.8) [26] | 0.2 (1.0) [15] | 0.2 (0.0, 0.3) | 0.037 |
| | Once (N=63) | 0.1 (0.5) [59] | 0.1 (0.5) [35] | 0.2 (0.9) [18] | .. | |
| | Multiple (N=63) | -0.1 (0.3) [58] | -0.1 (0.4) [36] | 0.0 (0.0) [12] | -0.1 (-0.2, 0.0) | 0.19 |
| | SOC (N=21) | -0.7 (1.4) [20] | -0.8 (1.6) [14] | -0.8 (1.5) [9] | -0.0 (-0.2, 0.2) | 0.69 |
| *Klebsiella* spp. $log_{10}$CFU | 1% CHG (N=42) | 1.0 (2.1) [39] | 1.5 (2.0) [24] | 3.8 (3.1) [6] | .. | |
| | 2% CHG (N=42) | 0.9 (1.7) [39] | 1.0 (2.1) [21] | 1.6 (2.1) [9] | -0.1 (-0.9, 0.6) | 0.7 |
| | OHP (N=42) | 0.8 (1.9) [39] | 1.0 (1.9) [26] | 1.5 (2.3) [15] | -0.4 (-1.1, 0.4) | 0.34 |
| | Once (N=63) | 0.8 (1.8) [59] | 1.0 (2.0) [35] | 1.7 (2.5) [18] | .. | |
| | Multiple (N=63) | 1.0 (2.0) [58] | 1.3 (2.0) [36] | 2.5 (2.5) [12] | 0.3 (-0.3, 0.9) | 0.37 |
| | SOC (N=21) | -0.2 (1.2) [20] | 0.2 (2.0) [14] | 0.4 (2.6) [9] | -1.1 (-2.1, -0.1) | 0.026 |
| *Serratia* spp. $log_{10}$CFU | 1% CHG (N=42) | 0.2 (1.0) [39] | 0.1 (0.4) [24] | 0.9 (2.2) [6] | .. | |
| | 2% CHG (N=42) | 0.4 (1.3) [39] | -0.2 (1.2) [21] | 0.0 (0.0) [9] | 0.1 (-0.2, 0.4) | 0.57 |
| | OHP (N=42) | 0.1 (0.7) [39] | 0.0 (0.0) [26] | 0.0 (0.0) [15] | -0.1 (-0.4, 0.1) | 0.33 |
| | Once (N=63) | 0.3 (1.2) [59] | -0.1 (0.9) [35] | 0.0 (0.0) [18] | .. | |
| | Multiple (N=63) | 0.1 (0.8) [58] | 0.0 (0.3) [36] | 0.5 (1.6) [12] | -0.1 (-0.3, 0.1) | 0.42 |
| | SOC (N=21) | -0.3 (1.2) [20] | -0.4 (1.5) [14] | -0.6 (1.8) [9] | -0.3 (-0.6, 0.0) | 0.089 |
| *E. coli* $log_{10}$CFU | 1% CHG (N=42) | 0.7 (1.8) [39] | 1.0 (2.6) [24] | 1.1 (2.2) [6] | .. | |
| | 2% CHG (N=42) | 0.6 (2.5) [39] | 1.1 (2.0) [21] | 1.8 (2.4) [9] | 0.1 (-0.8, 1.0) | 0.9 |
| | OHP (N=42) | 0.9 (2.3) [39] | 1.8 (2.9) [26] | 1.7 (2.1) [15] | 0.4 (-0.5, 1.3) | 0.39 |
| | Once (N=63) | 0.9 (2.2) [59] | 1.3 (2.8) [35] | 1.3 (1.9) [18] | .. | |
| | Multiple (N=63) | 0.6 (2.3) [58] | 1.3 (2.3) [36] | 2.0 (2.6) [12] | -0.1 (-0.9, 0.6) | 0.71 |
| | SOC (N=21) | 1.2 (2.2) [20] | 1.1 (2.3) [14] | 2.2 (2.4) [9] | 0.6 (-0.6, 1.7) | 0.33 |
| | 1% CHG (N=42) | -0.0 (1.4) [39] | -0.1 (1.7) [24] | 0.7 (1.3) [6] | .. | |

| Variable | Arm | Change baseline to 24h mean (SD) [N] | Change baseline to 48h mean (SD) [N] | Change baseline to 72h mean (SD) [N] | Modelled effect size (95% CI) | p-value |
|---|---|---|---|---|---|---|
| *Enterobacter* spp. $log_{10}$CFU | 2% CHG (N=42) | 0.2 (1.2) [39] | 0.2 (1.4) [21] | 0.5 (1.4) [9] | 0.0 (-0.5, 0.5) | 0.89 |
| | OHP (N=42) | 0.4 (1.5) [39] | 0.4 (2.0) [26] | 0.2 (0.8) [15] | 0.2 (-0.3, 0.7) | 0.5 |
| | Once (N=63) | 0.2 (1.1) [59] | 0.2 (1.6) [35] | 0.4 (1.2) [18] | .. | |
| | Multiple (N=63) | 0.2 (1.6) [58] | 0.1 (1.9) [36] | 0.3 (1.0) [12] | 0.0 (-0.4, 0.5) | 0.81 |
| | SOC (N=21) | -0.0 (1.3) [20] | 0.0 (1.6) [14] | 0.0 (1.7) [9] | 0.1 (-0.5, 0.7) | 0.75 |
| *Proteus* spp. $log_{10}$CFU | 1% CHG (N=42) | 0.0 (0.0) [39] | 0.0 (0.0) [24] | 0.0 (0.0) [6] | .. | |
| | 2% CHG (N=42) | 0.0 (0.0) [39] | 0.0 (0.0) [21] | 0.0 (0.0) [9] | -0.0 (-0.1, 0.1) | 0.97 |
| | OHP (N=42) | 0.0 (0.0) [39] | 0.1 (0.6) [26] | 0.1 (0.4) [15] | 0.0 (-0.0, 0.1) | 0.23 |
| | Once (N=63) | 0.0 (0.0) [59] | 0.1 (0.5) [35] | 0.1 (0.4) [18] | .. | |
| | Multiple (N=63) | 0.0 (0.0) [58] | 0.0 (0.0) [36] | 0.0 (0.0) [12] | -0.0 (-0.1, 0.0) | 0.29 |
| | SOC (N=21) | 0.0 (0.0) [20] | 0.0 (0.0) [14] | 0.0 (0.0) [9] | -0.0 (-0.1, 0.1) | 0.67 |
| *Acinetobacter* spp. $log_{10}$CFU | 1% CHG (N=42) | 0.1 (0.7) [39] | 0.1 (0.6) [24] | -0.3 (0.7) [6] | .. | |
| | 2% CHG (N=42) | -0.3 (1.1) [39] | -0.2 (0.8) [21] | -0.1 (0.3) [9] | -0.2 (-0.3, 0.0) | 0.096 |
| | OHP (N=42) | 0.0 (1.0) [39] | -0.1 (1.5) [26] | -0.2 (0.6) [15] | 0.0 (-0.2, 0.2) | 0.73 |
| | Once (N=63) | -0.1 (0.9) [59] | 0.0 (1.0) [35] | -0.3 (0.7) [18] | .. | |
| | Multiple (N=63) | -0.0 (0.9) [58] | -0.2 (1.1) [36] | 0.0 (0.0) [12] | 0.0 (-0.1, 0.2) | 0.66 |
| | SOC (N=21) | -0.2 (0.8) [20] | -0.0 (1.3) [14] | -0.1 (1.6) [9] | 0.0 (-0.2, 0.2) | 0.99 |

*CFU = colony forming units; CHG = chlorhexidine' GBS = Group B Streptococcus; OHP = octenidine 0.1% combined with phenoxyethanol 2%; SOC = standard of care*

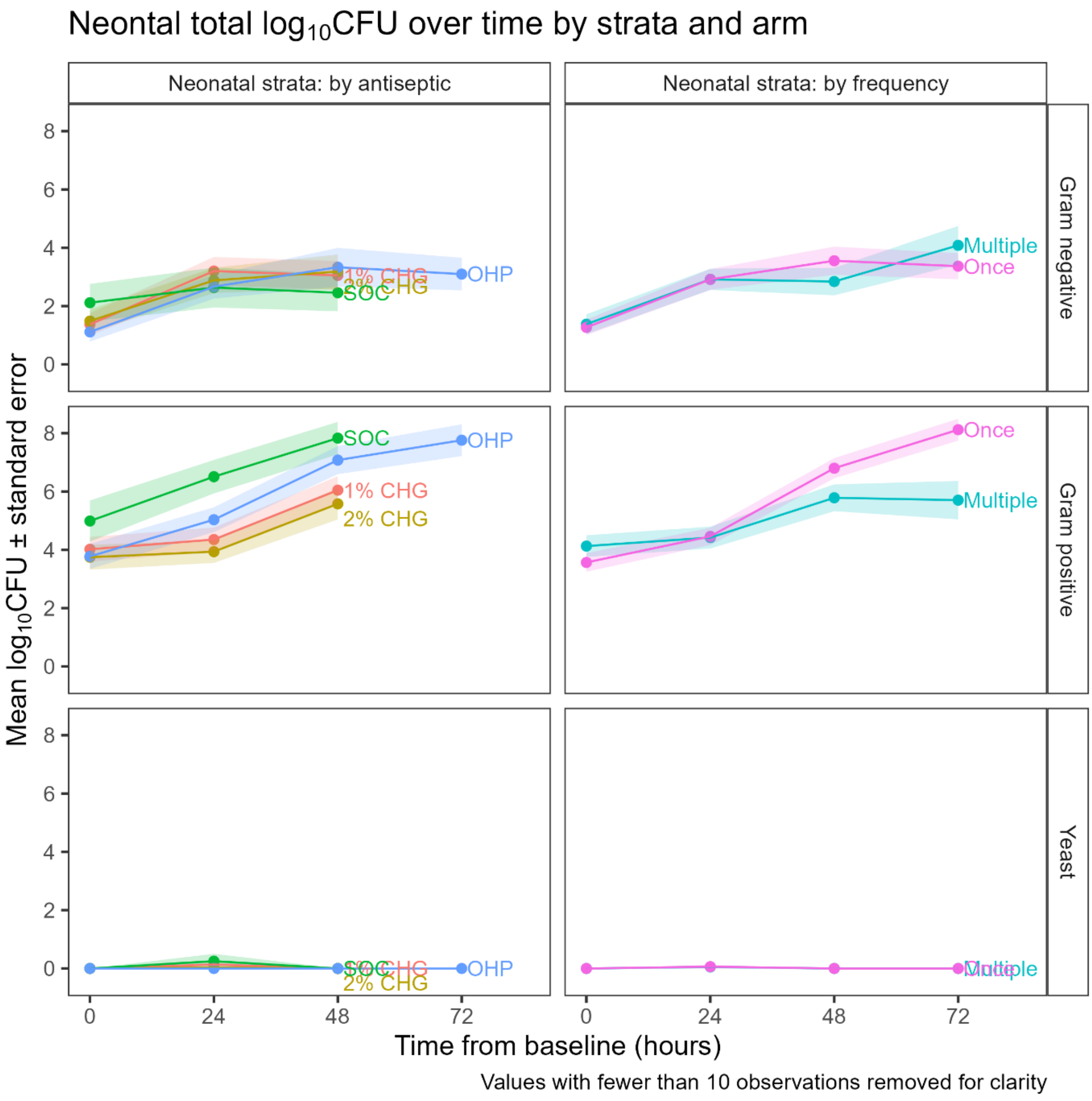

Neontal total log10CFU over time by strata and arm
Neonatal strata: by antiseptic
Neonatal strata: by frequency
Gram negative
Gram positive
Yeast
SOC
OHP
1% CHG
2% CHG
Multiple
Once
Mean log10CFU ± standard error
Time from baseline (hours)
Values with fewer than 10 observations removed for clarity

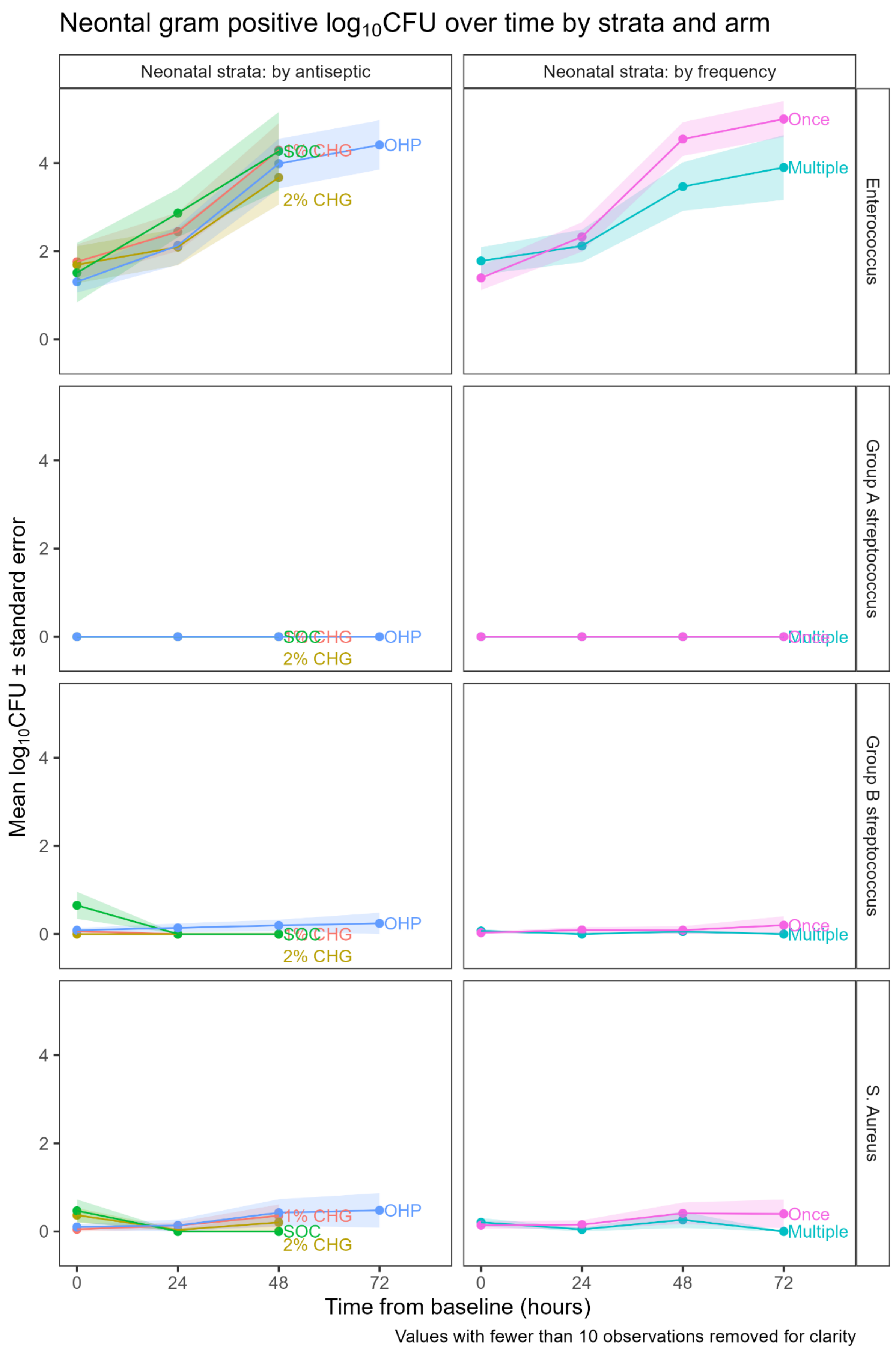
Neontal gram positive log10CFU over time by strata and arm
Neonatal strata: by antiseptic
Neonatal strata: by frequency
Enterococcus
Group A streptococcus
Group B streptococcus
S. Aureus
Mean log10CFU ± standard error
Time from baseline (hours)
0
24
48
72
0
2
4
OHP
SOC
1% CHG
2% CHG
Once
Multiple
Values with fewer than 10 observations removed for clarity

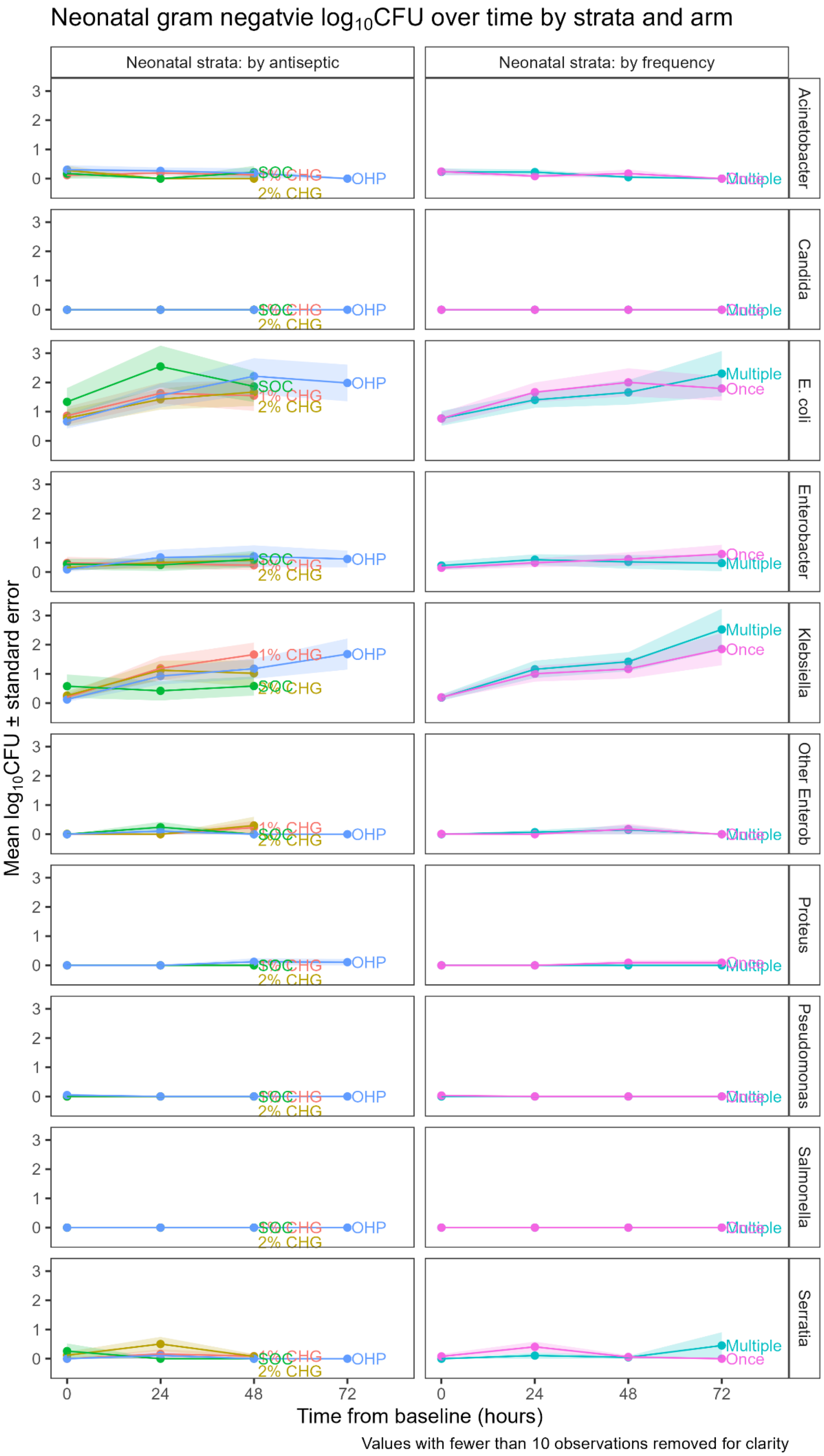

Neonatal gram negatvie log10CFU over time by strata and arm
Neonatal strata: by antiseptic
Neonatal strata: by frequency
Acinetobacter
Candida
E. coli
Enterobacter
Klebsiella
Other Enterob
Proteus
Pseudomonas
Salmonella
Serratia
SOC
1% CHG
2% CHG
OHP
Multiple
Once
Mean log10CFU ± standard error
Time from baseline (hours)
0
24
48
72
Values with fewer than 10 observations removed for clarity

#### 4.4 Evidence for interactions between main terms included in models for change total $log_{10}$CFU (primary outcome)

**Maternal stratum**

| Effect 1 | Effect 2 | p-value of interaction |
|---|---|---|
| drug | frequency | 0.6102 |
| drug | time | 0.6152 |
| frequency | time | 0.1690 |

p-values from interactions fitted one at a time to the focal model

**Neonatal stratum**

| Effect 1 | Effect 2 | p-value of interaction |
|---|---|---|
| drug | frequency | 0.1369 |
| drug | time | 0.9163 |
| frequency | time | 0.0194 |

p-values from interactions fitted one at a time to the focal model. Drug:time interaction added to the model did not converge so unstructured residuals were removed.

Frequency:time interaction added to the model:

| Effect | Estimate | lower 95% CI | upper 95% CI | p-value | p-value of factor |
|---|---|---|---|---|---|
| 2% CHG | -0.27 | -1.18 | 0.635 | 0.556 | 0.0616 |
| OHP | 0.648 | -0.188 | 1.48 | 0.127 | |
| Multiple | 0.747 | -0.59 | 2.08 | 0.271 | |
| SOC | 1.22 | 0.171 | 2.27 | 0.023 | |
| hour | 0.0679 | 0.0424 | 0.0934 | <0.001 | |
| Multiple#hour | -0.0294 | -0.0565 | -0.00229 | 0.034 | |

*CHG = chlorhexidine; OHP = octenidine 0.1% combined with phenoxyethanol 2%; SOC = standard of care*

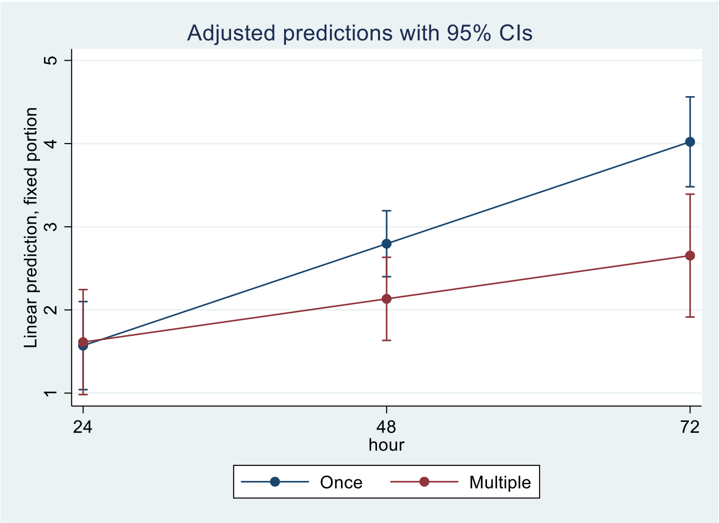

##### 4.5 Evidence for effects of baseline factors on total $log_{10}$CFU and effect modification of intervention effects

**Maternal stratum**

| Variable | Effect/ Interaction | p-value of effect/interaction |
|---|---|---|
| HIV status | Main effect | 0.1166 |
| | Drug interaction | 0.6464 |
| | Frequency interaction | 0.2136 |
| Antibiotics since admission at baseline | Main effect | 0.5730 |
| | Drug interaction (without 2% CHG as none found) | 0.4444 |
| | Frequency interaction | 0.1897 |
| Rupture of membranes at baseline | Main effect | 0.2373 |
| | Drug interaction | 0.7838 |
| | Frequency interaction | 0.6670 |
| Prolonged rupture of membranes at enrolment | Main effect | 0.9917 |
| | Drug interaction | 0.4183 |
| | Frequency interaction | 0.4398 |

p-values with specified subgroups measured at baseline and interaction terms added to the focal model one at a time. Note offensive liquor was pre-specified as a subgroup but none was detected at baseline so analysis could not be performed.
*HIV = human immunodeficiency virus*

**Neonatal stratum**

| Variable | Effect/ Interaction | p-value of effect/interaction |
|---|---|---|
| Maternal HIV status | Main effect | 0.0260 |
| | Drug interaction (with 9 unknown HIV status removed) | 0.1201 |
| | Frequency interaction | 0.1379 |
| Mother received antibiotics in labour | Main effect | 0.3412 |
| | Drug interaction | 0.8815 |
| | Frequency interaction | 0.2687 |
| Prolonged rupture of membranes | Main effect | 0.2685 |
| | Drug interaction* | 0.0917 |
| | Frequency interaction | 0.2872 |
| Offensive liquor | Main effect | 0.4161 |
| | Drug interaction | 0.2649 |
| | Frequency interaction | 0.8107 |
| Gestational age at enrolment | Main effect | 0.624 |
| | Drug interaction* | 0.9358 |
| | Frequency interaction | 0.9017 |
| Admission ward | Main effect | 0.0398 |
| | Drug interaction | 0.0959 |
| | Frequency interaction | 0.0160 |
| Antibiotics before enrolment | Main effect | 0.0033 |
| | Drug interaction | 0.0655 |
| | Frequency interaction* | 0.0315 |
| CHG cord care | Main effect | 0.6937 |

| Variable | Effect/ Interaction | p-value of effect/interaction |
|---|---|---|
| | Drug interaction | 0.4390 |
| | Frequency interaction | 0.8032 |

Values with specified subgroups measured at baseline and interaction terms added to the focal model one at a time.
*Unstructured residuals excluded from the model due to model not converging.
HIV status coefficient = 1.5 (95% CI: 0.2, 2.9) for positive (higher) vs negative (lower)
*CHG = chlorhexidine; HIV = human immunodeficiency virus*

##### ADMISSION WARD # FREQUENCY INTERACTION

| Effect | Estimate | lower 95% CI | upper 95% CI | p-value | p-value of factor |
|---|---|---|---|---|---|
| 2% CHG | -0.264 | -1.16 | 0.633 | 0.562 | 0.0584 |
| OHP | 0.662 | -0.173 | 1.5 | 0.119 | |
| Multiple | 0.997 | -0.411 | 2.4 | 0.163 | |
| SOC | 1.28 | 0.221 | 2.34 | 0.018 | |
| Nursery | 0.00216 | -1.14 | 1.14 | 0.997 | |
| Nursery HDU | -0.0101 | -1.11 | 1.09 | 0.985 | |
| Nursery#multiple | -2.34 | -4.11 | -0.571 | 0.01 | |
| Nursery HDU# multiple | -1.77 | -3.48 | -0.066 | 0.042 | |

Note: reference ward was Postnatal ward with mother. Postnatal ward with mother 66 (25%), Nursery 90 (33%), Nursery HDU 113 (42%).
*CHG = chlorhexidine; HDU = high dependency unit; OHP = octenidine 0.1% combined with phenoxyethanol 2%; SOC = standard of care*

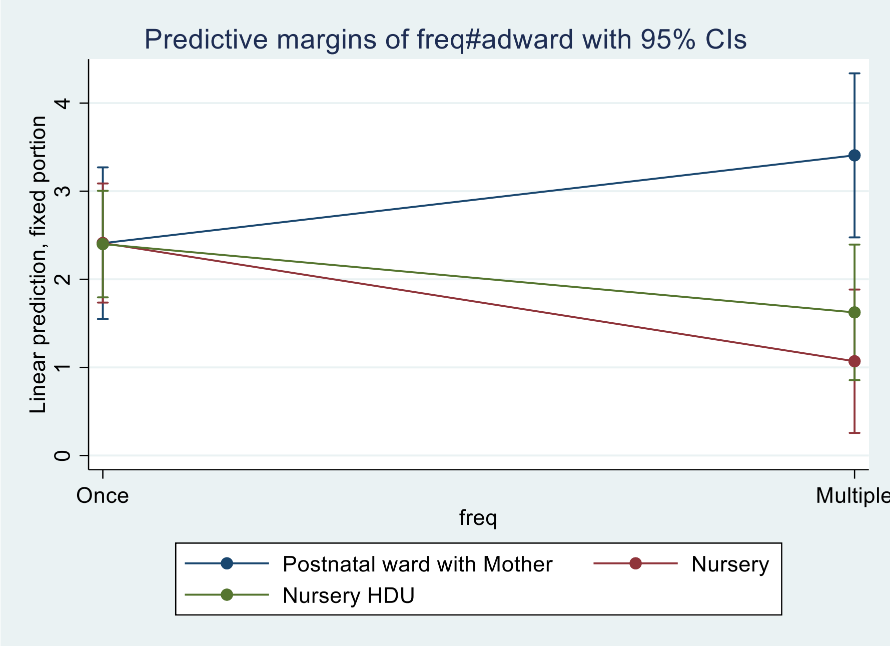

**ANTIBIOTICS BEFORE ENROLMENT # FREQUENCY INTERACTION**

| Effect | Estimate | lower 95% CI | upper 95% CI | p-value |
|---|---|---|---|---|
| 2% CHG | -0.142 | -1 | 0.719 | 0.745 |
| OHP | 0.834 | -0.00628 | 1.67 | 0.052 |
| Multiple | 0.171 | -0.601 | 0.942 | 0.662 |
| SOC | 1.51 | 0.439 | 2.59 | 0.006 |
| Antibiotics before enrolment | -0.314 | -1.33 | 0.7 | 0.54 |
| Antibiotics before enrolment#Multiple | -1.68 | -3.22 | -0.131 | 0.034 |

Antibiotics before enrolment: 65 (24%)

*CHG = chlorhexidine; OHP = octenidine 0.1% combined with phenoxyethanol 2%; SOC = standard of care*

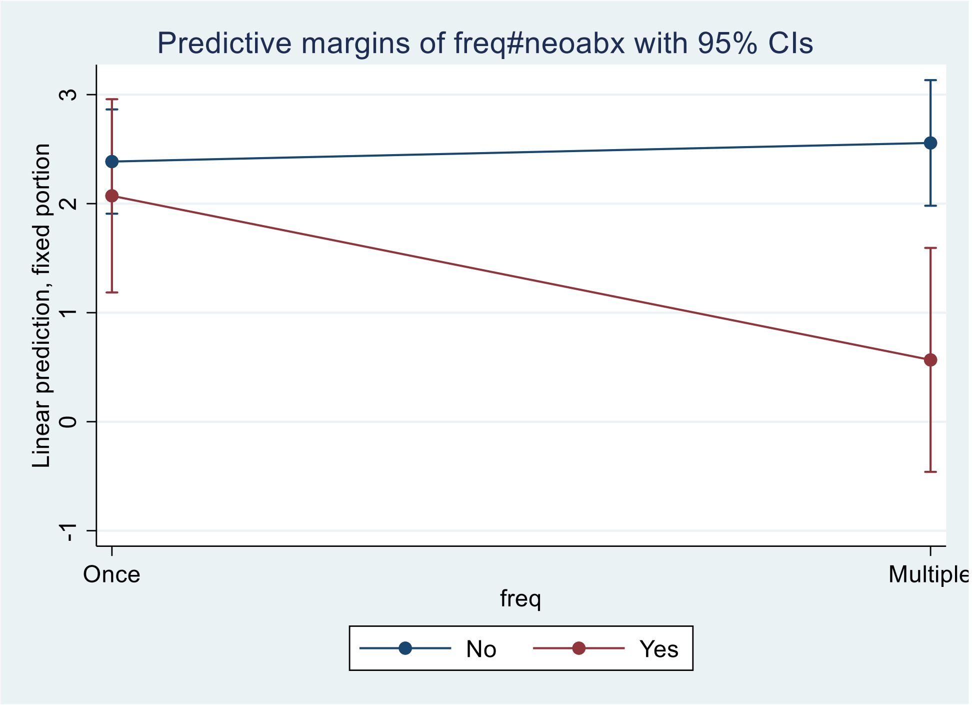

**Neonates born to mothers in the maternal stratum**

| Variable | Effect/ Interaction | p-value of effect/interaction |
|---|---|---|
| HIV status | Main effect | 0.0349 |
| | Drug interaction | 0.9116 |
| | Frequency interaction | 0.6582 |
| Mode of delivery | Main effect (forceps/vacuum assisted removed as observed only once) | 0.0243 |
| | Drug interaction (forceps/vacuum assisted removed as observed only once) | 0.7680 |
| | Frequency interaction (forceps/vacuum assisted removed as observed only once) | 0.6104 |

p-values with specified subgroups measured at baseline and interaction terms added to the focal model one at a time.
HIV status coefficient = -1.9 (95% CI: -3.7, -0.1) for positive (lower) vs negative (higher)
Model of delivery coefficient = 1.7 (95% CI: 0.2, 3.2) for spontaneous vaginal (higher) vs Emergency c-section (lower)
HIV status coefficient for spontaneous vaginal only = -2.5 (95% CI: -4.7, -0.4; p = 0.020) for positive (lower) vs negative (higher)
*HIV = human immunodeficiency virus*

##### 4.6 ACCEPT analyses of efficacy primary outcome (change in $\log_{10}$CFU)

Probability treatment effect compared to 1% CHG/once is more negative than acceptability threshold under different priors. E.g under non-informative priors

#### Maternal stratum

- Probability 2% CHG is better than 1% CHG is 94%, and the probability is better by at least 0.5 $\log_{10}$CFU is 62%
- Probability OHP is better than 1% CHG is 0%
- Probability multiple is better than once is 90%, and probably is better by at least 0.5 $\log_{10}$CFU is 39%
- Probability SOC is better than 1% CHG once is 0%

See the Statistical analysis plan for further details of priors

| Factor | Acceptability threshold | Non-informative | Optimistic | Sceptical |
|---|---|---|---|---|
| 2% CHG | -2 | 0% | 2% | 0% |
| | -1 | 17% | 79% | 61% |
| | -0.5 | 62% | 99% | 96% |
| | 0 | 94% | 100% | 100% |
| | 0.5 | 100% | 100% | 100% |
| | 1 | 100% | 100% | 100% |
| | 2 | 100% | 100% | 100% |
| OHP | -2 | 0% | 0% | 0% |
| | -1 | 0% | 0% | 0% |
| | -0.5 | 0% | 0% | 0% |
| | 0 | 0% | 2% | 0% |
| | 0.5 | 0% | 26% | 12% |
| | 1 | 5% | 79% | 60% |
| | 2 | 80% | 100% | 100% |
| Multiple applications | -2 | 0% | 0% | 0% |
| | -1 | 4% | 7% | 2% |
| | -0.5 | 39% | 68% | 45% |
| | 0 | 90% | 100% | 97% |
| | 0.5 | 100% | 100% | 100% |
| | 1 | 100% | 100% | 100% |
| | 2 | 100% | 100% | 100% |
| SOC | -2 | 0% | 0% | 0% |
| | -1 | 0% | 0% | 0% |
| | -0.5 | 0% | 0% | 0% |
| | 0 | 0% | 0% | 0% |
| | 0.5 | 0% | 0% | 3% |
| | 1 | 0% | 12% | 32% |
| | 2 | 0% | 97% | 99% |

*CHG = chlorhexidine; OHP = octenidine 0.1% combined with phenoxyethanol 2%; SOC = standard of care*

**Neonatal stratum**

- Probability 2% CHG is better than 1% CHG is 63%, and the probability is better by at least 0.5 $log_{10}$CFU is 22%
- Probability OHP is better than 1% CHG is 4%
- Probability multiple is better than once is 72%, and probably is better by at least 0.5 $log_{10}$CFU is 22%.
- Probability SOC is better than 1% CHG once is 1%.

See the Statistical Analysis Plan for further details of priors

| Factor | Acceptability threshold | Non-informative | Optimistic | Sceptical |
|---|---|---|---|---|
| 2% CHG | -2 | 0% | 0% | 0% |
| | -1 | 3% | 21% | 4% |
| | -0.5 | 22% | 74% | 40% |
| | 0 | 63% | 99% | 89% |
| | 0.5 | 93% | 100% | 100% |
| | 1 | 99% | 100% | 100% |
| | 2 | 100% | 100% | 100% |
| OHP | -2 | 0% | 0% | 0% |
| | -1 | 0% | 0% | 0% |
| | -0.5 | 0% | 5% | 1% |
| | 0 | 4% | 45% | 16% |
| | 0.5 | 24% | 91% | 68% |
| | 1 | 67% | 100% | 97% |
| | 2 | 100% | 100% | 100% |
| Multiple applications | -2 | 0% | 0% | 0% |
| | -1 | 2% | 0% | 0% |
| | -0.5 | 22% | 36% | 9% |
| | 0 | 72% | 97% | 79% |
| | 0.5 | 97% | 100% | 100% |
| | 1 | 100% | 100% | 100% |
| | 2 | 100% | 100% | 100% |
| SOC | -2 | 0% | 0% | 0% |
| | -1 | 0% | 0% | 0% |
| | -0.5 | 0% | 0% | 0% |
| | 0 | 1% | 1% | 8% |
| | 0.5 | 4% | 32% | 70% |
| | 1 | 20% | 92% | 99% |
| | 2 | 82% | 100% | 100% |

*CHG = chlorhexidine; OHP = octenidine 0.1% combined with phenoxyethanol 2%; SOC = standard of care*

#### 4.7 Sensitivity analyses for total $log_{10}$CFU

All analyses are as per the primary analysis either restricted to one sample only (and day of sampling removed from the model) or with additional factors added. Estimates are versus the **1% CHG, applied once** reference group from a normal linear regression, adjusted for arm, s, and time of sample where required.

##### REPEATED SAMPLES SENSITIVITY ANALYSIS

**First outcome measure after baseline**

##### Maternal stratum

| Effect | Estimate | lower 95% CI | upper 95% CI | p-value | p-value of factor |
|---|---|---|---|---|---|
| 2% CHG | -0.608 | -1.42 | 0.204 | 0.141 | <0.001 |
| OHP | 2.09 | 1.28 | 2.91 | <0.001 | |
| Multiple | 0.241 | -0.423 | 0.905 | 0.473 | |
| SOC | 4.5 | 3.37 | 5.63 | <0.001 | |

First swab timepoint: 4h: 123 (100%)

*CHG = chlorhexidine; OHP = octenidine 0.1% combined with phenoxyethanol 2%; SOC = standard of care*

##### Neonatal stratum

| Effect | Estimate | lower 95% CI | upper 95% CI | p-value | p-value of factor |
|---|---|---|---|---|---|
| 2% CHG | -0.287 | -1.37 | 0.796 | 0.601 | 0.2283 |
| OHP | 0.638 | -0.44 | 1.72 | 0.244 | |
| Multiple | 0.161 | -0.726 | 1.05 | 0.72 | |
| SOC | 1.76 | 0.375 | 3.15 | 0.013 | |

*CHG = chlorhexidine; OHP = octenidine 0.1% combined with phenoxyethanol 2%; SOC = standard of care*

**Final outcome measure**

##### Maternal stratum

| Effect | Estimate | lower 95% CI | upper 95% CI | p-value | p-value of factor |
|---|---|---|---|---|---|
| 2% CHG | -0.78 | -1.76 | 0.197 | 0.116 | <0.001 |
| OHP | 1.14 | 0.15 | 2.12 | 0.024 | |
| Multiple | -0.777 | -1.57 | 0.0204 | 0.056 | |
| SOC | 3.1 | 1.75 | 4.46 | <0.001 | |

*CHG = chlorhexidine; OHP = octenidine 0.1% combined with phenoxyethanol 2%; SOC = standard of care*

##### Neonatal stratum

| Effect | Estimate | lower 95% CI | upper 95% CI | p-value | p-value of factor |
|---|---|---|---|---|---|
| 2% CHG | 0.252 | -0.658 | 1.16 | 0.585 | 0.0745 |
| OHP | 1.02 | 0.109 | 1.93 | 0.029 | |
| Multiple | -0.0888 | -0.833 | 0.656 | 0.814 | |
| SOC | 1.8 | 0.623 | 2.98 | 0.003 | |

*CHG = chlorhexidine; OHP = octenidine 0.1% combined with phenoxyethanol 2%; SOC = standard of care*

##### 4.8 Analysis of serious adverse events (SAEs)

SAEs are presented as N (%) M where N is the number of participants experiencing at least one SAE, % is the associated percentage of participants and M is the total number of events.

**Maternal stratum**

(a) by randomisation to antiseptic or SOC

| Variable | Overall (N=149) | 1% CHG (N=43) | 2% CHG (N=43) | OHP (N=42) | SOC (N=21) |
|---|---|---|---|---|---|
| Overall SAEs | 7 (5%) 7 | 3 (7%) 3 | 2 (5%) 2 | 1 (2%) 1 | 1 (5%) 1 |
| Death | 5 (3%) 5 | 2 (5%) 2 | 1 (2%) 1 | 1 (2%) 1 | 1 (5%) 1 |
| Prolonged hospitalisation | 2 (1%) 2 | 1 (2%) 1 | 1 (2%) 1 | 0 (0%) 0 | 0 (0%) 0 |
| Congenital Anomaly | 1 (1%) 1 | 0 (0%) 0 | 1 (2%) 1 | 0 (0%) 0 | 0 (0%) 0 |

*CHG = chlorhexidine; OHP = octenidine 0.1% combined with phenoxyethanol 2%; SAE = serious adverse event; SOC = standard of care*

(b) by randomisation frequency (excluding SOC)

| Variable | Once (N=64) | Multiple (N=64) |
|---|---|---|
| Overall SAEs | 4 (6%) 4 | 2 (3%) 2 |
| Death | 2 (3%) 2 | 2 (3%) 2 |
| Prolonged hospitalisation | 2 (3%) 2 | 0 (0%) 0 |
| Congenital Anomaly | 1 (2%) 1 | 0 (0%) 0 |

**ANALYSIS OF OVERALL SAEs**

| Arm | Odds ratio estimate | lower 95% CI | upper 95% CI | p-value |
|---|---|---|---|---|
| CHG 2% | 0.66 | 0.05 | 6.11 | 1 |
| OHP | 0.33 | 0.01 | 4.32 | 0.640 |
| Multiple | 0.49 | 0.04 | 3.55 | 0.689 |
| SOC | 0.50 | 0.01 | 7.33 | 0.99 |

Estimates showing comparison to 1% CHG, applied once from an exact logistic regression.
*CHG = chlorhexidine; OHP = octenidine 0.1% combined with phenoxyethanol 2%; SAE = serious adverse event; SOC = standard of care*

**SAEs by PT**

(a) by randomisation to antiseptic or SOC

| Variable | 1% CHG (N=43) | 2% CHG (N=43) | OHP (N=42) | SOC (N=21) |
|---|---|---|---|---|
| Congenital, familial and genetic disorders /Congenital anomaly | 0 (0%) 0 | 1 (2%) 1 | 0 (0%) 0 | 0 (0%) 0 |
| Infections and infestations /Sepsis neonatal | 0 (0%) 0 | 0 (0%) 0 | 1 (2%) 1 | 1 (5%) 1 |
| Pregnancy, puerperium and perinatal conditions /Foetal death | 1 (2%) 1 | 0 (0%) 0 | 0 (0%) 0 | 0 (0%) 0 |
| Pregnancy, puerperium and perinatal conditions /Postpartum haemorrhage | 1 (2%) 1 | 0 (0%) 0 | 0 (0%) 0 | 0 (0%) 0 |
| Pregnancy, puerperium and perinatal conditions /Stillbirth | 0 (0%) 0 | 1 (2%) 1 | 0 (0%) 0 | 0 (0%) 0 |
| Respiratory, thoracic and mediastinal disorders /Neonatal asphyxia | 1 (2%) 1 | 0 (0%) 0 | 0 (0%) 0 | 0 (0%) 0 |

*CHG = chlorhexidine; OHP = octenidine 0.1% combined with phenoxyethanol 2%; SAE = serious adverse event; SOC = standard of care*

(b) by randomisation frequency (excluding SOC)

| Variable | Once (N=64) | Multiple (N=64) |
|---|---|---|
| Congenital, familial and genetic disorders /Congenital anomaly | 1 (2%) 1 | 0 (0%) 0 |
| Infections and infestations /Sepsis neonatal | 1 (2%) 1 | 0 (0%) 0 |
| Pregnancy, puerperium and perinatal conditions /Foetal death | 1 (2%) 1 | 0 (0%) 0 |
| Pregnancy, puerperium and perinatal conditions /Postpartum haemorrhage | 1 (2%) 1 | 0 (0%) 0 |
| Pregnancy, puerperium and perinatal conditions /Stillbirth | 0 (0%) 0 | 1 (2%) 1 |
| Respiratory, thoracic and mediastinal disorders /Neonatal asphyxia | 0 (0%) 0 | 1 (2%) 1 |

**Neonatal stratum**

(a) by randomisation to antiseptic or SOC

| Variable | Overall (N=147) | 1% CHG (N=42) | 2% CHG (N=42) | OHP (N=42) | SOC (N=21) |
|---|---|---|---|---|---|
| Overall SAEs | 16 (11%) 17 | 4 (10%) 4 | 7 (17%) 8 | 2 (5%) 2 | 3 (14%) 3 |
| Death | 9 (6%) 9 | 2 (5%) 2 | 3 (7%) 3 | 1 (2%) 1 | 3 (14%) 3 |
| Life threatening | 3 (2%) 3 | 1 (2%) 1 | 1 (2%) 1 | 0 (0%) 0 | 1 (5%) 1 |
| Prolonged hospitalisation | 7 (5%) 8 | 2 (5%) 2 | 3 (7%) 4 | 1 (2%) 1 | 1 (5%) 1 |

*CHG = chlorhexidine; OHP = octenidine 0.1% combined with phenoxyethanol 2%; SAE = serious adverse event; SOC = standard of care*

(b) by randomisation frequency (excluding SOC)

| Variable | Once (N=63) | Multiple (N=63) |
|---|---|---|
| Overall SAEs | 5 (8%) 6 | 8 (13%) 8 |
| Death | 2 (3%) 2 | 4 (6%) 4 |
| Life threatening | 1 (2%) 1 | 1 (2%) 1 |
| Prolonged hospitalisation | 2 (3%) 3 | 4 (6%) 4 |

**ANALYSIS OF OVERALL SAEs**

| Arm | Odds ratio estimate | lower 95% CI | upper 95% CI | p-value |
|---|---|---|---|---|
| CHG 2% | 1.88 | 0.43 | 9.54 | 0.520 |
| OHP | 0.48 | 0.04 | 3.57 | 0.676 |
| Multiple | 1.6 | 0.45 | 7.03 | 0.558 |
| SOC | 2.05 | 0.23 | 16.74 | 0.691 |

*CHG = chlorhexidine; OHP = octenidine 0.1% combined with phenoxyethanol 2%; SAE = serious adverse event; SOC = standard of care*

(a) by randomisation to antiseptic or SOC

| Variable | 1% CHG (N=42) | 2% CHG (N=42) | OHP (N=42) | SOC (N=21) |
|---|---|---|---|---|
| Infections and infestations /Pneumonia | 0 (0%) 0 | 2 (5%) 2 | 0 (0%) 0 | 0 (0%) 0 |
| Infections and infestations /Sepsis neonatal | 3 (7%) 3 | 3 (7%) 3 | 1 (2%) 1 | 1 (5%) 1 |
| Pregnancy, puerperium and perinatal conditions /Hypoxic ischaemic encephalopathy neonatal | 0 (0%) 0 | 3 (7%) 3 | 0 (0%) 0 | 1 (5%) 1 |
| Pregnancy, puerperium and perinatal conditions /Neonatal asphyxia | 1 (2%) 1 | 0 (0%) 0 | 1 (2%) 1 | 0 (0%) 0 |
| Respiratory, thoracic and mediastinal disorders /Neonatal respiratory distress syndrome | 0 (0%) 0 | 0 (0%) 0 | 0 (0%) 0 | 1 (5%) 1 |

(b) by randomisation frequency (excluding SOC)

| Variable | Once (N=63) | Multiple (N=63) |
|---|---|---|
| Infections and infestations /Pneumonia | 1 (2%) 1 | 1 (2%) 1 |
| Infections and infestations /Sepsis neonatal | 3 (5%) 3 | 4 (6%) 4 |
| Pregnancy, puerperium and perinatal conditions /Hypoxic ischaemic encephalopathy neonatal | 2 (3%) 2 | 1 (2%) 1 |
| Pregnancy, puerperium and perinatal conditions /Neonatal asphyxia | 0 (0%) 0 | 2 (3%) 2 |
| Respiratory, thoracic and mediastinal disorders /Neonatal respiratory distress syndrome | 0 (0%) 0 | 0 (0%) 0 |

##### 4.9 Line listing of SAEs

**Maternal SAEs**

Note: no mothers experienced multiple SAEs

| SAE Ref | Trial Arm | Main Diagnosis | Severity | Relatedness | Event Outcome |
|---|---|---|---|---|---|
| 1 | CHG 1% Multiple | Birth asphyxia | Grade 5 (Death) | Unrelated | Death |
| 2 | CHG 1% Once | Post partum haemorrhage | Grade 2 (Moderate) | Unrelated | Resolved |
| 3 | CHG 1% Once | Neonatal Intrauterine Death (fresh still birth) due to missed Transverse Lie | Grade 5 (Death) | Unrelated | Death |
| 4 | SOC | Neonatal Sepsis | Grade 5 (Death) | Unrelated | Death |
| 5 | CHG 2% Once | Congenital Abnormality | Grade 2 (Moderate) | Unrelated | Resolved with sequelae |
| 6 | OHP Once | Neonatal sepsis | Grade 5 (Death) | Unlikely to be related | Death |
| 7 | CHG 2% Multiple | Stillbirth | Grade 5 (Death) | Unrelated | Death |

*CHG = chlorhexidine; IMP = investigational medicinal product; OHP = octenidine 0.1% combined with phenoxyethanol 2%; SAE = serious adverse event; SOC = standard of care*

**Neonatal SAEs**

| SAE Ref | SAE num | Trial Arm | Main Diagnosis | Severity | Relatedness | Event Outcome |
|---|---|---|---|---|---|---|
| 1 | 1 | CHG 2% Once | Pneumonia | Grade 2 (Moderate) | Unrelated | Resolved |
| 2 | 1 | CHG 2% Multiple | Neonatal sepsis | Grade 2 (Moderate) | Unrelated | Resolved |
| 3 | 1 | OHP Multiple | Neonatal sepsis | Grade 2 (Moderate) | Unrelated | Resolved |
| 4 | 1 | SOC | Neonatal Sepsis | Grade 5 (Death) | Unrelated | Death |
| 5 | 1 | CHG 2% Once | Neonatal sepsis | Grade 3 (Severe) | Unrelated | Resolved |
| 5 | 2 | CHG 2% Once | hypoxic ischaemic encephalopathy | Grade 3 (Severe) | Unrelated | Resolved with sequalae |
| 6 | 1 | CHG 1% Multiple | Birth asphyxia | Grade 5 (Death) | Unrelated | Death |
| 7 | 1 | CHG 2% Once | hypoxic ischaemic encephalopathy | Grade 4 (Life-threatening) | Unrelated | Death |
| 8 | 1 | CHG 1% Multiple | Neonatal sepsis (Klebsiella pneumoniae) | Grade 2 (Moderate) | Unrelated | Death |
| 9 | 1 | CHG 1% Once | Neonatal sepsis | Grade 5 (Death) | Unrelated | Death |
| 10 | 1 | SOC | Respiratory distress syndrome | Grade 5 (Death) | Unrelated | Death |
| 11 | 1 | SOC | Neonatal death secondary to severe hypoxic ischaemic encephalopathy | Grade 5 (Death) | Unrelated | Death |

*CHG = chlorhexidine; IMP = investigational medicinal product; OHP = octenidine 0.1% combined with phenoxyethanol 2%; SAE = serious adverse event; SOC = standard of care*

#### 5. eReferences

1. Lund CH, Osborne JW. Validity and reliability of the Neonatal Skin Condition score. *JOGNN - Journal of Obstetric, Gynecologic, and Neonatal Nursing*. 2004;33(3):320-327. doi:10.1177/0884217504265174
2. Harwood-Nuss A, Wolfson A LC. *The Clinical Practice of Emergency Medicine*. (Lippincott-Raven;, ed.).; 1996.
3. U.S. Department of Health and Human Services. National Cancer Institute Common Terminology Criteria for Adverse Events (CTCAE). Version 5. November 27, 2017. Accessed August 7, 2025. https://www.ctc.ucl.ac.uk/TrialDocuments/Uploaded/Common%20Terminology%20Criteria%20for%20Adverse%20Events%20(CTCAE)%20v5.0_14092023_0.pdf
4. IMPAACT P1106: Pharmacokinetic Characteristics of Antiretrovirals and Tuberculosis Medicines in Low Birth Weight Infants. Accessed August 7, 2025. https://clinicaltrials.gov/study/NCT02383849
